# Thromboembolic risk in lung cancer patients receiving systemic therapy

**DOI:** 10.1101/2020.12.21.20248675

**Authors:** Cecelia J. Madison, Ryan A. Melson, Michael J. Conlin, Kenneth R. Gundle, Reid F. Thompson, David C. Calverley

## Abstract

In this retrospective study, we investigated the influence of chemotherapy and immunotherapy on thromboembolic risk among US Veterans with lung cancer during their first six months of systemic therapy. Patients in the study cohort received treatment with common frontline agents that were divided into four groups: chemotherapy alone, immunotherapy alone, combination of chemo- and immunotherapies, and molecularly targeted therapies. The latter served as a control group of systemically treated lung cancer patients who received neither chemotherapy nor immunotherapy. The cohort experienced a 6.8% overall incidence of thromboembolic events with a median time to event of 49 days, but the analysis demonstrated significantly different rates among the different treatment groups. We explored models incorporating multiple confounding variables as well as the competing risk of death, and these results indicated that both chemotherapy and immunotherapy were associated with an increased incidence of thrombosis, either when given alone or combined, compared with the control group (6.91%, 9.09%, and 7.47% respectively versus 3.68%, p < 0.024). Both the Khorana score assessing thrombosis risk for cancer patients and the Charlson comorbidity score were found to be associated with increased risk of thrombosis in our analyses. Paradoxically, we found an association between risk of thrombosis and the use of prophylactic anticoagulation or aspirin during the first month of systemic treatment, accounting for several confounding variables including a patient’s prior history of thrombosis. Additionally, our data suggest that thromboembolic events may occur more commonly in lung cancer patients treated with immunotherapy compared with chemotherapy. Further study is warranted to better determine the drivers of thromboembolic risk and to identify ways to mitigate this risk for patients.

## Introduction

Venous thromboembolism (VTE) is a common complication of cancer with an incidence of 1-20% (1). While randomized trials have shown prophylactic anticoagulation roughly halves the relative risk of cancer-associated VTE, the absolute risk reduction among ambulatory patients with cancer is relatively modest, given that the baseline VTE risk in this population is about 3-5% in the first months of chemotherapy (2). Bleeding complications associated with anticoagulation are more common in cancer patients compared with those without cancer, and in patients at low risk of thromboembolism, this means that the risks of anticoagulation may exceed the benefits. Thus, risk stratification is an important consideration when deciding whether to recommend prophylactic anticoagulation in the cancer setting.

Arterial and venous thromboembolic events (TEEs) in cancer patients are associated with several known risk factors such as age, type and stage of cancer, hospitalization, and indwelling catheters. Active chemotherapy treatment has been shown to increase the incidence of TEEs in patients with breast cancer (3, 4) and lung cancer (5, 6). While cancer itself was associated with a 4.1-fold increase in the risk of thrombosis, the addition of chemotherapy increased the risk to 6.5 times the baseline level in a large population-based case control study (7). Lung cancer is among the most common cancer types; an estimated 2.1 million new cases were diagnosed in 2018, resulting in an estimated 1.8 million deaths (8). Over half of newly diagnosed patients are considered incurable due to the presence of metastatic disease (9) but are eligible for systemic therapy.

Current standard of care for first line treatment of advanced stage non-small cell lung cancer (NSCLC) varies depending on histology, the presence or absence of driver mutations, and programmed death ligand-1 (PD-L1) expression levels (10-12). Approximately 25-40% of NSCLCs have a PD-L1 tumor proportional score (TPS) of 50% or higher (13). Currently recommended first-line therapies for patients with higher PD-L1 TPS levels and without a driver mutation are either a single agent checkpoint inhibitor (e.g. pembrolizumab, nivolumab) or checkpoint inhibitor plus pemetrexed and platinum regimen. For patients with a confirmed and actionable driver mutation, first line therapy may employ molecularly targeted agents (e.g. crizotinib, erlotinib).

In this large retrospective population-based cohort study, we determined the TEE incidence and identified associated VTE risk factors among 95,466 lung cancer patients receiving systemic therapy.

## Methods

We performed a retrospective cohort study of US national health records available within the Veterans Health Administration’s (VA) Corporate Data Warehouse (CDW). Our VA CDW derived database contains individually identifiable clinical and demographic information from the 1990s through 2020 for ∼19 million individual Veterans who received care provided or paid for by the VA. All work was performed under local IRB approval.

The inclusion criteria for this study were a diagnosis of a primary malignant neoplasm of lung or bronchus and subsequent treatment with any of fourteen systemic therapies. The specific chemotherapy agents studied were cisplatin, carboplatin, paclitaxel, docetaxel, pemetrexed, etoposide, and gemcitabine, and the immunotherapy agents studied were pembrolizumab and nivolumab. The molecularly targeted therapies included were crizotinib, alectinib, gefitinib, erlotinib, and afatinib. Diagnoses were identified in the database using ICD-9 or ICD-10-CM codes (Appendix 1), and therapeutic agents were identified by name using RxNorm (14). Using custom Structured Query Language (SQL) and R scripts (Appendix 2), 95,466 Veterans meeting these criteria were identified. We divided the overall cohort into four groups based on type of initial systemic therapy administered: 1) conventional chemotherapy, 2) immunotherapy, 3) combination chemo- and immunotherapies, and 4) molecularly targeted therapy (non-chemo non-immunotherapy control group). Characteristics of the overall patient cohort are shown in Table 1.

**Table 1.** Patient Characteristics. Characteristics of the 95,466 Veteran cohort. P-values were computed by Wilcox test.

|  | **Chemotherapy**<br>**alone**<br>**n = 83,981** | **Immunotherapy**<br>**alone**<br>**n = 2,574** | **Combined**<br>**therapies**<br>**n = 3,347** | **Control**<br>**n = 5,564** | **p-value** |
| --- | --- | --- | --- | --- | --- |
| **Male** | 81,996 (97.6%) | 2,501 (97.2%) | 3,260 (97.4%) | 5,428 (97.6%) | p > 0.1 |
| **Age (years)**<br>**median ± MAD** | 66 ± 8.90 | 71 ± 7.41 | 69 ± 5.93 | 75 ± 10.4 | p < 0.001 |
| **Year of**<br>**diagnosis**<br>**median ± MAD** | 2009 ± 7.4 | 2017 ± 1.5 | 2018 ± 1.5 | 2007 ± 4.4 | p < 0.001 |
| **Months**<br>**between**<br>**diagnosis and**<br>**treatment**<br>**mean ± SD** | 6.32 ± 19.2 | 16.6 ± 31.2 | 8.33 ± 22.8 | 11.7 ± 20.4 | p < 0.001 |
| **Khorana Score**<br>**mean ± SD** | 1.63 ± 0.79 | 1.63 ± 0.82 | 1.63 ± 0.79 | 1.53 ± 0.75 | p < 0.03<sup>a</sup> |
| **Charlson**<br>**Comorbidity**<br>**Index**<br>**mean ± SD** | 9.78 ± 4.1 | 12.4 ± 4.3 | 11.6 ± 3.9 | 9.19 ± 3.9 | p < 0.001 |
| **BMI**<br>**mean ± SD** | 25.9 ± 5.5 | 26.4 ± 5.8 | 26.2 ± 5.4 | 25.8 ± 5.1 | p < 0.009<sup>b</sup> |
| **Prior TEE** | 7,001 (8.34%) | 481 (18.7%) | 443 (13.2%) | 426 (7.66%) | p < 0.001<sup>c</sup> |
| **Prophylactic**<br>**Anticoagulation** | 5,812 (6.92%) | 262 (10.2%) | 288 (8.60%) | 390 (7.01%) | p < 0.02<sup>c</sup> |
| **Prophylactic**<br>**Aspirin** | 7,075 (8.42%) | 182 (7.07%) | 248 (7.41%) | 284 (5.10%) | p < 0.03<sup>d</sup> |

<sup>a</sup> Control group differed from other three only
<sup>b</sup> Chemotherapy and Control groups did not differ from each other; Immunotherapy and Combined groups did not differ from each other
<sup>c</sup> Chemotherapy and Control groups did not differ from each other
<sup>d</sup> Immunotherapy and Combined groups did not differ from each other

Initial cancer diagnosis was defined as the first occurrence in the patient’s record of one of the specified lung cancer diagnoses, and elapsed time until treatment was calculated using the initial diagnosis date and the beginning of systemic therapy with any of this study’s specified agents.

Incidence of and number of days to any TEE was assessed for a 180-day period following a patient’s first treatment with one of the specified systemic therapies. TEEs were defined as deep vein thrombosis, pulmonary embolism, stroke, or myocardial infarction and identified in the database using ICD-9 or ICD-10-CM codes (Appendix 1). Patients who experienced two or more TEEs simultaneously were counted as a single event, and for those who experienced multiple events at different times, only the first event was considered. History of prior TEE was defined as the occurrence in the patient’s record of any of the specified TEE diagnoses at any time before the first day of systemic therapy with any of this study’s specified agents. Patients who experienced a TEE on the first day of systemic therapy were excluded.

The 5,377 patients lost to follow up during the study period were censored from the analyses at the time of last follow up. Of the patients who did not die during the study, the average follow-up time was 179 days.

We calculated the Khorana score (as per Table 2) for prediction of VTE risk for our cohort (15). Pre-treatment platelet counts, hemoglobin levels, and leukocyte counts were extracted from the database, using most recent value up to 90 days prior to start of systemic therapy. Body mass index (BMI) at the approximate time of treatment initiation was calculated using custom R and SQL scripts. We omitted 21,023 patients without sufficient laboratory data for Khorana score calculation, resulting in 74,443 patients with Khorana scores.

**Table 2.** Khorana Score Calculation.

| Condition | Scoring |
| --- | --- |
| Cancer type (lung) | +1 |
| Platelet count $\geq 350 \times 10^9/L$ | +1 |
| Hemoglobin level $< 10 \text{ g/dL}$ | +1 |
| Leukocyte count $> 11 \times 10^9/L$ | +1 |
| BMI $\geq 35 \text{ kg/m}^2$ | +1 |

The Charlson comorbidity index score at the beginning of systemic cancer treatment was calculated using custom R and SQL scripts (Appendix 2), adapted from published methods (16-18). Diagnoses pertaining to the index categories and occurring before the start of cancer therapy were extracted from the database and scored according to the index algorithm (Table 3). We omitted 187 patients without sufficient data for Charlson score calculation, resulting in 95,279 patients with Charlson scores.

**Table 3.** Charlson Score Calculation.

| Condition | Scoring |
| --- | --- |
| Myocardial infarction | +1 |
| Congestive heart failure | +1 |
| Peripheral vascular disease | +1 |
| Cerebrovascular disease | +1 |
| Dementia | +1 |
| Chronic pulmonary disease | +1 |
| Rheumatologic disease | +1 |
| Peptic ulcer disease | +1 |
| Mild liver disease | +1 |
| Diabetes without chronic complication | +1 |
| Diabetes with chronic complication | +2 |
| Hemiplegia or paraplegia | +2 |
| Renal disease | +2 |
| Any malignancy, including leukemia and lymphoma | +2 |
| Moderate or severe liver disease | +3 |
| Metastatic solid tumor | +6 |
| AIDS/HIV | +6 |
| Age $\geq 80$ | +4 |
| Age 70-79 | +3 |
| Age 60-69 | +2 |
| Age 50-59 | +1 |

We also examined the presence and effects of prophylactic anticoagulation and prophylactic aspirin. Anticoagulant drugs studied were warfarin, low molecular weight heparin, enoxaparin, dalteparin, apixaban, rivaroxaban, dabigatran, and edoxaban. A patient was considered to be on prophylactic anticoagulation or aspirin if they had a prescription filled within a window of 5 days prior to 30 days after start of systemic therapy, so long as the prescription preceded (by at least one day) any diagnosed TEE while on treatment. All drugs were identified in the database by name using RxNorm (14).

Kaplan Meier survival analyses were performed using the survminer package (version 0.4.8; 19) for R (version 4.0.2; 20) and custom R and SQL scripts (Appendix 2). Risk of TEE was modeled using Cox proportional hazard regression with and without interactions including the following features: method of treatment (chemotherapy, immunotherapy, combined chemo- and immunotherapies, or non-chemo non-immunotherapy control), Khorana score, Charlson score, prior TEE, prophylactic anticoagulation or aspirin use, the time between initial cancer diagnosis and treatment, and patient’s age at treatment. Further analyses were performed using the cmprsk (version 2.2.10; 21) and aod (version 1.3.1; 22) packages, and the ggplot2 (version 3.3.2; 23) package was used for generating graphs. Cumulative probabilities of TEE and death during the first 180 days of systemic therapy were calculated for each treatment group using a competing risks framework. Fine-Gray subdistribution hazard ratios were computed for the covariates (24,25). We censored data according to date of last recorded clinical follow-up or date of death, whichever occurred first. We used the Strengthening the Reporting of Observational Studies in Epidemiology (STROBE) Checklist for cohort studies to ensure the comprehensiveness of this report (26, Appendix 3). All p-values below 0.001 were reported as “p < 0.001.”

## Results

We first studied the incidence of TEE in our cohort, stratified by type of systemic therapy (chemotherapy, immunotherapy, combined chemo- and immunotherapies, or non-chemo non-immunotherapy control). Note that the majority of events observed were pulmonary embolus (52.0%), followed by deep vein thrombosis (20.4%), cerebral infarction (18.7%), and acute myocardial infarction (6.79%); the remaining events were of multiple types. We identified an overall TEE rate of 6.8% (n = 6,493) within the first 180 days of treatment initiation, with a median time to TEE of 49 days (range 1 – 180). Because the majority (88.0%) of our cohort received only chemotherapy, the TEE rate and median time to TEE were similar in the chemotherapy alone group compared to the cohort overall. A significantly higher proportion of patients receiving immunotherapy alone (9.09%, n = 234, p < 0.02) experienced a TEE with a median time to event of 32.5 days (range 1 – 175 days), and a significantly lower proportion of patients receiving the non-chemo non-immunotherapy control agents (3.68%, n = 205, p < 0.001) experienced a TEE with a median time to event of 26 days (range 1 – 178 days). Results for all groups are summarized in Table 4. Using Kaplan-Meier cumulative risk analysis, we noted that patients who received immunotherapy alone were not only most likely to experience a TEE, but that TEE in this group occurred sooner after the start of treatment when compared to patients receiving chemotherapy alone, combined chemo- and immunotherapies, or neither of these therapies (Figure 1). These results were unchanged when using a competing risk analysis accounting for an observed death rate of 26.5% by 180 days (Supplementary Figure 1).

**Table 4.** Incidence of and Time to TEE Stratified by Treatment Type. The incidence of TEE was lowest in the Control group and highest in the Immunotherapy group. The Chemotherapy and Combined groups did not differ significantly from each other, but each did differ from the other groups. The median time to TEE was significantly shorter in the Control and Immunotherapy groups than in the Chemotherapy and Combined groups.

|  | **Chemotherapy**<br>**alone**<br>**n = 83,981** | **Immunotherapy**<br>**alone**<br>**n = 2,574** | **Combined**<br>**therapies**<br>**n = 3,347** | **Control**<br>**n = 5,564** | **p-value** |
| --- | --- | --- | --- | --- | --- |
| **TEE** | 5804 (6.91%) | 234 (9.09%) | 250 (7.47%) | 205 (3.68%) | p < 0.024<sup>a</sup> |
| **Time to event**<br>**median ± MAD** | 49 ± 51.9 | 32.5 ± 40.8 | 58.5 ± 66.0 | 26 ± 32.6 | p < 0.001<sup>b</sup> |

<sup>a</sup> Chemotherapy and Combined groups did not differ from each other
<sup>b</sup> Immunotherapy and Control groups did not differ from each other; Chemotherapy and Combined groups did not differ from each other

**Figure 1.**
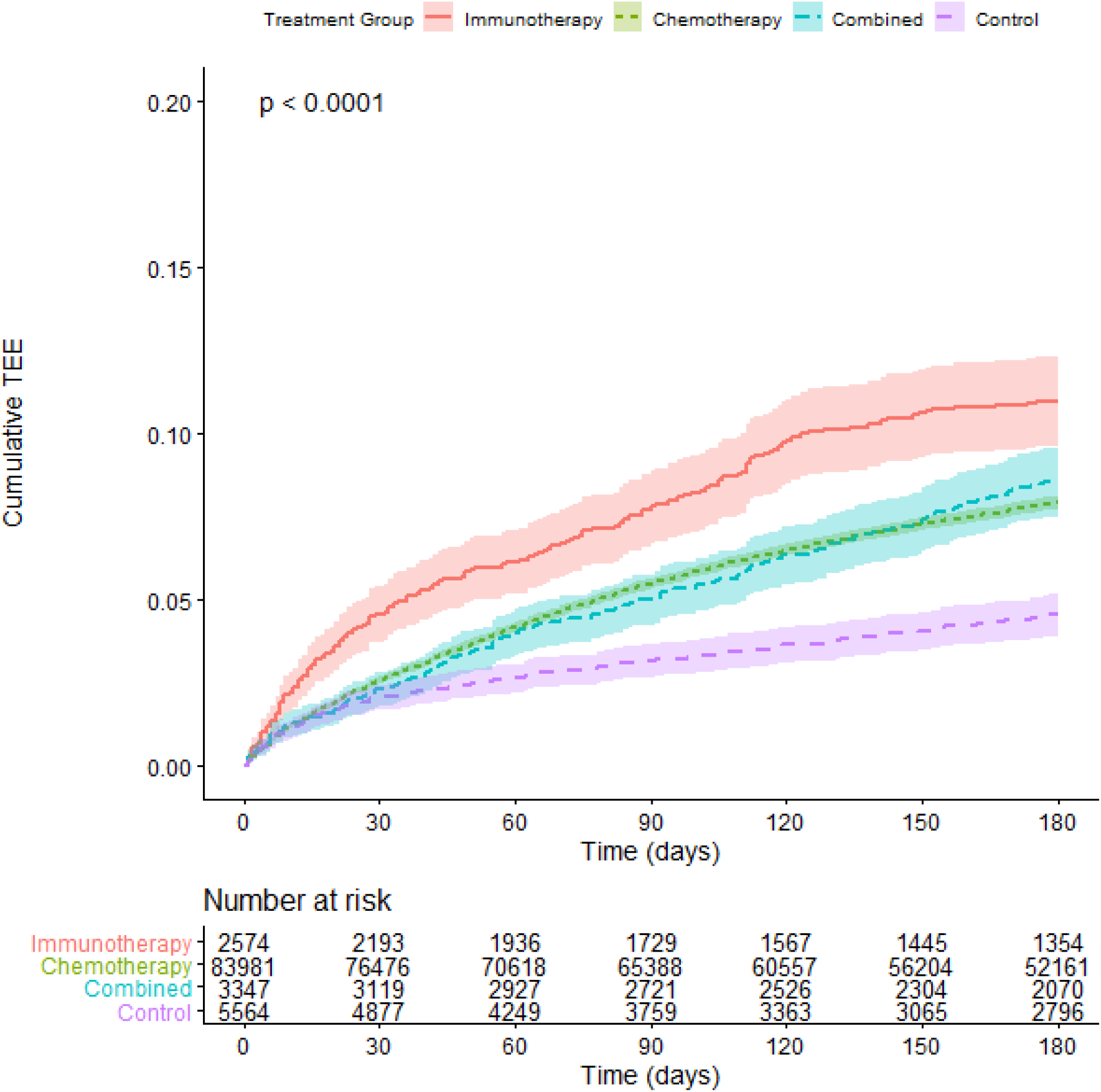
Cumulative Risk of Thrombosis in Lung Cancer Patients Stratified by Treatment Type. Risk of experiencing a TEE during the first 180 days after the start of treatment for lung cancer patients. The cohort is stratified by treatment group: immunotherapy alone (red), chemotherapy alone (green), or combination of both chemo- and immunotherapies (blue), as well as a non-chemo non-immunotherapy control group (purple). Time (in days from start of treatment) is shown along the x-axis, with cumulative risk of TEE along the y-axis. The lower table depicts number of individuals at risk for each 30-day interval. P-value reflects a significant difference in cumulative risk of TEE for the immunotherapy v. chemotherapy (alone or combined) groups, the control v. chemotherapy (alone or combined) groups, and the immunotherapy v. control groups. P-value computed by log-rank.

Because platinum-based chemotherapy agents have been shown to be more thrombogenic than other chemotherapy agents (27-29), we subdivided the treatment group who received chemotherapy alone into a group that received at least one platinum-based agent (n=77,522) and a group that received no platinum-based agents during their cancer treatment (n=6,459). These two subgroups differed significantly by Wilcox test in time to but not incidence of TEE (Supplementary Table 1). However, the Kaplan-Meier plot modeling both time to and incidence together demonstrates the similarity of the risk of TEE in these two subgroups (Figure 2).

**Figure 2.**
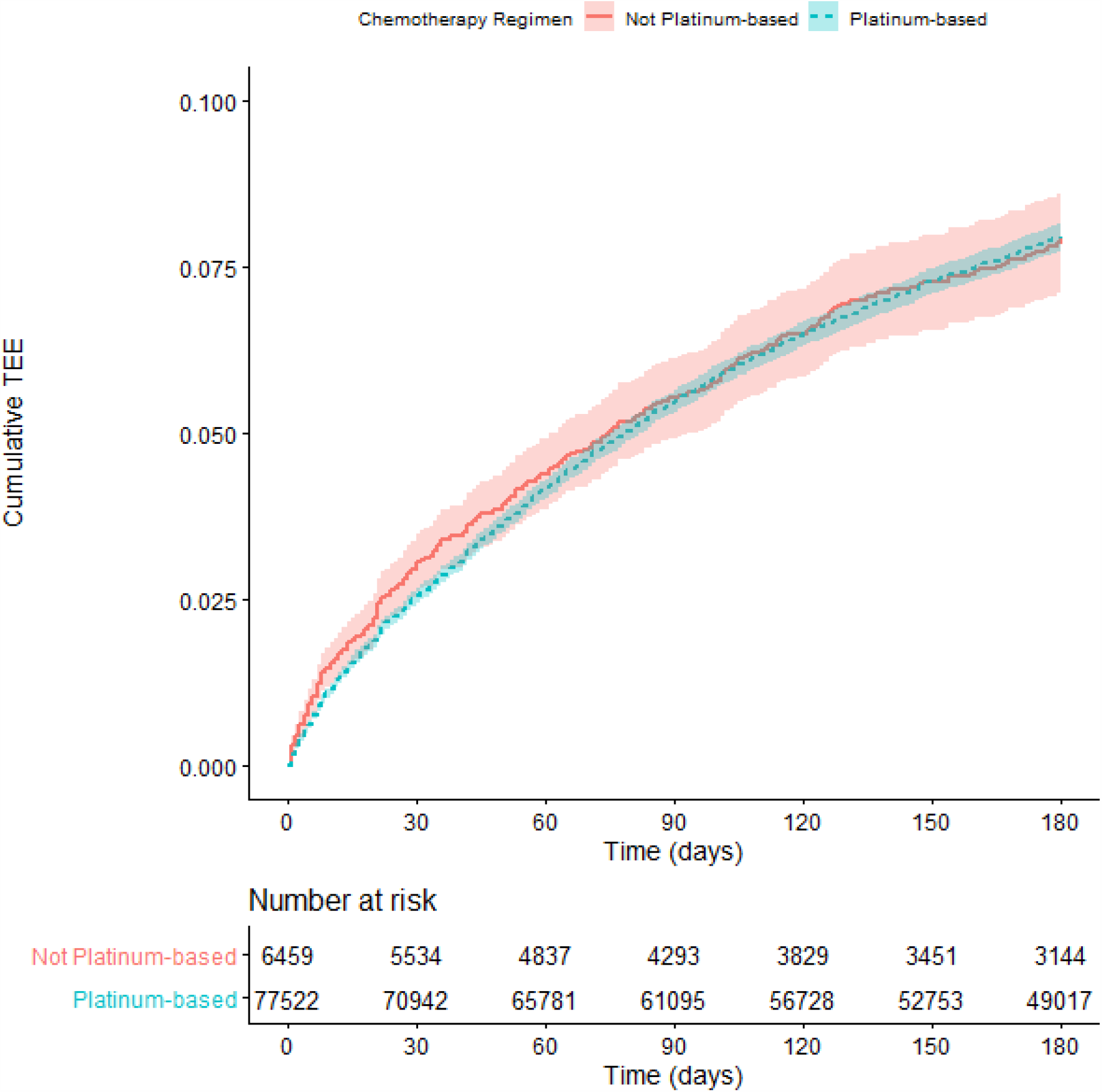
Cumulative Risk of Thrombosis in Lung Cancer Patients Stratified by Chemotherapy Regimen. Risk of experiencing a thromboembolic event during the first 180 days after the start of treatment for lung cancer patients. The cohort is stratified by chemotherapy regimen: not platinum-based agents (red) or platinum-based agents (blue). The two groups did not experience statistically significant different risks of events (p = 0.77). P-value computed by log-rank.

A prior history of TEE elevates a patient’s risk of experiencing a TEE during systemic therapy (30, 31) and may play a role in the decision to prescribe prophylactic anticoagulant or aspirin. We hypothesized that either or both of these factors could significantly affect observed TEE rates in our cohort and could be influencing the differences we observed among systemic treatment groups. We therefore analyzed TEE in our study cohort according to prior history of TEE and prophylactic anticoagulation or aspirin use, alone and in interaction. Surprisingly, we found that patients receiving prophylactic anticoagulation or aspirin experienced TEE at both a higher rate and a shorter interval than patients who did not (Supplementary Figure 2 and Supplementary Figure 3, respectively). In particular, the cohort receiving prophylactic anticoagulation had a much higher incidence of TEE compared to those receiving aspirin. While additional undetermined clinical risk factors could be significantly influencing patient TEE outcomes, we assessed the incidence of a prior history of TEE and its influence on subsequent TEE risk in this context and note that it accounts for the majority of increased TEE risk (Supplementary Figure 4). However, the use of prophylactic anticoagulation or aspirin does remain associated with a higher incidence of TEE (Figure 3 and Figure 4, respectively).

**Figure 3.**
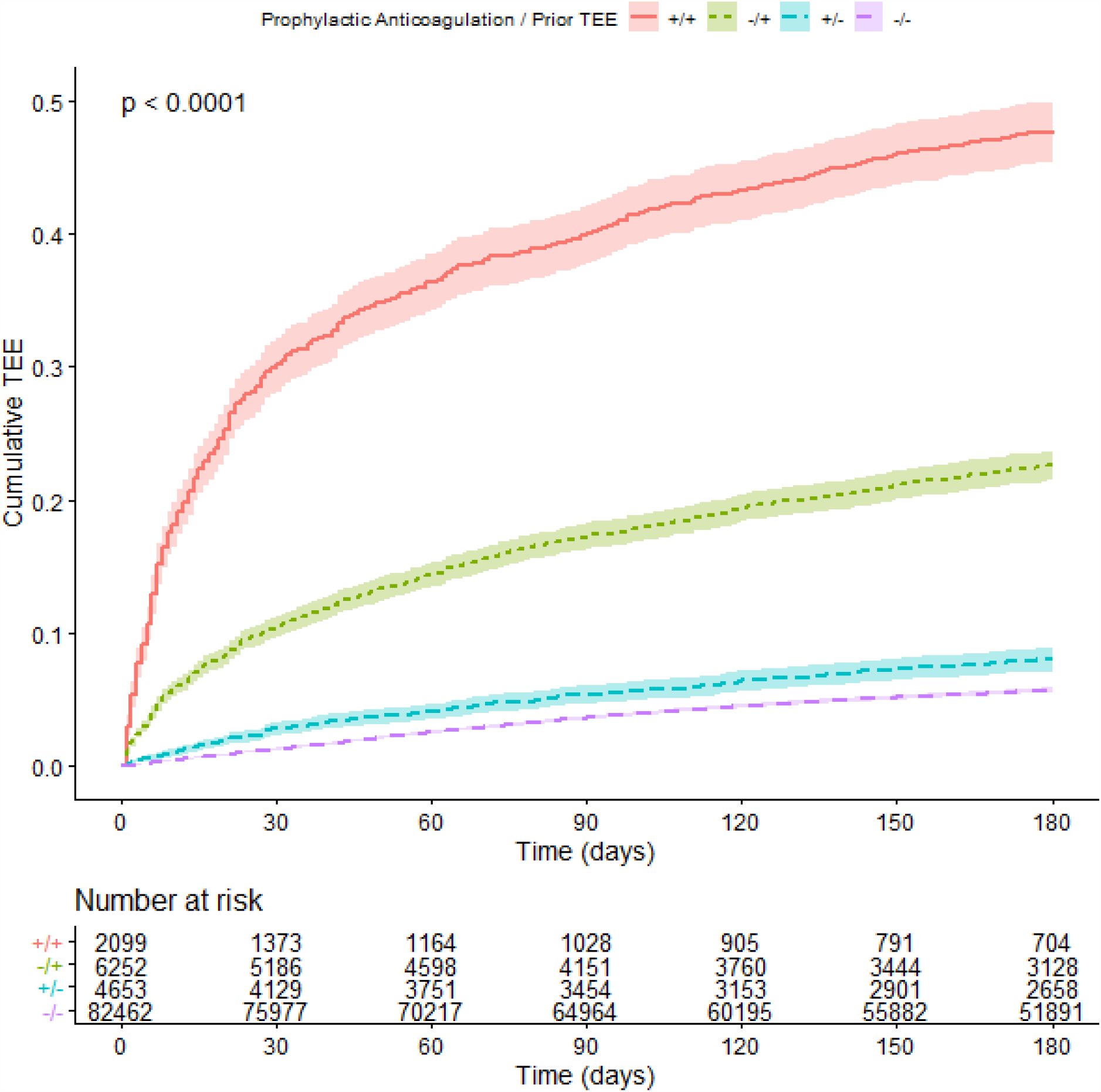
Cumulative Risk of Thrombosis in Lung Cancer Patients Stratified by Prior TEE and Prophylactic Anticoagulation Use. Risk of experiencing a thromboembolic event during the first 180 days after the start of treatment for lung cancer patients. The cohort is stratified by occurrence of prior TEE and prophylactic anticoagulation use: the presence of neither (red), the presence of a prior TEE and of anticoagulation use (purple), the presence of a prior TEE without anticoagulation use (green), or the use of prophylactic anticoagulation without a history of TEE (blue). Time (in days from start of treatment) is shown along the x-axis, with cumulative risk of TEE along the y-axis. The lower table depicts number of individuals at risk for each 30-day interval. P-value reflects significant differences in cumulative risk of TEE. P-value computed by log-rank.

**Figure 4.**
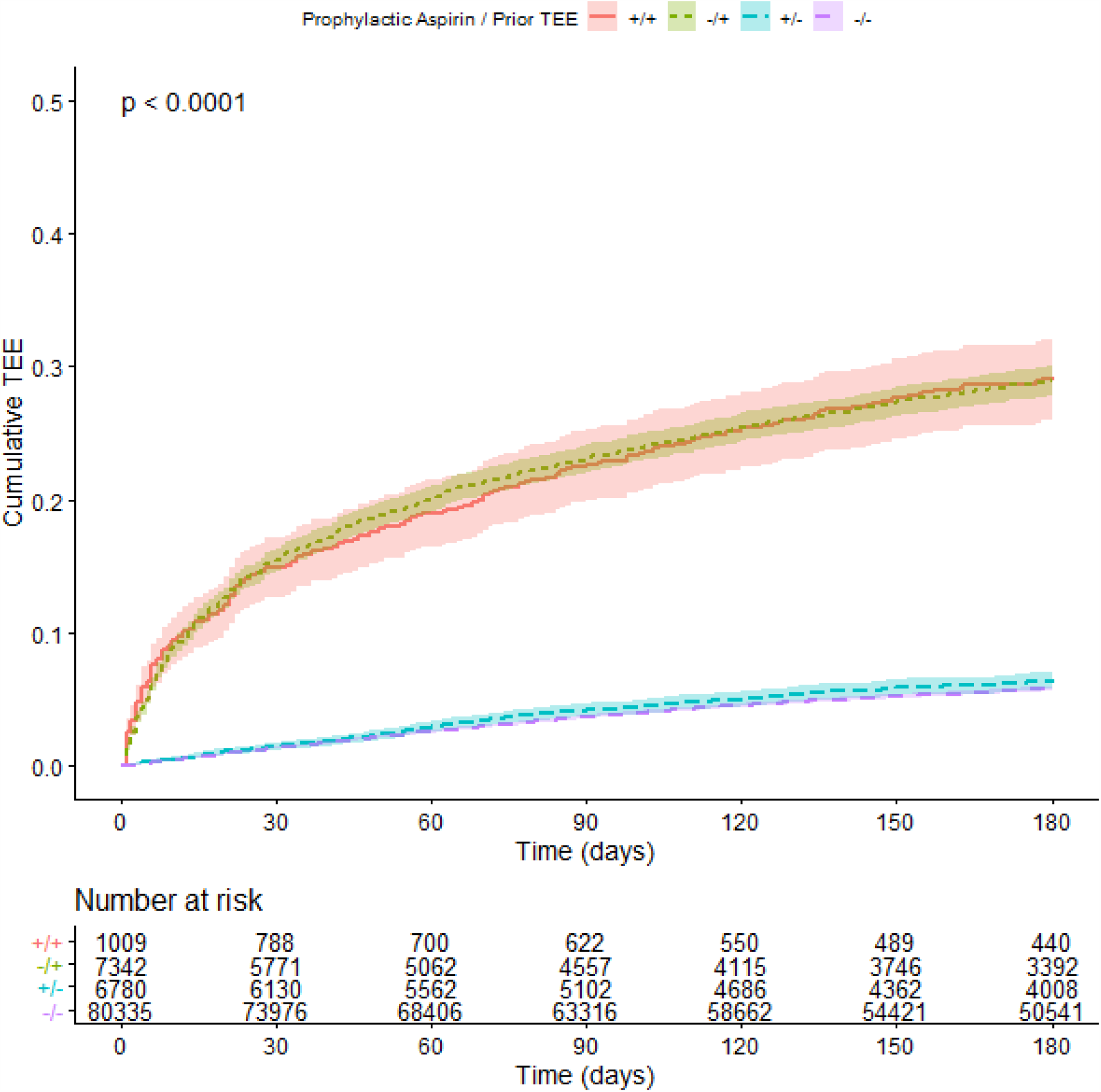
Cumulative Risk of Thrombosis in Lung Cancer Patients Stratified by Prior TEE and Aspirin Use. Risk of experiencing a thromboembolic event during the first 180 days after the start of treatment for lung cancer patients. The cohort is stratified by occurrence of prior TEE and prophylactic aspirin use: the use of aspirin without a history of TEE (blue), neither (red), the presence of a prior TEE and aspirin use (purple), or the presence of a prior TEE without aspirin use (green). Time (in days from start of treatment) is shown along the x-axis, with cumulative risk of TEE along the y-axis. The lower table depicts number of individuals at risk for each 30-day interval. P-value reflects significant differences in cumulative risk of TEE. P-value computed by log-rank.

We used the Cox proportional hazard model to quantify the effects of the different systemic treatments on patients’ risk of TEE (Figure 5). In the univariate model, patients treated with immunotherapy experienced 2.5 times greater risk of TEE compared to controls, while patients treated with chemotherapy experienced 1.7 times greater risk; whereas, combining chemo- and immunotherapies appeared to offset risk of TEE by 2.4-fold. To assess the relative importance of additional risk factors, we therefore created multivariate models describing the effects of the different systemic treatments combined with prophylactic anticoagulation or aspirin use, history of prior TEE, age, elapsed time between diagnosis and systemic treatment, Khorana score, and Charlson comorbidity score. When accounting for these covariates, the overall influence of treatment type was diminished, but the relative effects of the different treatment types remained the same (Figure 6). Unsurprisingly, as both Khorana and Charlson scores increase, so does the risk of TEE. Increased age and increased time between initial diagnosis and systemic treatment both demonstrated a slight protective effect, likely due to a more robust underlying health of patients who could either safely delay systemic treatment or be considered good candidates for systemic treatment even at an advanced age. Unsurprisingly, prior TEE was a strong predictor of subsequent TEE (hazard ratio of 4.04, p < 0.001). However, both prophylactic aspirin and anticoagulant use paradoxically increased the residual risk of TEE (hazard ratios of 1.10 and 1.38, respectively, p < 0.04), despite their known TEE-preventative effects; we suspect that this may arise from other latent clinical characteristics influencing prophylactic prescription rates.

**Figure 5.**
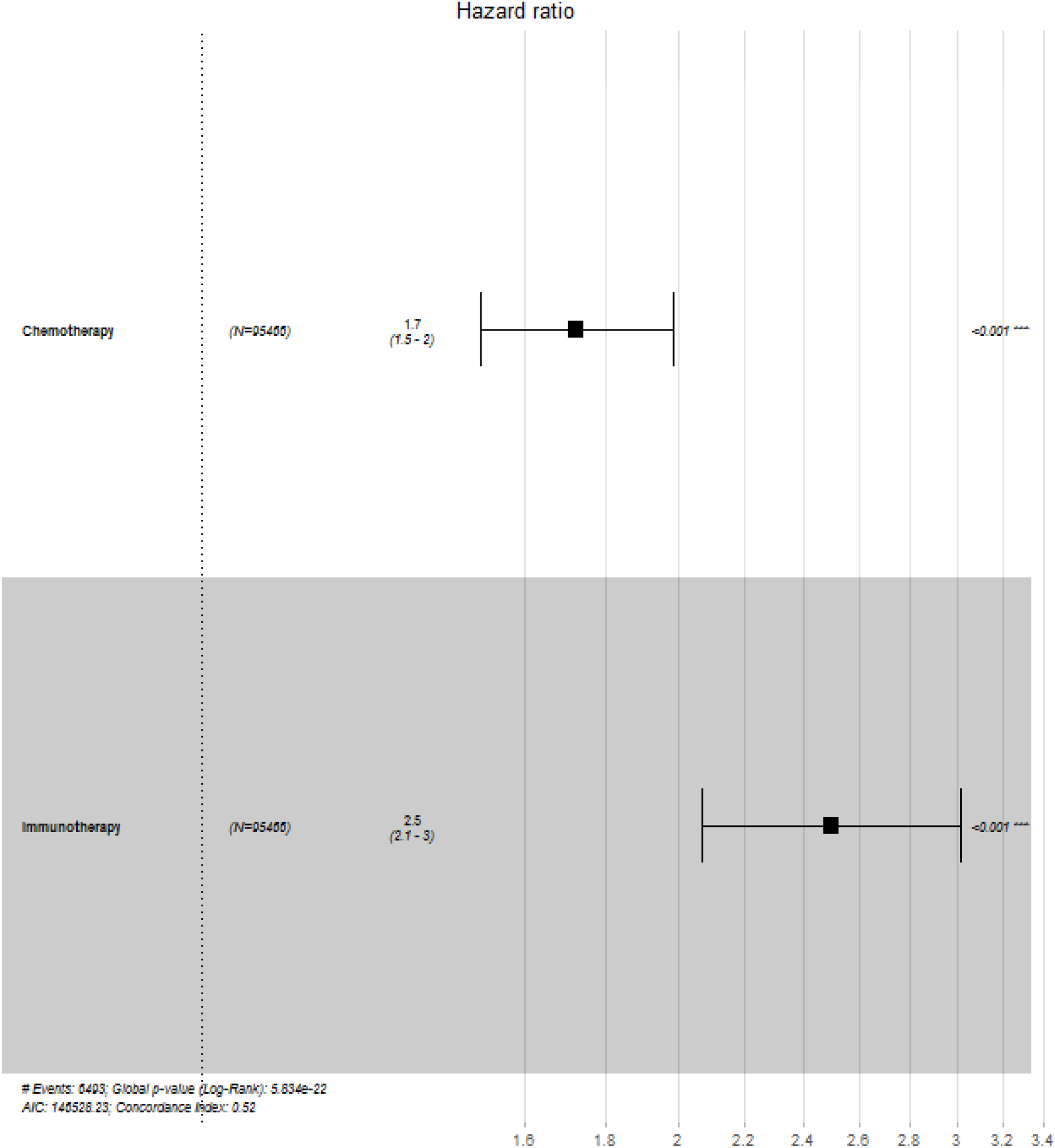
Cox Proportional Hazard Model of Effect on TEE of Treatment Type. Hazard ratios are represented on the x-axis, with a neutral value of 1 represented by the vertical dotted line and corresponding to the relative TEE risk among controls. The calculated value for hazard ratio and the 95% confidence interval are shown for both chemotherapy and immunotherapy. P-values for each treatment type are displayed on the right-hand side of the plot, and the global p-value and Akaike information criterion (AIC) estimate are in the lower left corner.

**Figure 6.**
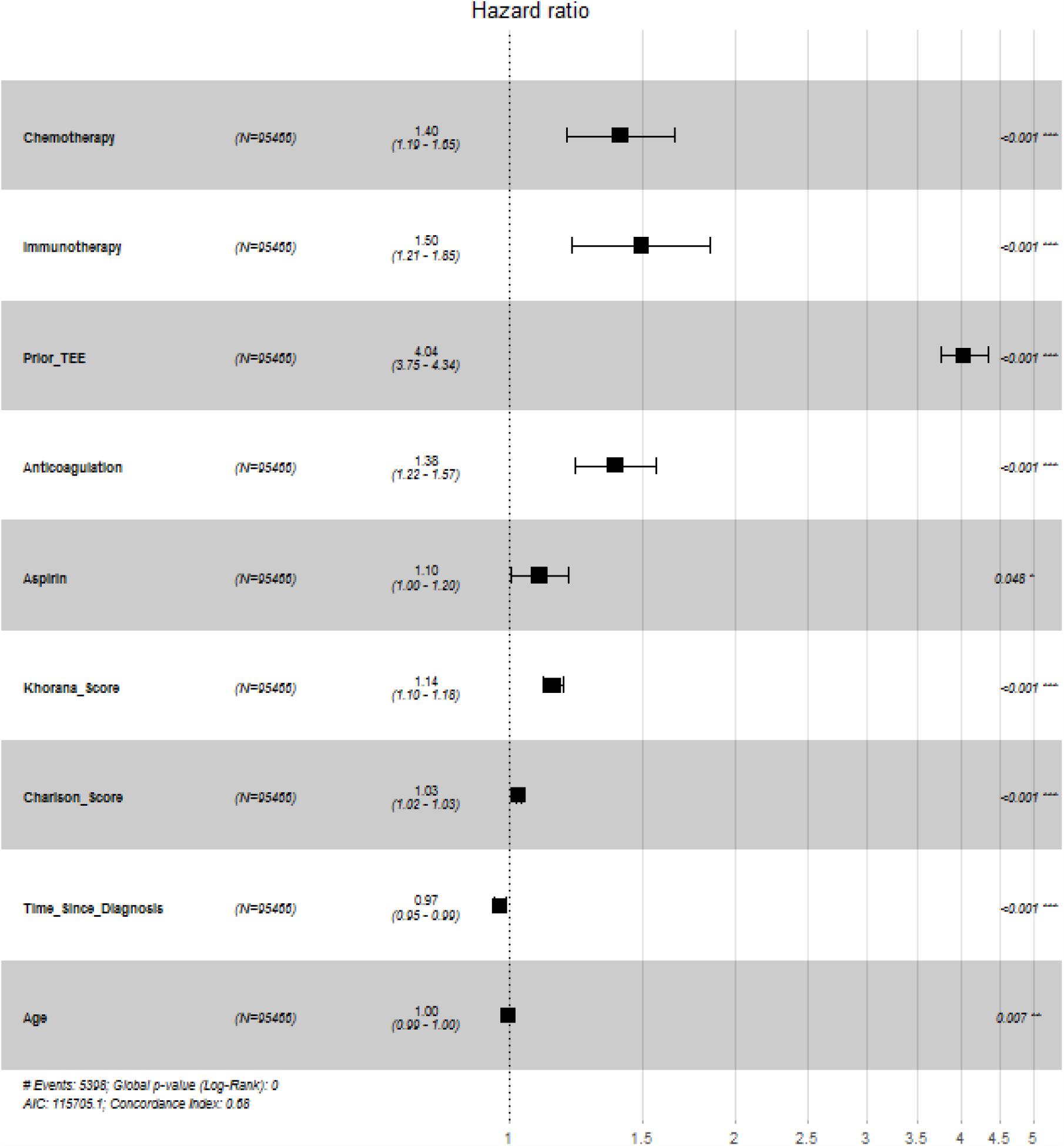
Cox Proportional Hazard Model of Effect on TEE of Treatment Type and Covariates. Hazard ratios are represented on the x-axis, with a neutral value of 1 represented by the vertical dotted line and corresponding to the relative TEE risk among controls. The calculated value for hazard ratio and the 95% confidence interval are shown for both chemotherapy and immunotherapy and all covariates. P-values for each covariate are displayed on the right-hand side of the plot, and the global p-value and Akaike information criterion (AIC) estimate are in the lower left corner.

We also used a competing risks model to determine Fine-Gray subdistribution hazards of TEE for the systemic treatments and covariates in consideration of the competing risk of death (Table 5). Treatment with chemotherapy demonstrated 1.2 times greater risk of TEE under this model, but the effect of treatment with immunotherapy no longer showed a significant effect; the hazard ratios of the covariates were similar to those developed in the Cox model.

**Table 5.** Fine-Gray Subdistribution Hazard Ratios for TEE Considering Competing Risk of Death. Using a competing risks model, systemic therapies for lung cancer demonstrated a significant increased risk of TEE. P-values were calculated by Wald test.

| **Variable** | **HR** | **95% CI** | **p-value** |
| --- | --- | --- | --- |
| **Chemotherapy** | 1.23 | 1.10 – 1.38 | p < 0.001 |
| **Immunotherapy** | 1.08 | 0.970 – 1.20 | p = 0.16 |
| **Prior TEE** | 4.79 | 4.50 – 5.09 | p = 0 |
| **Khorana Score** | 1.07 | 1.04 – 1.11 | p < 0.001 |
| **Charlson Score** | 1.02 | 1.01 – 1.02 | p < 0.001 |
| **Time Since Diagnosis** | 0.977 | 0.961 – 0.995 | p = 0.01 |
| **Age** | 0.993 | 0.990 – 0.997 | p < 0.001 |
| **Aspirin** | 1.07 | 0.972 – 1.17 | p = 0.18 |
| **Anticoagulation** | 2.12 | 1.98 – 2.27 | p = 0 |

Finally, in recognition of the strong effect on our study cohort of prior history of TEE on the risk of TEE during systemic treatment of lung cancer, we eliminated the 8,351 patients who had experienced a TEE before beginning lung cancer therapy, resulting in a reduced cohort of 87,115 Veterans. This change substantially reduced the cumulative risk of TEE experienced by the group who received immunotherapy alone; nonetheless, both chemotherapy and immunotherapy retained an increased risk of TEE (Figure 7).

**Figure 7.**
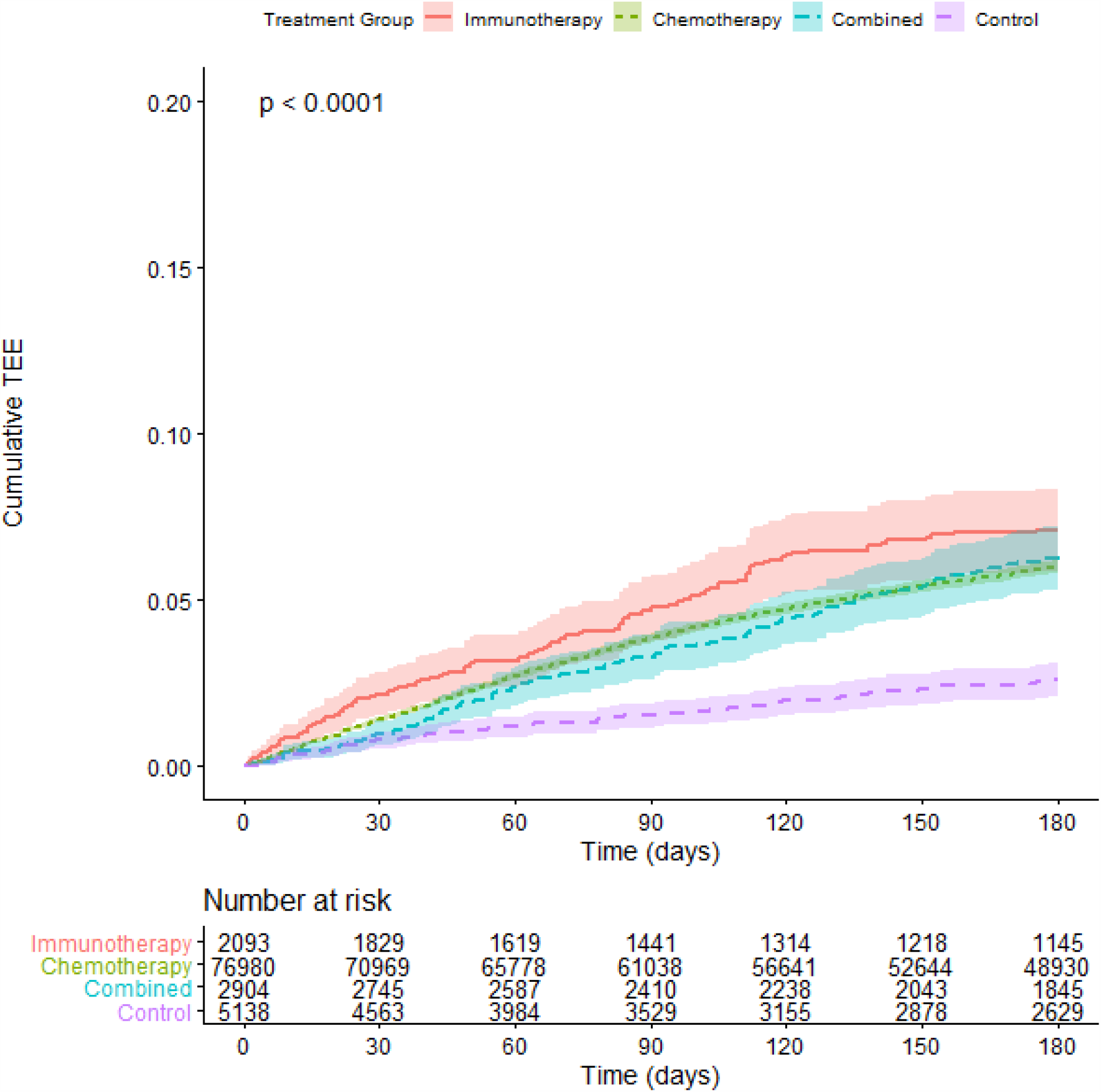
Cumulative Risk of Thrombosis in Lung Cancer Patients with no prior history of TEE, stratified by Treatment Type. Risk of experiencing a TEE during the first 180 days after the start of treatment for lung cancer patients. The cohort is stratified by treatment group: chemotherapy alone (green), immunotherapy alone (red), or combination of both chemotherapy and immunotherapies (blue), as well as a non-chemo non-immunotherapy control group (purple). Time (in days from start of treatment) is shown along the x-axis, with cumulative risk of TEE along the y-axis. The lower table depicts number of individuals at risk for each 30-day interval. P-value reflects a significant difference in cumulative risk of TEE for the immunotherapy v. chemotherapy (alone or combined) groups, the control v. chemotherapy (alone or combined) groups, and the immunotherapy v. control groups.

## Discussion

To the best of our knowledge, this is the largest study to-date exploring thromboembolic risk in the setting of lung cancer, and the first to report on these risks among a cohort of US Veterans. This is also the largest study to suggest a correlation in cancer patients between checkpoint inhibitor immunotherapy and increased thromboembolic risk. We surprisingly found that lung cancer patients on prophylactic anticoagulation or aspirin had a higher TEE incidence compared with those not on prophylaxis, and we speculate this counterintuitive finding may be due to latent clinical variables associated with a higher baseline thrombosis risk, in particular, but not wholly, the existence of a prior TEE.

Thromboembolic events have not been reported in phase III trials evaluating checkpoint inhibitors in patients with cancer, and little is known about thromboembolic risk in this setting. In one secondary analysis within a prospective observational protocol of 217 NSCLC patients treated with checkpoint inhibitors, an elevated thrombosis incidence of 13.8% was noted (32). Other small, single institutional studies have also observed a correlation between checkpoint inhibitor therapy and thrombosis in cancer patients (33, 34). In our study, we found a potential increased incidence of thromboembolic events among lung cancer patients who received first line checkpoint inhibitor monotherapy compared with chemotherapy either alone or combined with immunotherapy. These data likely merit further clinical and translational studies to investigate the potential thrombogenicity of checkpoint inhibitors in the lung cancer setting (whether alone or in combination with chemotherapy) and argue for incorporating this therapeutic class into thrombosis risk models and recommendations for prophylactic anticoagulation.

In the present study, we also found evidence of a weak correlation between thrombosis risk and the Khorana score (a widely utilized VTE risk model in cancer patients). However, we note that other studies have been unable to demonstrate a significant association between the Khorana score and VTE in lung cancer patients (35-37). These data call into question the routine clinical utility of the Khorana Score in the lung cancer setting.

We note differences in the overall observed TEE rates in our lung cancer cohort treated with systemic therapy compared to the published literature. For instance, a retrospective analysis of 204 lung cancer patients treated with cisplatin-based chemotherapy, found an 11.8% incidence of TEE during treatment or within 4 weeks of the last treatment (27). While this incidence is higher than the cumulative incidence we observe (6.80%), we note this discrepancy could be explained by different length of observation between the two studies (our study only investigated events occurring within 180 days of treatment initiation) and the small cohort size in the prior study.

There are limitations associated with administrative databases, including numerous sources of possible bias and missing or inaccurate data. Additionally, while the cohort we analyzed was national in scope, it is most reflective of the US Veteran population, and therefore contains relatively few women. Moreover, we did not explicitly study the influence of initial disease staging, potentially relevant laboratory results such as D-dimer, family history of VTE, or non-prescription medications. We only analyzed the impact of clinical predictors at the time of treatment initiation and did not assess risk factors that may have arisen later in the treatment course. Finally, we confined analysis to lung cancers, as they are known to be at increased risk for TEEs compared to most other malignancies (15) and did not discriminate between lung cancer subtypes (e.g. small cell).

In conclusion, given the continued high rate of TEEs in lung cancer and the increased risks associated with chemotherapy and immunotherapy, future thromboprophylaxis risk-adapted trials in this setting may be warranted, particularly accounting for immunotherapy-based treatment regimens.

## Data Availability

Patient-level data is available only to qualified VA researchers.

## Acknowledgements

This work was supported by VA Career Development Award (1 IK2 CX002049-01) to RFT.

All authors made substantial contributions to the research and approved the final version of the manuscript. CJM, RFT, and DCC designed the research question. CJM performed the research. CJM, MJC, KRG, and RFT created the SQL and R programming tools used to extract and analyze the data. CJM, RAM, KRG, and RFT analyzed the data. CJM, RAM, RFT, and DCC wrote the paper.

This product uses publicly available data courtesy of the U.S. National Library of Medicine (NLM), National Institutes of Health, Department of Health and Human Services; NLM is not responsible for the product and does not endorse or recommend this or any other product.

## Appendix 1 Diagnostic criteria for “lung cancer” and “TEE”

**Table 1.** Diagnostic Inclusion Criteria. Patients included in the study experienced at least one of the lung cancer diagnoses listed in this table.

| System | Code | Description |
| --- | --- | --- |
| ICD-9 | 162.2 | Malignant neoplasm of main bronchus |
| ICD-9 | 162.3 | Malignant neoplasm of upper lobe, bronchus, or lung |
| ICD-9 | 162.4 | Malignant neoplasm of middle lobe, bronchus, or lung |
| ICD-9 | 162.5 | Malignant neoplasm of lower lobe, bronchus, or lung |
| ICD-9 | 162.8 | Malignant neoplasm of other parts of bronchus or lung |
| ICD-9 | 162.9 | Malignant neoplasm of bronchus and lung, unspecified |
| ICD-10-CM | C34.00 | Malignant neoplasm of unspecified main bronchus |
| ICD-10-CM | C34.01 | Malignant neoplasm of right main bronchus |
| ICD-10-CM | C34.02 | Malignant neoplasm of left main bronchus |
| ICD-10-CM | C34.10 | Malignant neoplasm of upper lobe, unspecified bronchus, or lung |
| ICD-10-CM | C34.11 | Malignant neoplasm of upper lobe, right bronchus, or lung |
| ICD-10-CM | C34.12 | Malignant neoplasm of upper lobe, left bronchus, or lung |
| ICD-10-CM | C34.30 | Malignant neoplasm of lower lobe, unspecified bronchus, or lung |
| ICD-10-CM | C34.31 | Malignant neoplasm of lower lobe, right bronchus, or lung |
| ICD-10-CM | C34.32 | Malignant neoplasm of lower lobe, left bronchus, or lung |
| ICD-10-CM | C34.80 | Malignant neoplasm of overlapping sites of unspecified bronchus and lung |
| ICD-10-CM | C34.81 | Malignant neoplasm of overlapping sites of right bronchus and lung |
| ICD-10-CM | C34.82 | Malignant neoplasm of overlapping sites of left bronchus and lung |
| ICD-10-CM | C34.90 | Malignant neoplasm of unspecified part of unspecified bronchus or lung |
| ICD-10-CM | C34.91 | Malignant neoplasm of unspecified part of right bronchus or lung |
| ICD-10-CM | C34.92 | Malignant neoplasm of unspecified part of left bronchus or lung |

**Table 2.** TEE definitions. This table lists the specific diagnostic codes used to define TEE.

| **System** | **Code** | **Description** |
| --- | --- | --- |
| ICD-9 | 410.90 | Acute myocardial infarction, unspecified site, episode of care unspecified |
| ICD-9 | 415.12 | Septic pulmonary embolism |
| ICD-9 | 415.13 | Saddle embolus of pulmonary artery |
| ICD-9 | 415.19 | Other pulmonary embolism and infarction |
| ICD-9 | 434.91 | Cerebral artery occlusion, unspecified, with cerebral infarction |
| ICD-9 | 453.40 | Acute venous embolism and thrombosis of unspecified deep vessels of lower extremity |
| ICD-9 | 453.82 | Acute venous embolism and thrombosis of deep veins of upper extremity |
| ICD-10-CM | I21.9 | Acute myocardial infarction, unspecified |
| ICD-10-CM | I26.01 | Septic pulmonary embolism with acute cor pulmonale |
| ICD-10-CM | I26.02 | Saddle embolus of pulmonary artery with acute cor pulmonale |
| ICD-10-CM | I26.09 | Other pulmonary embolism with acute cor pulmonale (includes NOS) |
| ICD-10-CM | I26.90 | Septic pulmonary embolism without acute cor pulmonale |
| ICD-10-CM | I26.92 | Saddle embolus of pulmonary artery without acute cor pulmonale |
| ICD-10-CM | I26.93 | Single subsegmental pulmonary embolism without acute cor pulmonale |
| ICD-10-CM | I26.94 | Multiple subsegmental pulmonary emboli without acute cor pulmonale |
| ICD-10-CM | I26.99 | Other pulmonary embolism without acute cor pulmonale |
| ICD-10-CM | I63.9 | Cerebral infarction, unspecified |
| ICD-10-CM | I82.401 | Acute embolism and thrombosis of unspecified deep veins of right lower extremity |
| ICD-10-CM | I82.402 | Acute embolism and thrombosis of unspecified deep veins of left lower extremity |
| ICD-10-CM | I82.403 | Acute embolism and thrombosis of unspecified deep veins of lower extremity, bilateral |
| ICD-10-CM | I82.409 | Acute embolism and thrombosis of unspecified deep veins of unspecified lower extremity |
| ICD-10-CM | I82.601 | Acute embolism and thrombosis of unspecified deep veins of right upper extremity |
| ICD-10-CM | I82.602 | Acute embolism and thrombosis of unspecified deep veins of left upper extremity |
| ICD-10-CM | I82.603 | Acute embolism and thrombosis of unspecified deep veins of upper extremity, bilateral |
| ICD-10-CM | I82.609 | Acute embolism and thrombosis of unspecified deep veins of unspecified upper extremity |

## Appendix 2 SQL and R Statements

*Unredacted, complete statements can be made available to qualified VA VINCI researchers*

I. Data Aggregation
  a. Identify patients with specified cancer diagnoses.
  b. Identify patients with specified treatment agents.
  c. Form cohort from intersection of patients with specified diagnoses and treatments.
  d. Retrieve relevant clinical and demographic data for cohort.
    i. Retrieve demographic data (sex, DOB, DOD, date of last follow up).
    ii. Retrieve anticoagulation data.
    iii. Calculate BMI.
    iv. Retrieve lab information (platelets, hemoglobin, leukocytes).
    v. Calculate Khorana score.
    vi. Calculate Charlson score.
  e. Identify TEEs.
II. Data Analysis
  a. Develop descriptive statistics of cohort.
  b. Perform Kaplan-Meier analysis.
  c. Perform Cox Proportional Hazard analysis.
  d. Perform competing risk and subdistribution hazard analyses.

### Data Aggregation

#### Identify patients with specified cancer diagnoses

A custom R function was used to connect with both the CDW and the research database and perform the following steps. The relevant dimension tables of the data warehouse were queried to determine the identifiers associated with ICD codes for the diagnoses listed in Appendix 1. All relevant views of the research database were queried for these identifiers to identify all instances of diagnoses. The resultant dataset comprised all unique instances of the specified diagnoses and associated dates, aggregated to the highest level (location-agnostic) of unique patient identifier. From this dataset, a table was created in the research database containing patient identifiers and the earliest associated date of diagnosis (one row per individual patient).

#### Identify patients with specified treatment agents

This portion of the work was performed in SQL. RxNorm was used to ensure all variants of agent names, as listed in the main body of the paper, were included. The names were used to determine the identifiers associated with the specified treatment agents in multiple dimension tables of the data warehouse: CPT codes, IV usage, EHR orderable items, and drugs. All relevant views of the research database were queried for these identifiers to identify all instances of an association between a patient and a treatment agent. The resultant dataset comprised all unique instances of the specified treatment agents and associated dates, aggregated to the highest level (location-agnostic) of unique patient identifier. From this dataset, a table was created in the research database containing patient identifiers and the earliest associated date of treatment with each agent (one row per combination of patient and agent).

Example:

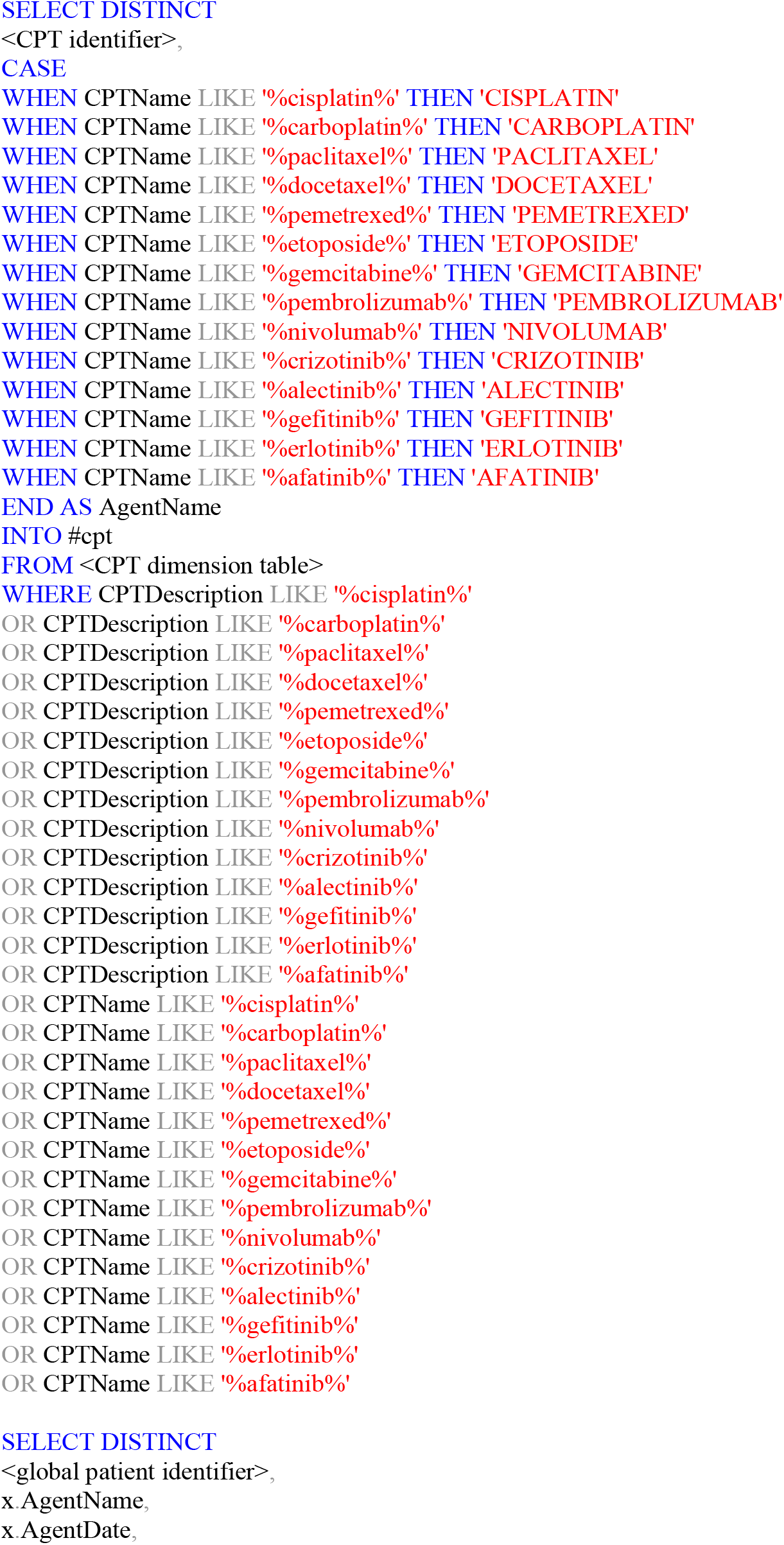

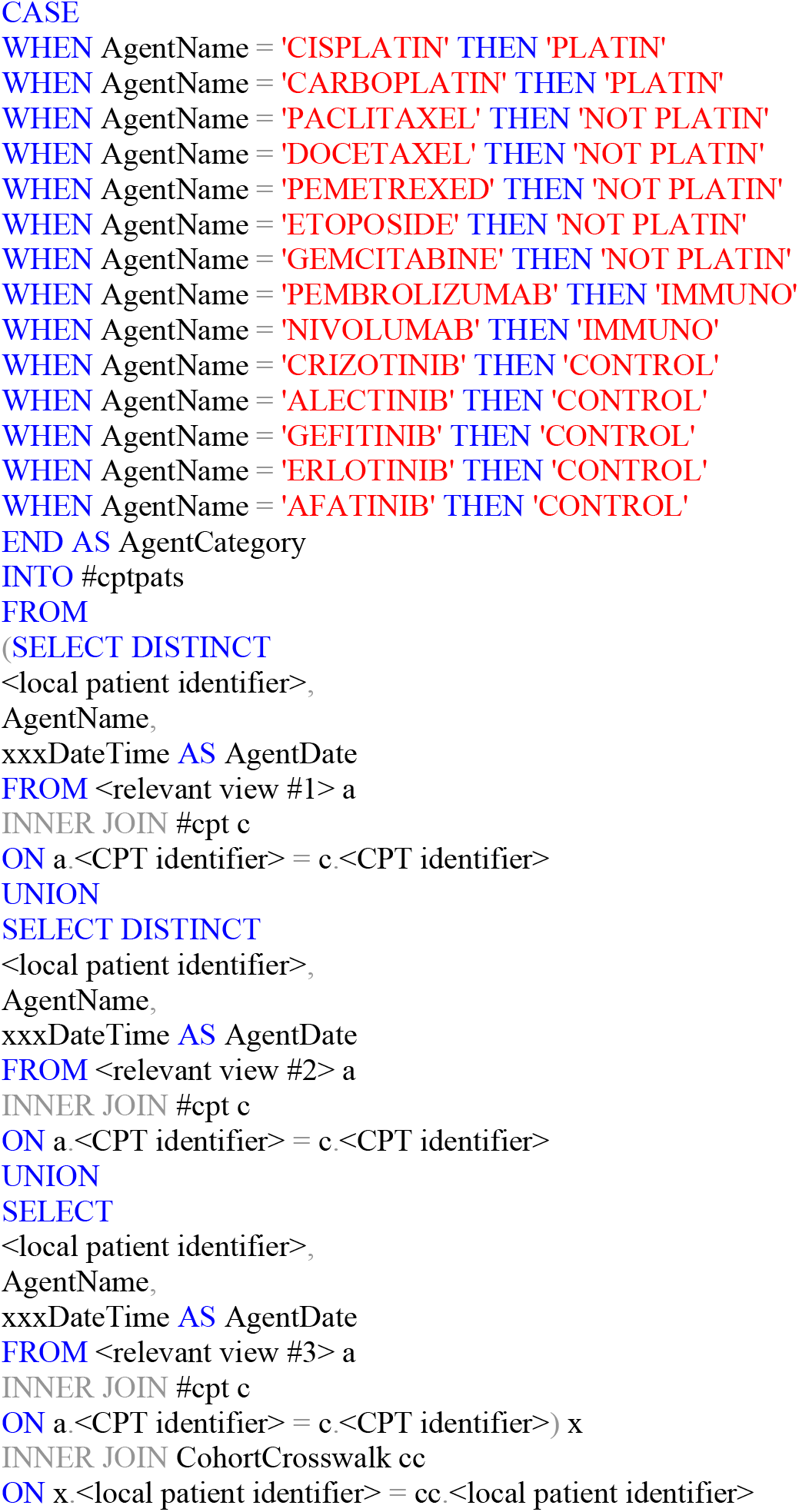

#### Form cohort from intersection of patients with specified diagnoses and treatments

This portion of the work was performed in SQL. The study cohort comprised the patients represented by rows in both the created diagnosis and treatment tables, with the restriction that treatment must be both after diagnosis and within 180 days of earliest treatment (for patients treated with multiple agents).

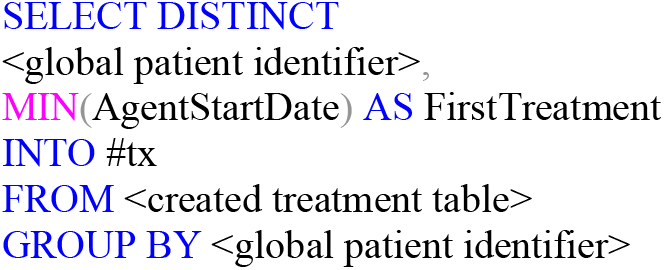

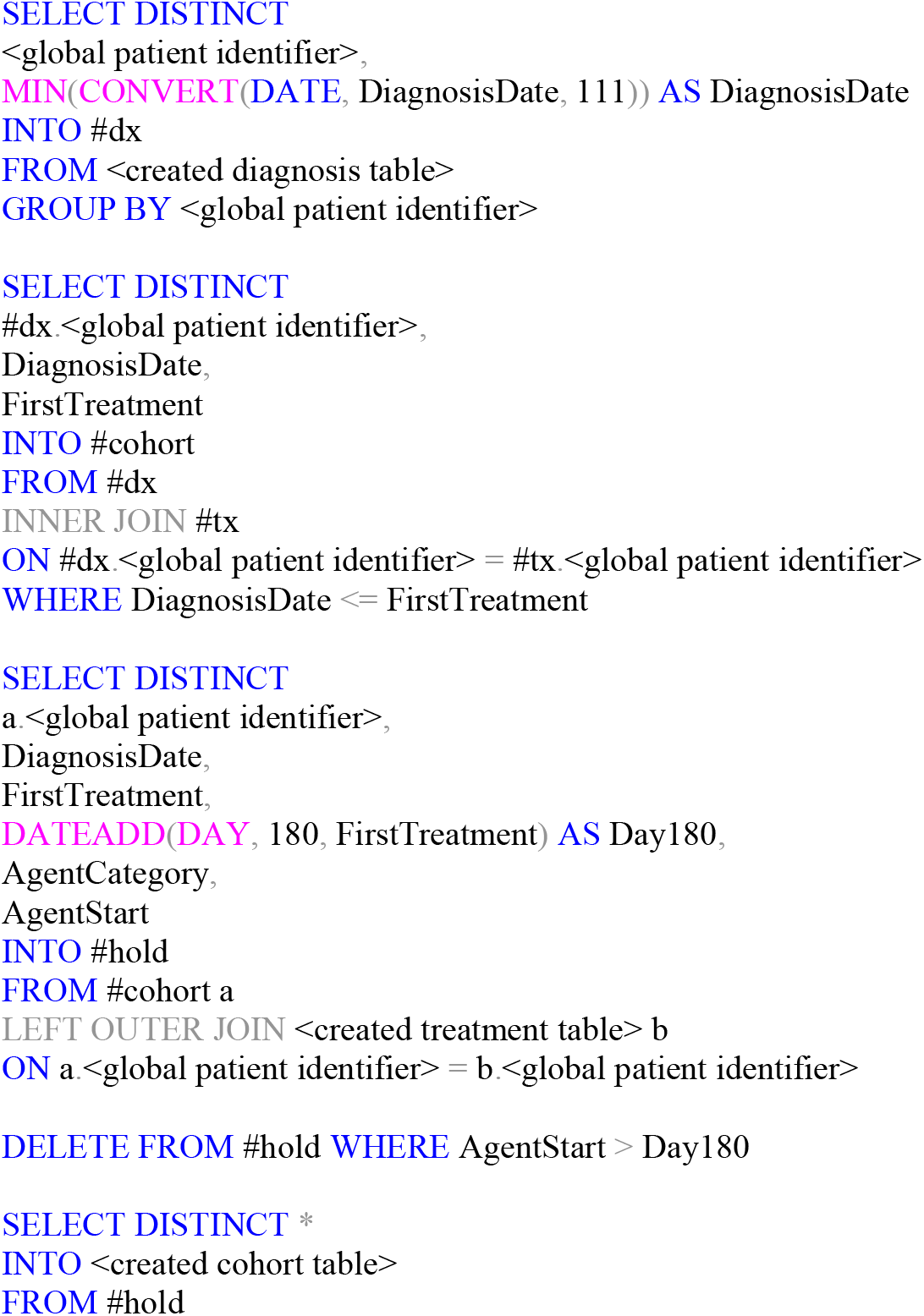

#### Retrieve relevant clinical and demographic data for cohort

##### Retrieve demographic data (sex, DOB, DOD, date of last follow up)

This portion of the work was performed in SQL. Sex and dates of birth, death, and last follow up were retrieved for each member of the cohort from the relevant views in the research database.

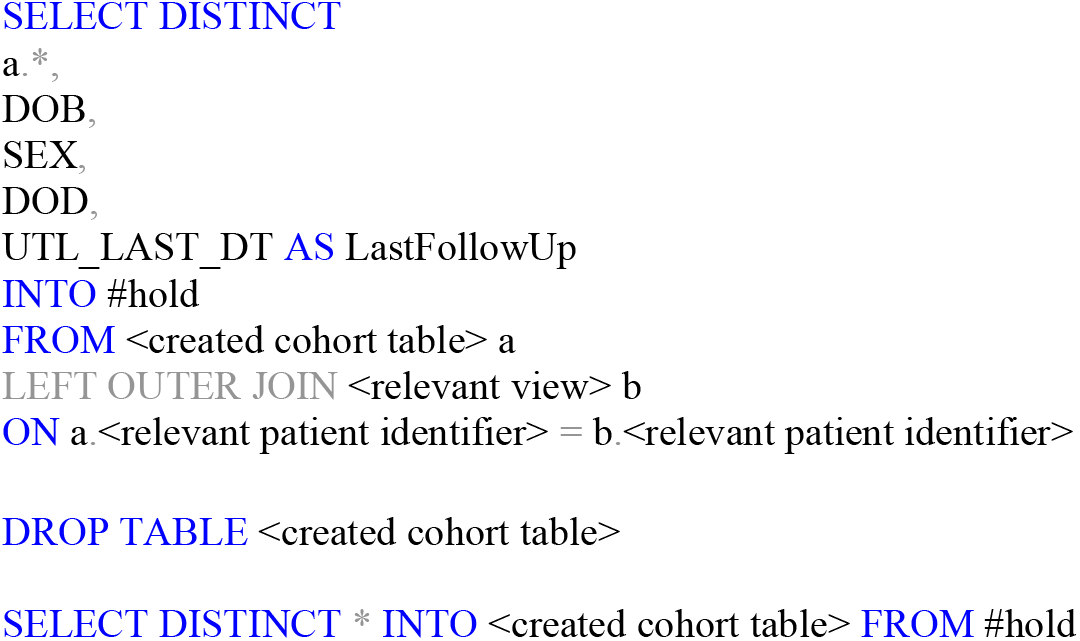

##### Retrieve anticoagulation data

This portion of the work was performed in SQL. Anticoagulants, as listed in the main body of the paper, were identified by name and retrieved using the same methods as described for the treatment agents.

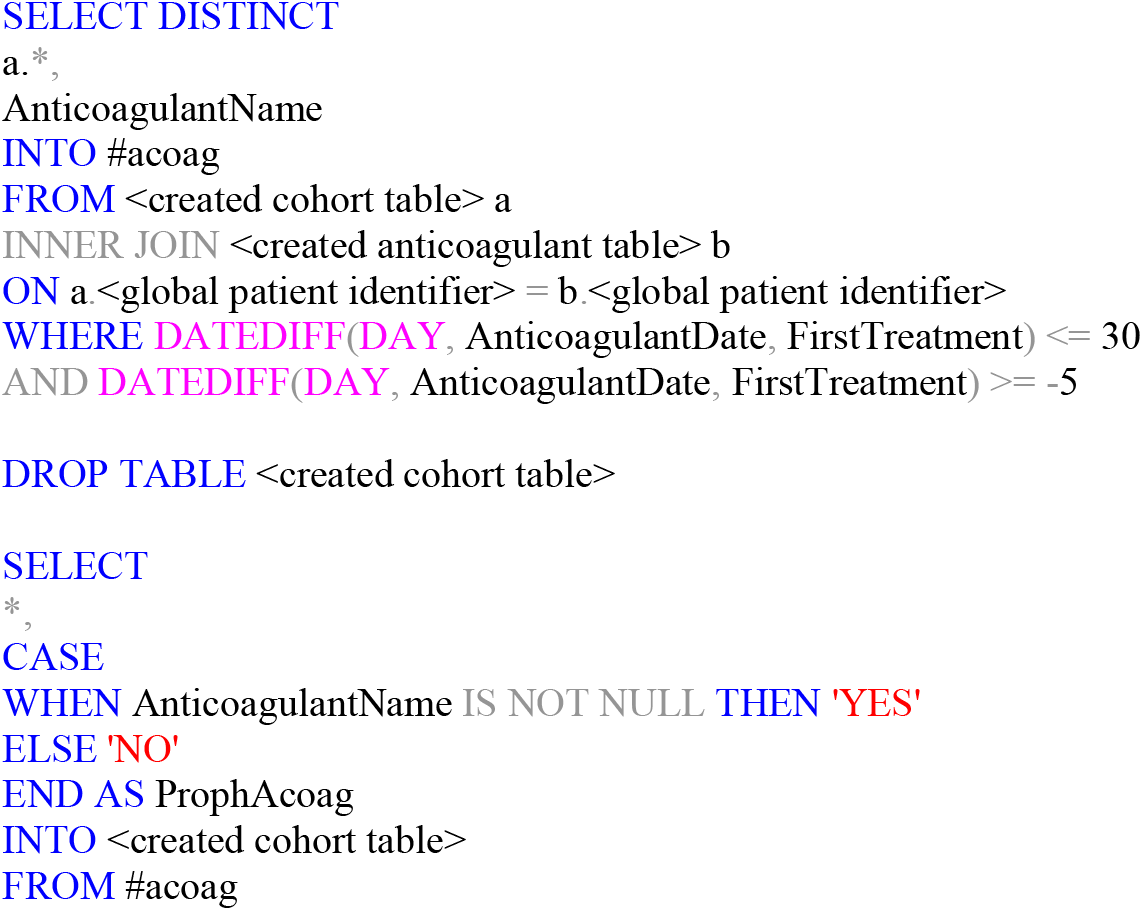

##### Calculate BMI

A custom R function was used to determine each patient’s BMI at the time of treatment start.

##### Retrieve lab information (platelets, hemoglobin, leukocytes)

This portion of the work was performed in SQL. LOINC codes were used to identify relevant lab tests.

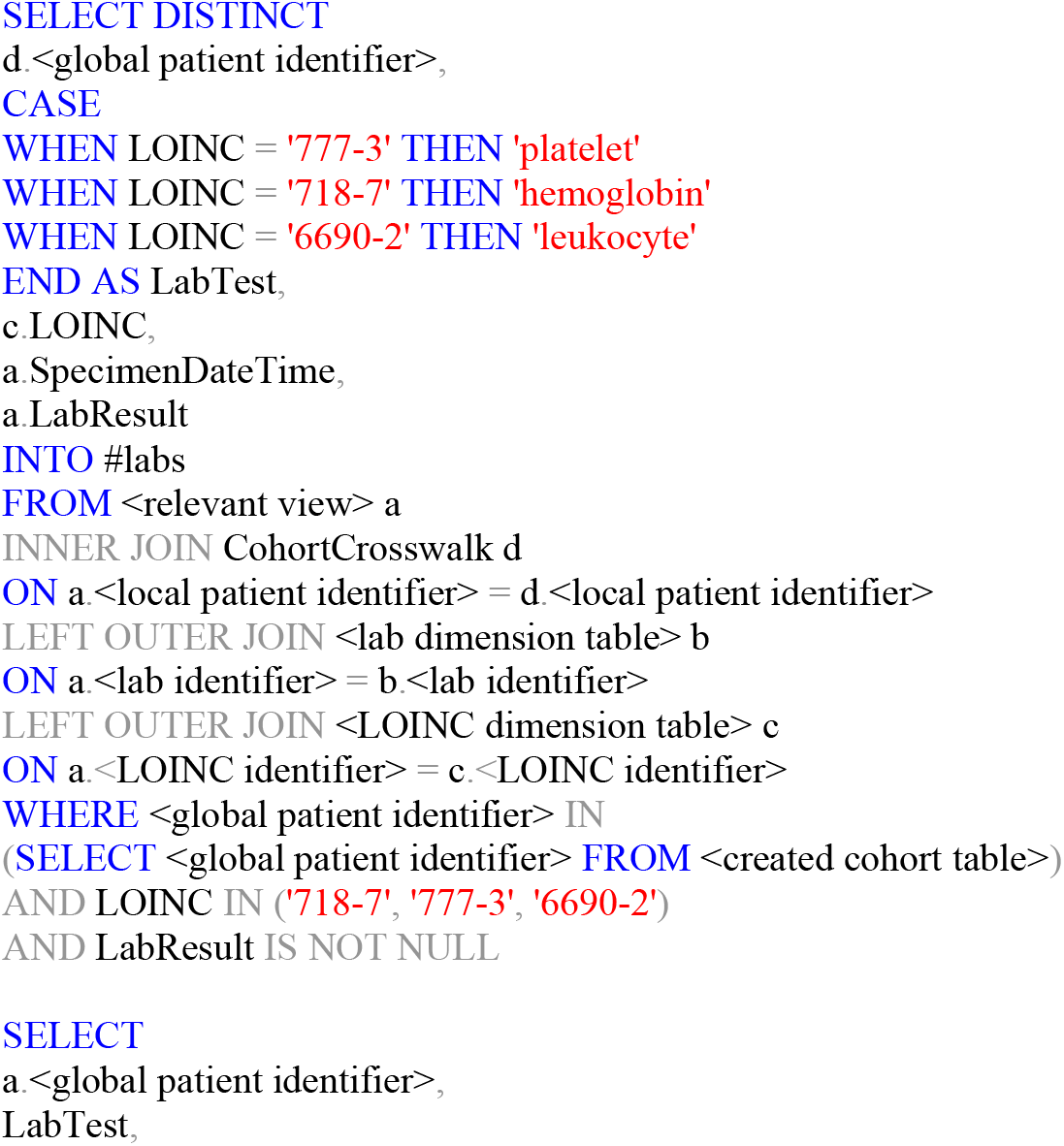

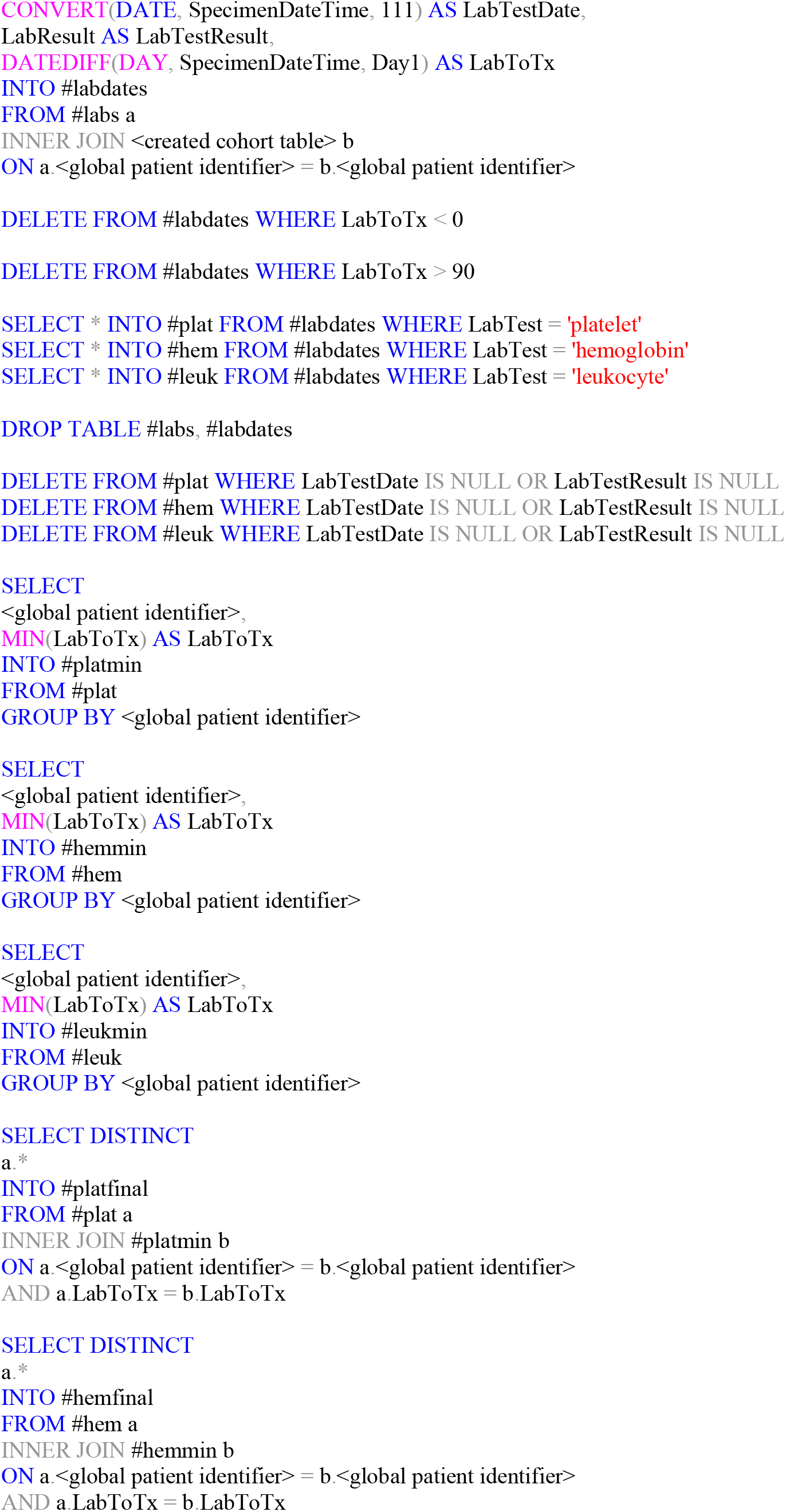

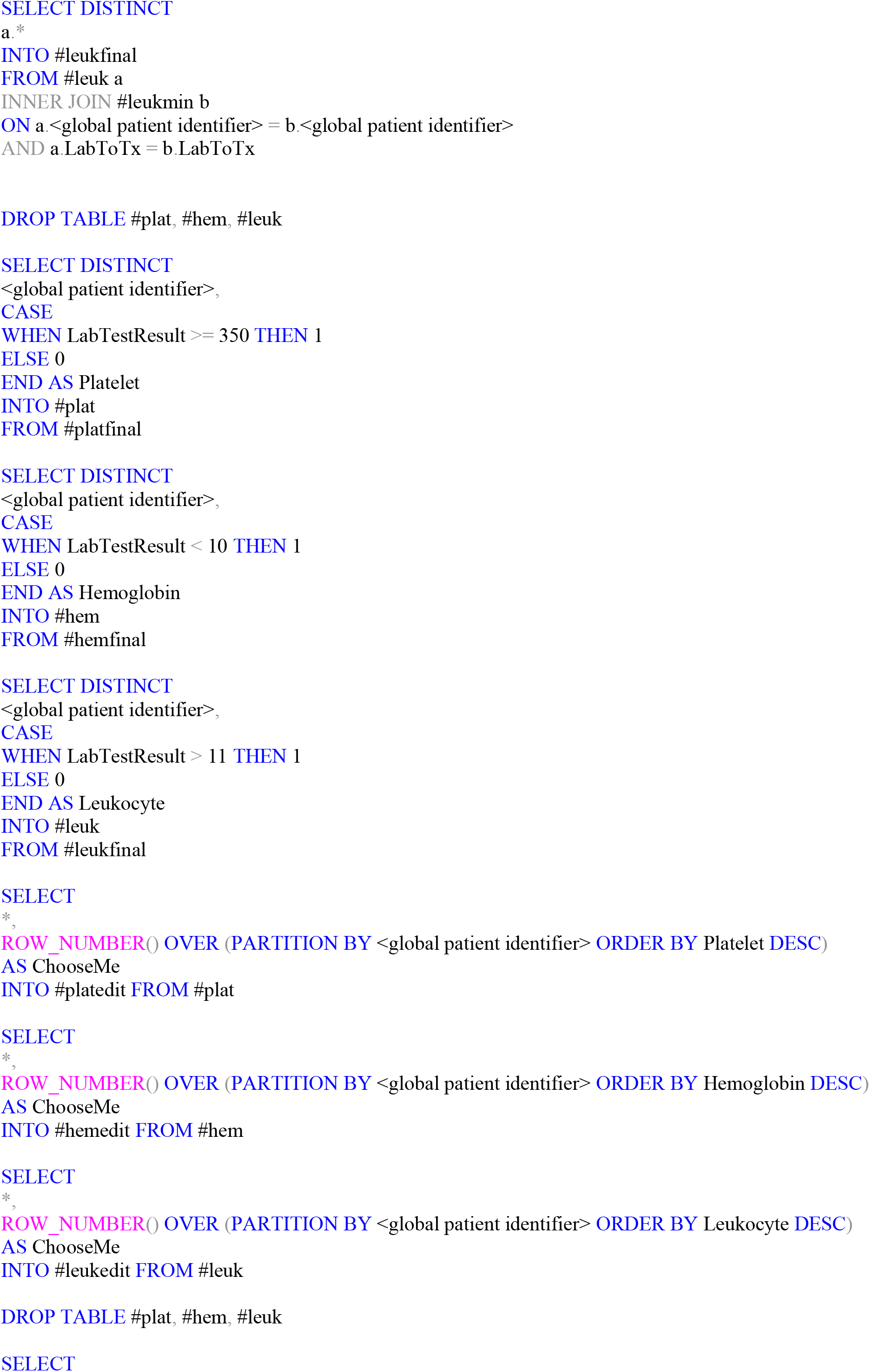

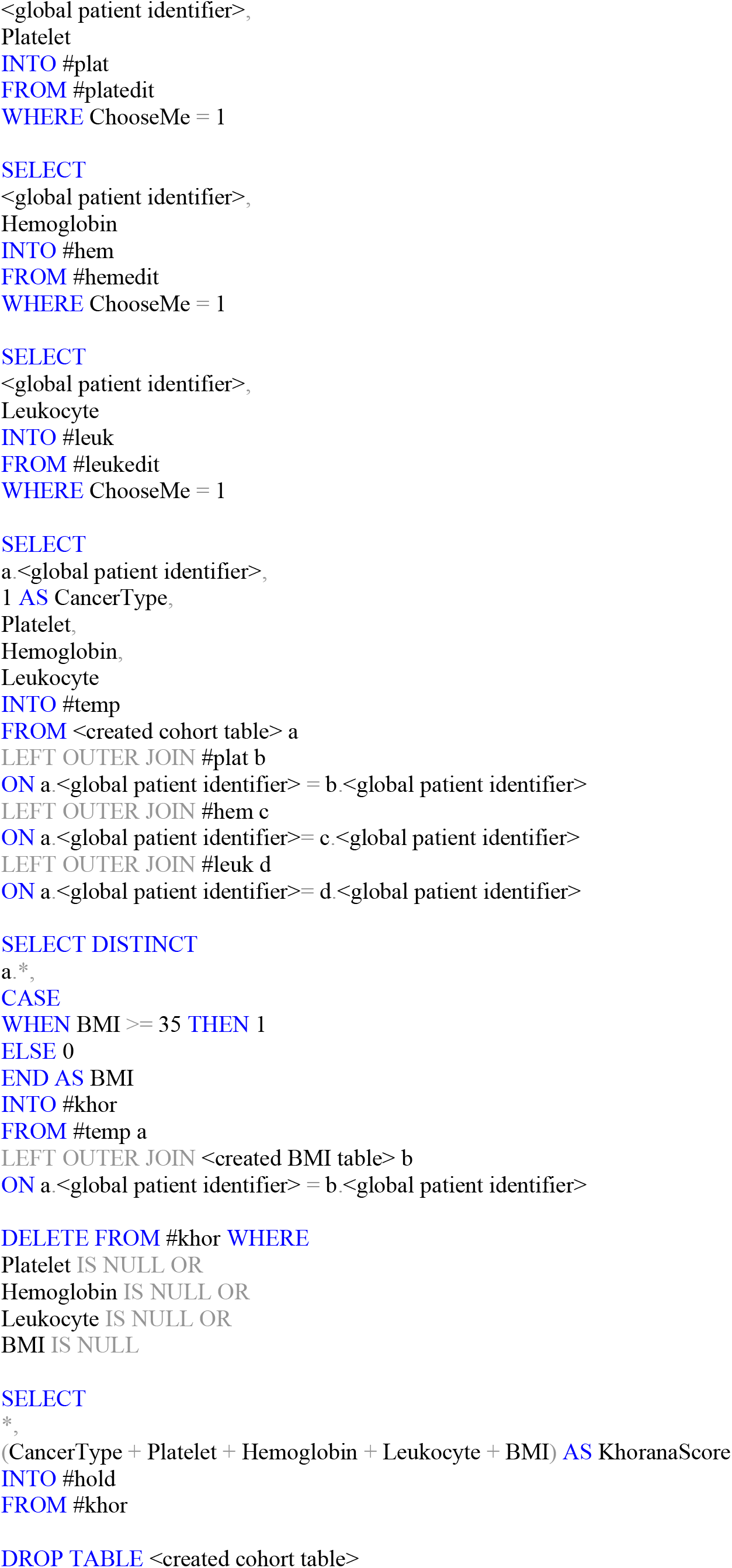

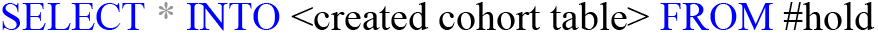

##### Calculate Charlson score

A custom R function was used to determine each patient’s Charlson score at the time of treatment start.

##### Identify TEEs

TEEs were identified using the same custom R function as described for cancer diagnoses. Using the resultant dataset, patients with a history of prior TEE were identified as any patient with an instance of a TEE diagnosis occurring before the first day of treatment, patients who experienced a TEE during treatment were identified as any patient with an instance of a TEE diagnosis between days 1 and 180 of treatment, and patients with an instance of a TEE diagnosis occurring exactly on the first day of treatment were deleted from the cohort.

#### Data Analysis

##### Prepare analytical table

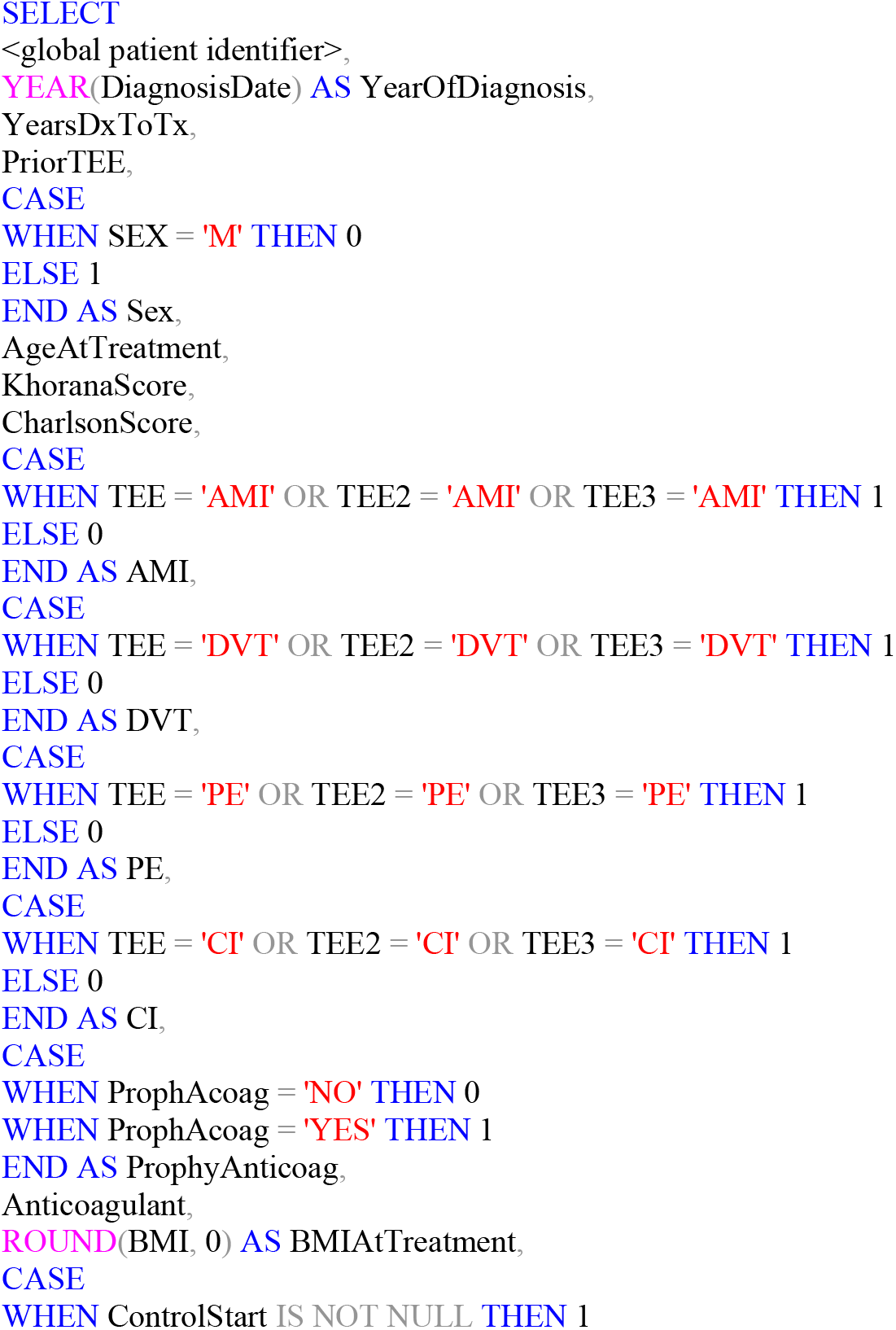

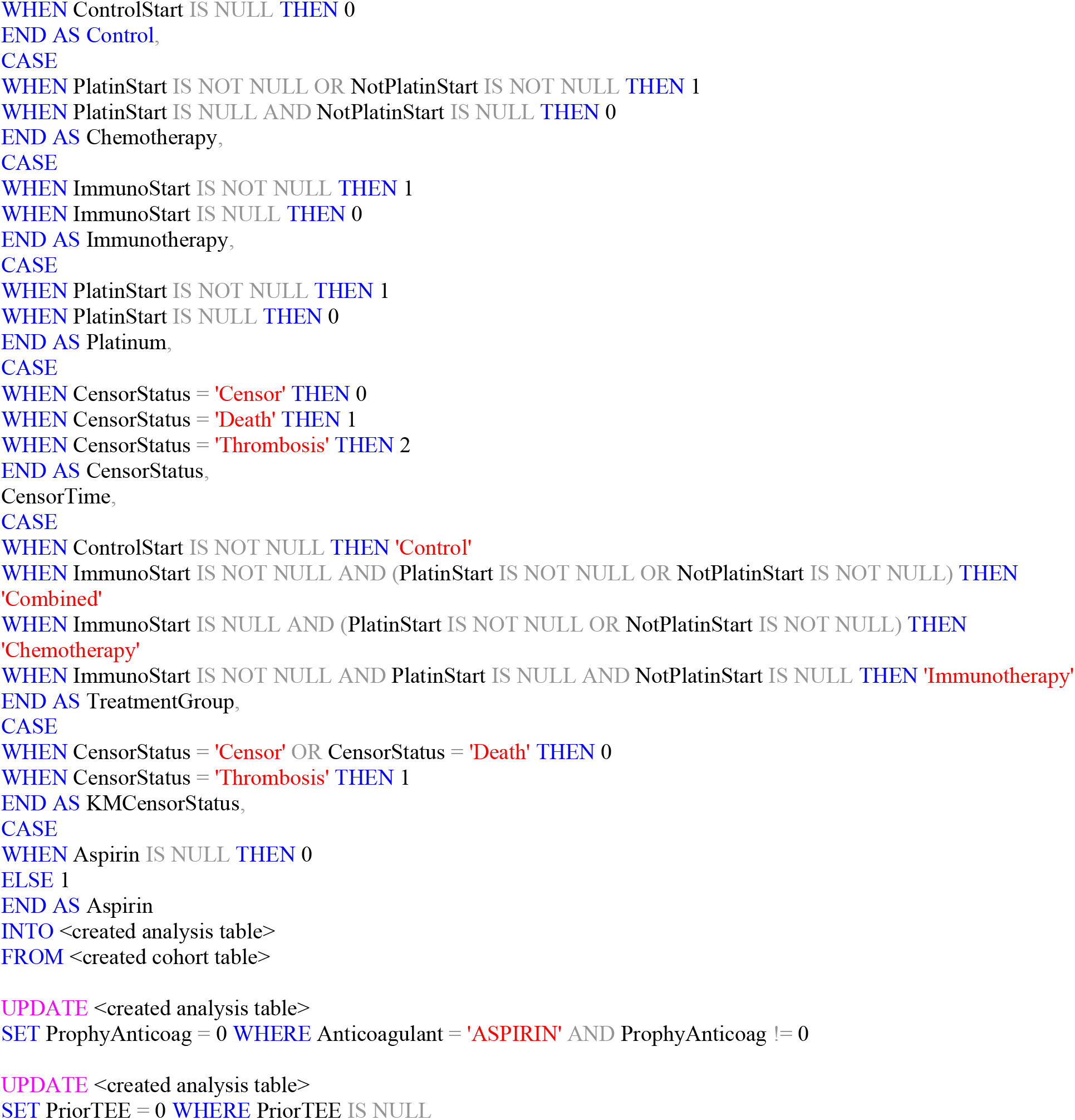

##### Develop descriptive statistics of cohort

The statistical analysis was performed in R (version 4.0.2; citation noted in main body of paper). The analysis table created in the research database was imported into R.

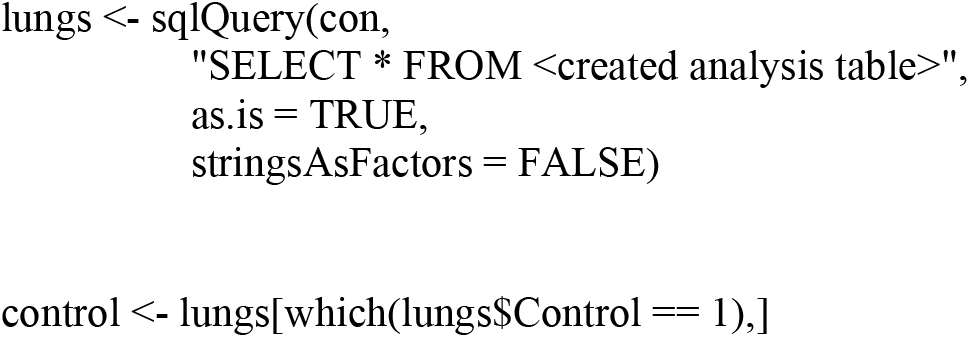

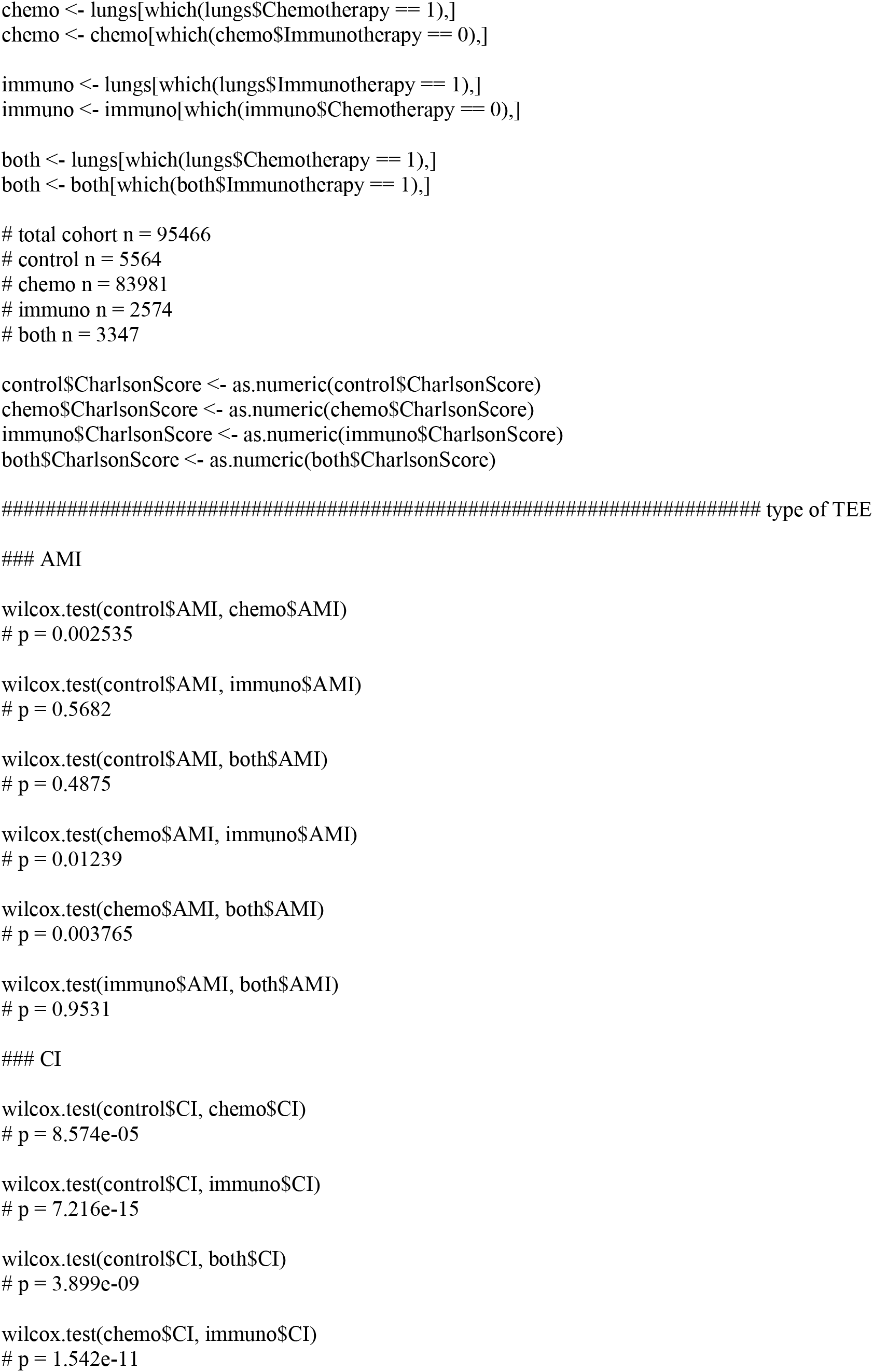

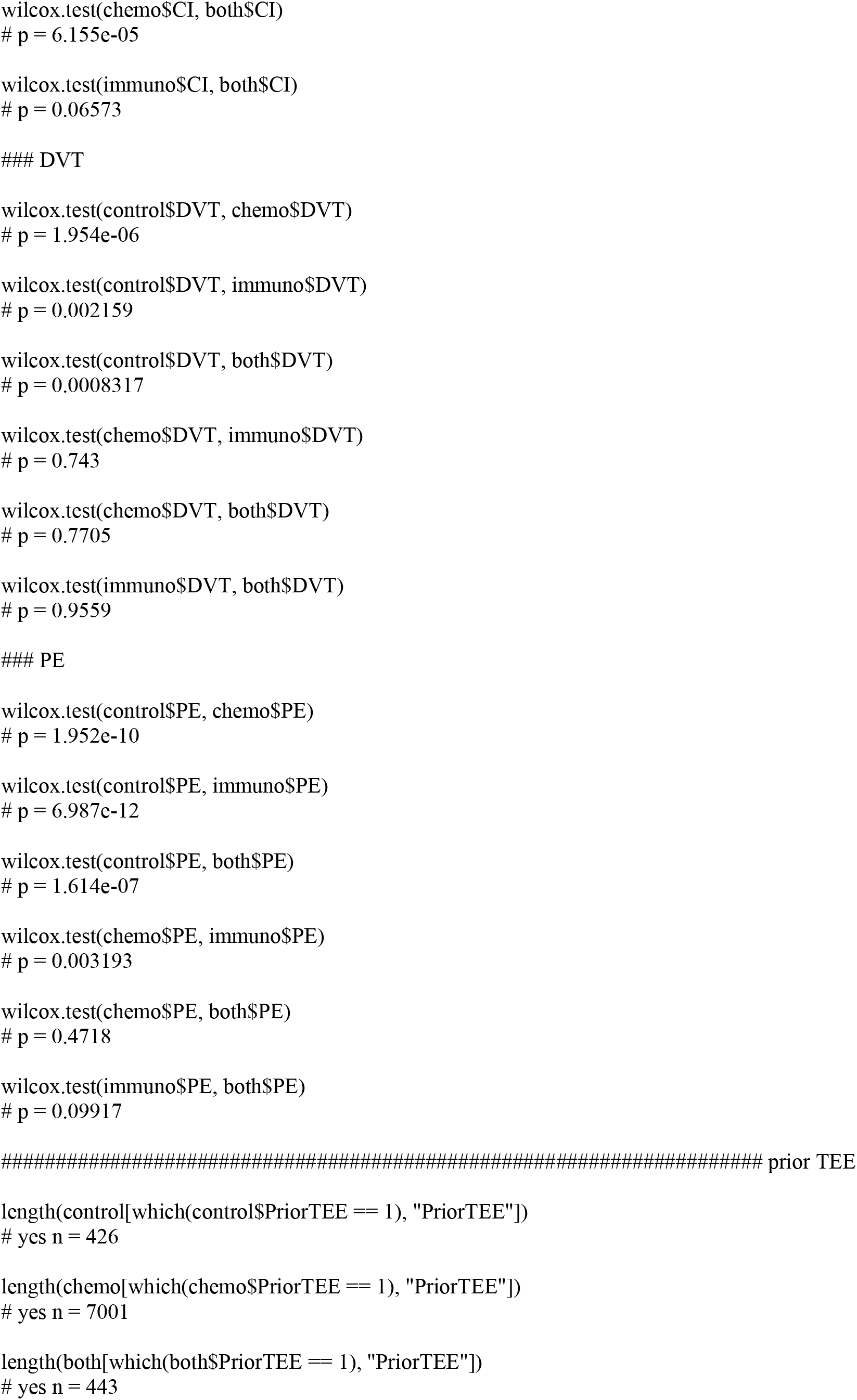

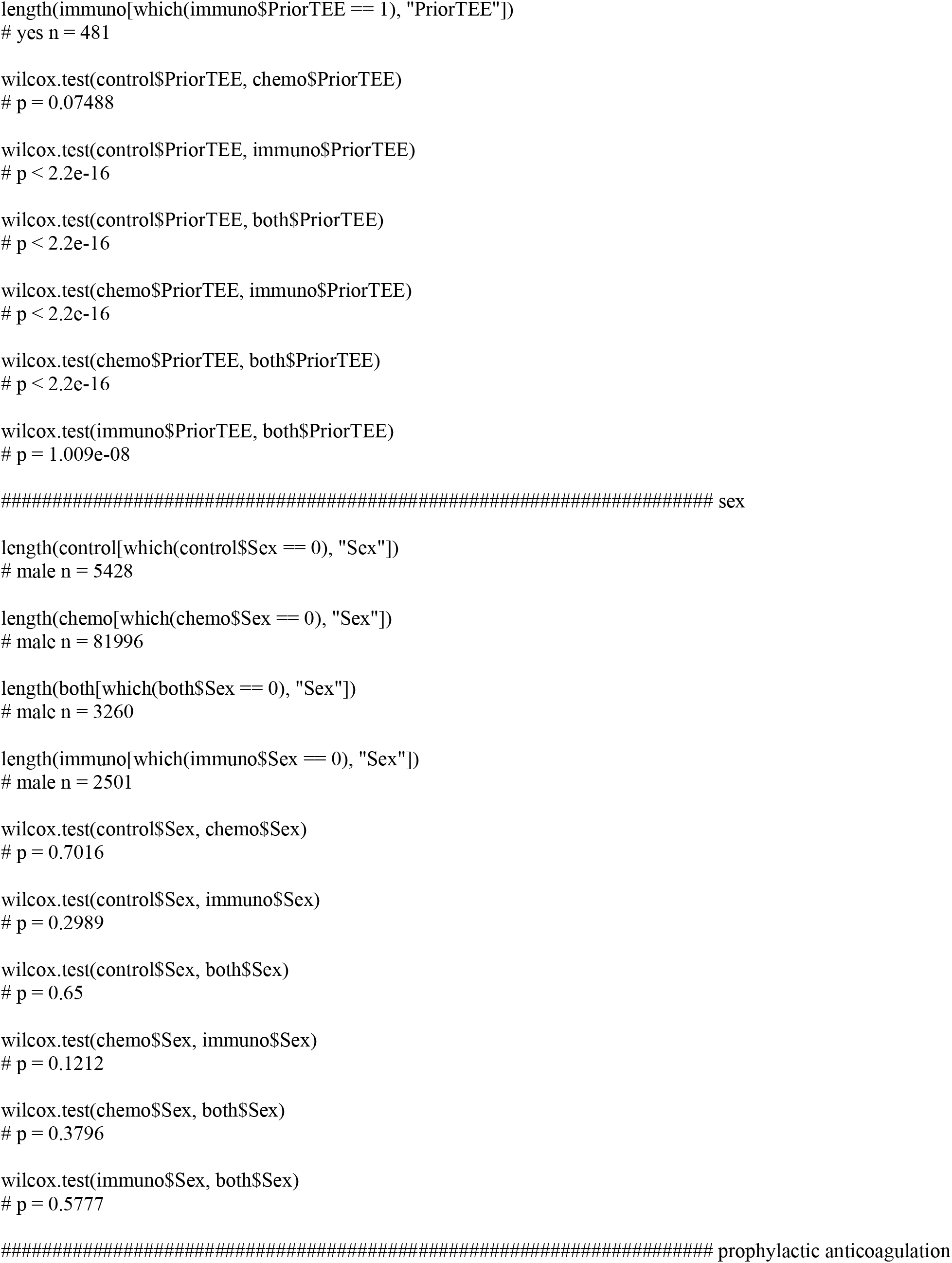

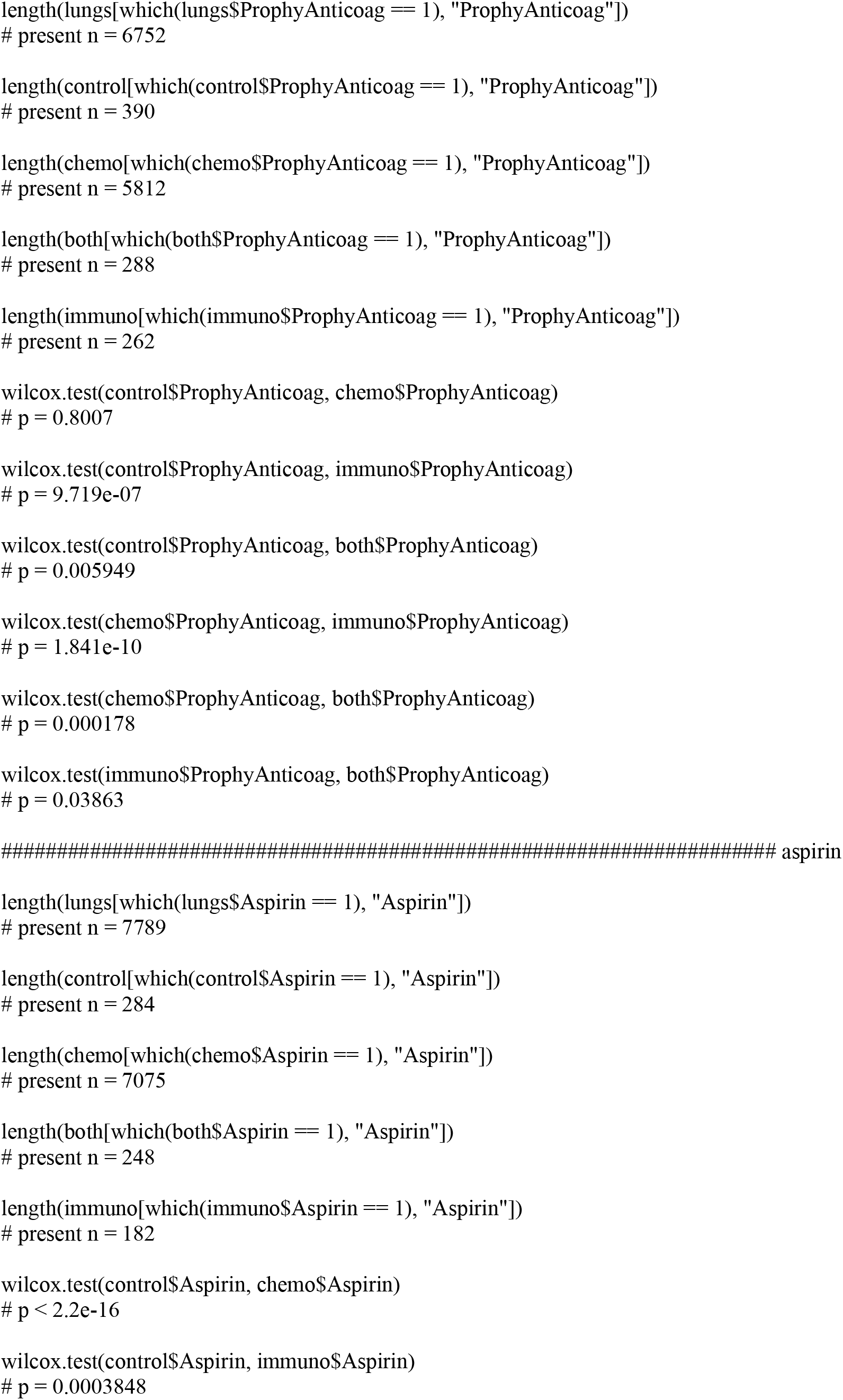

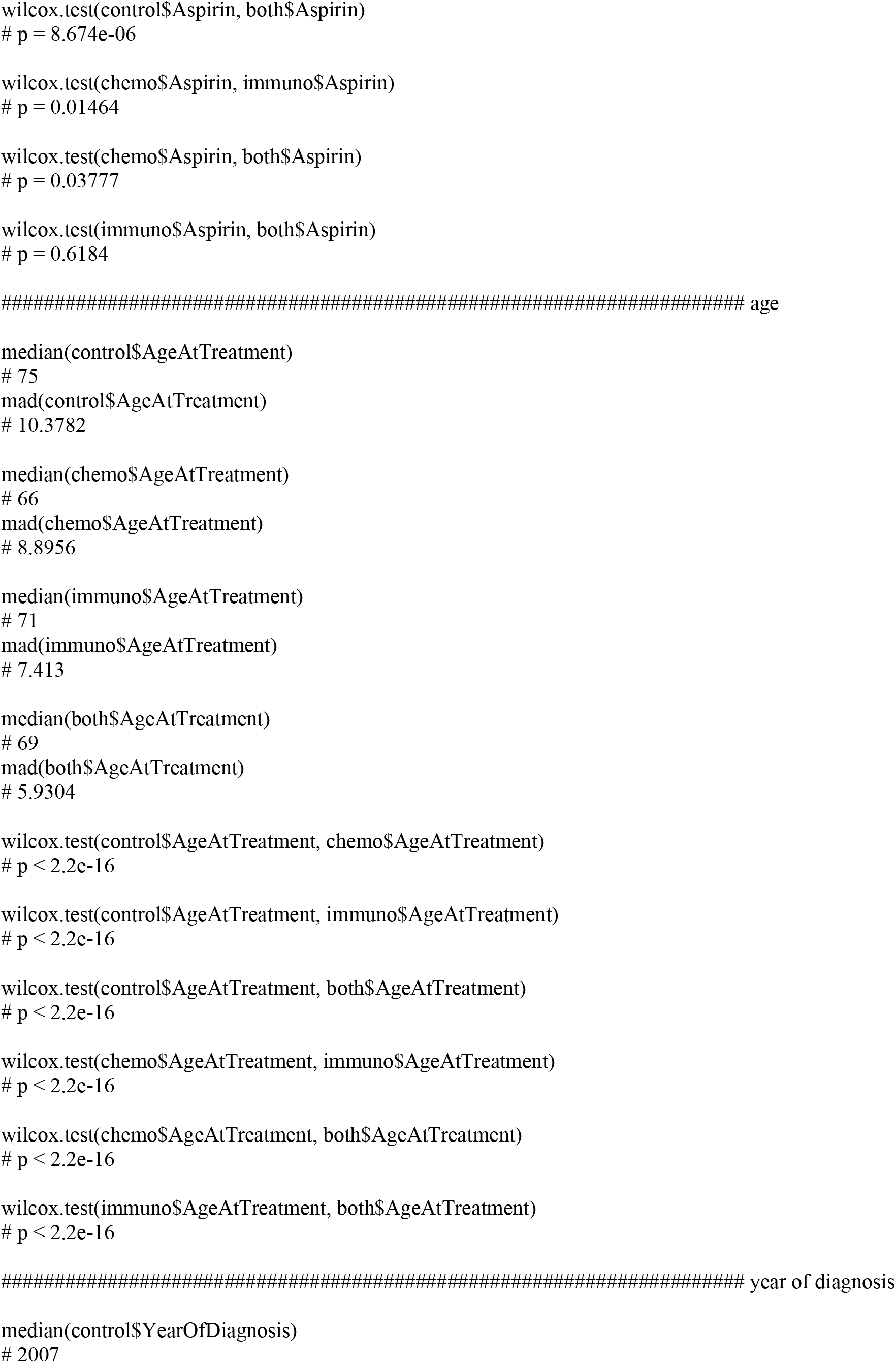

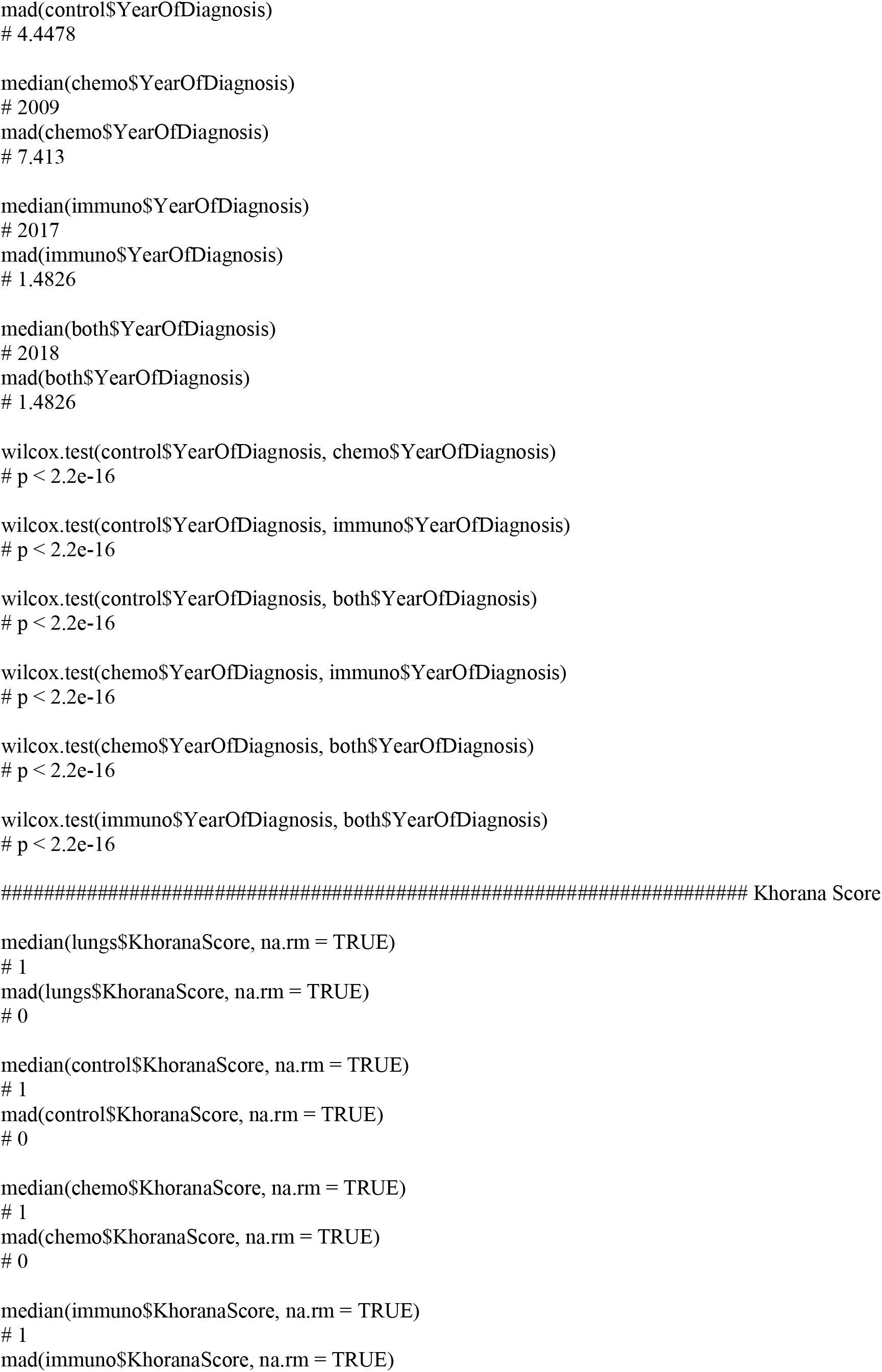

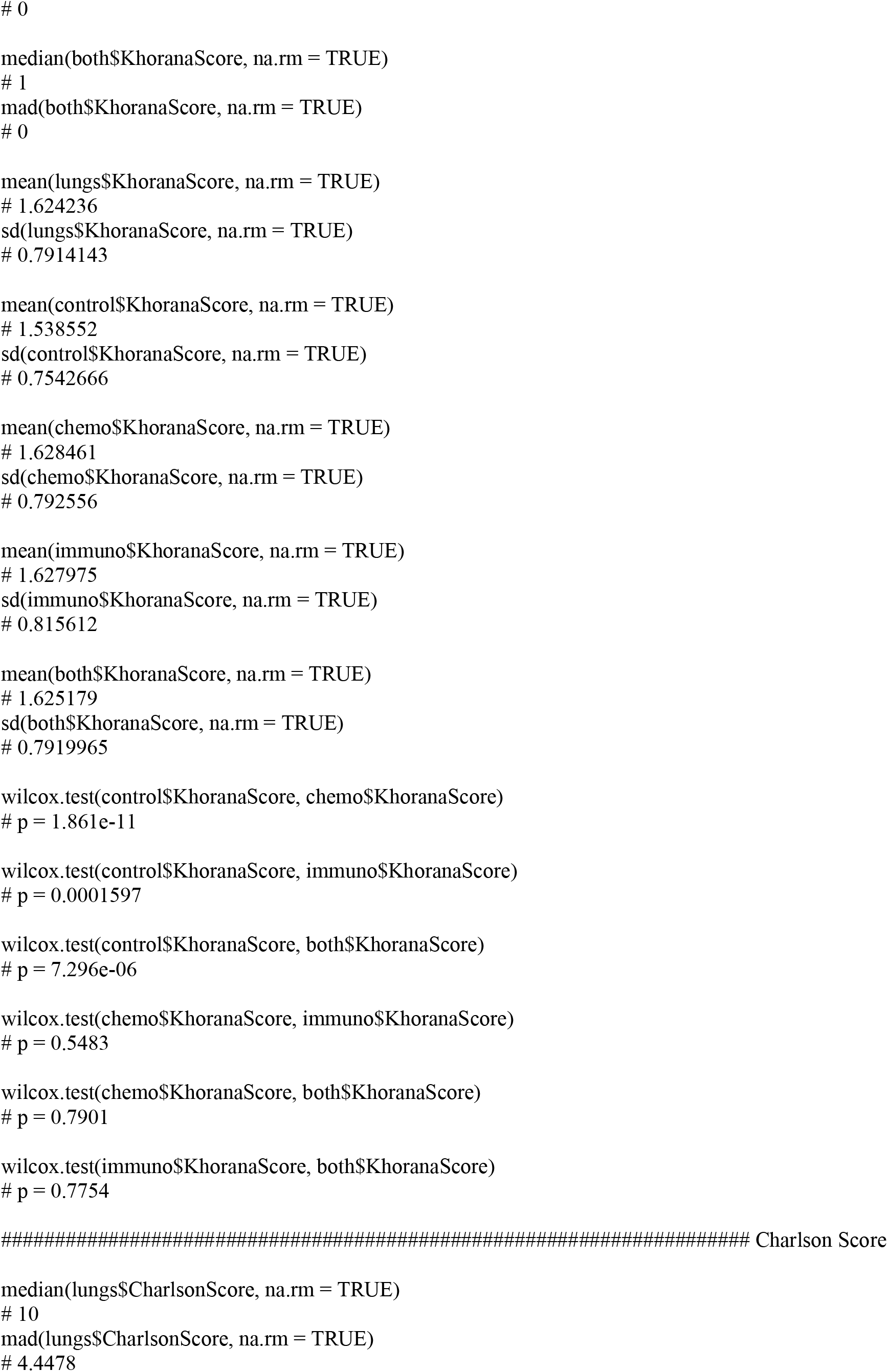

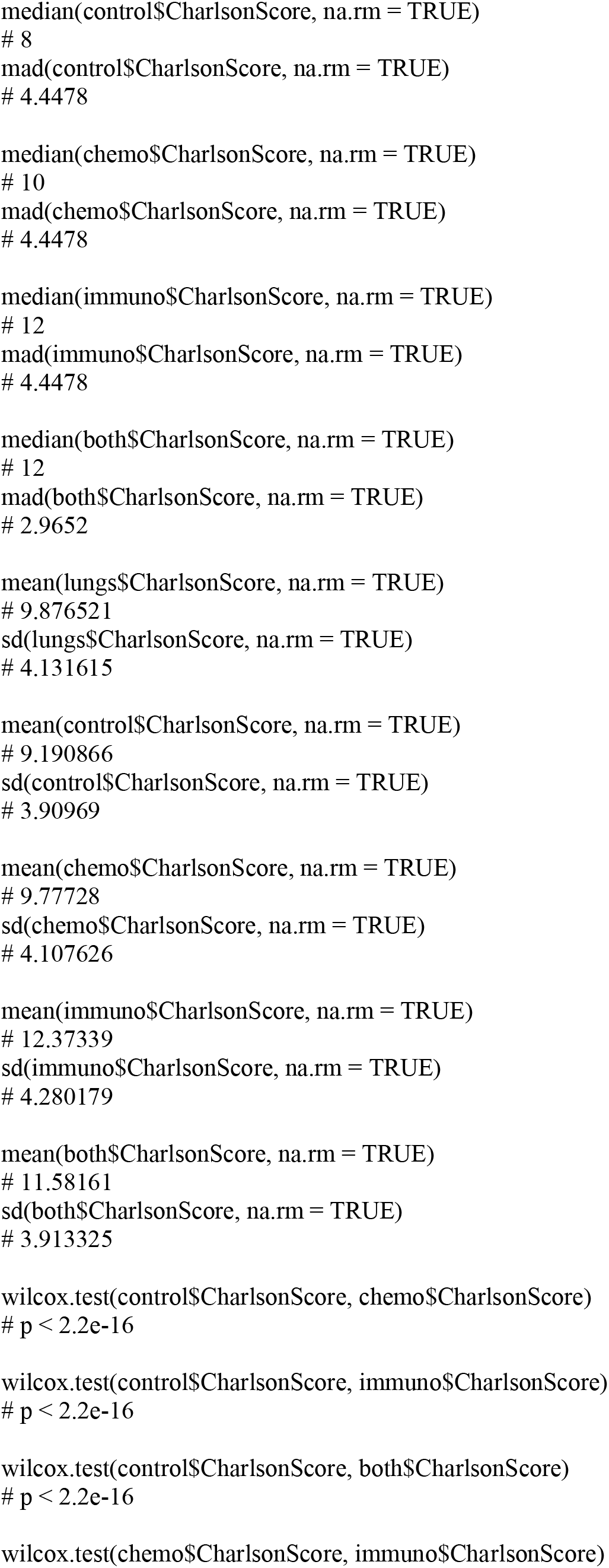

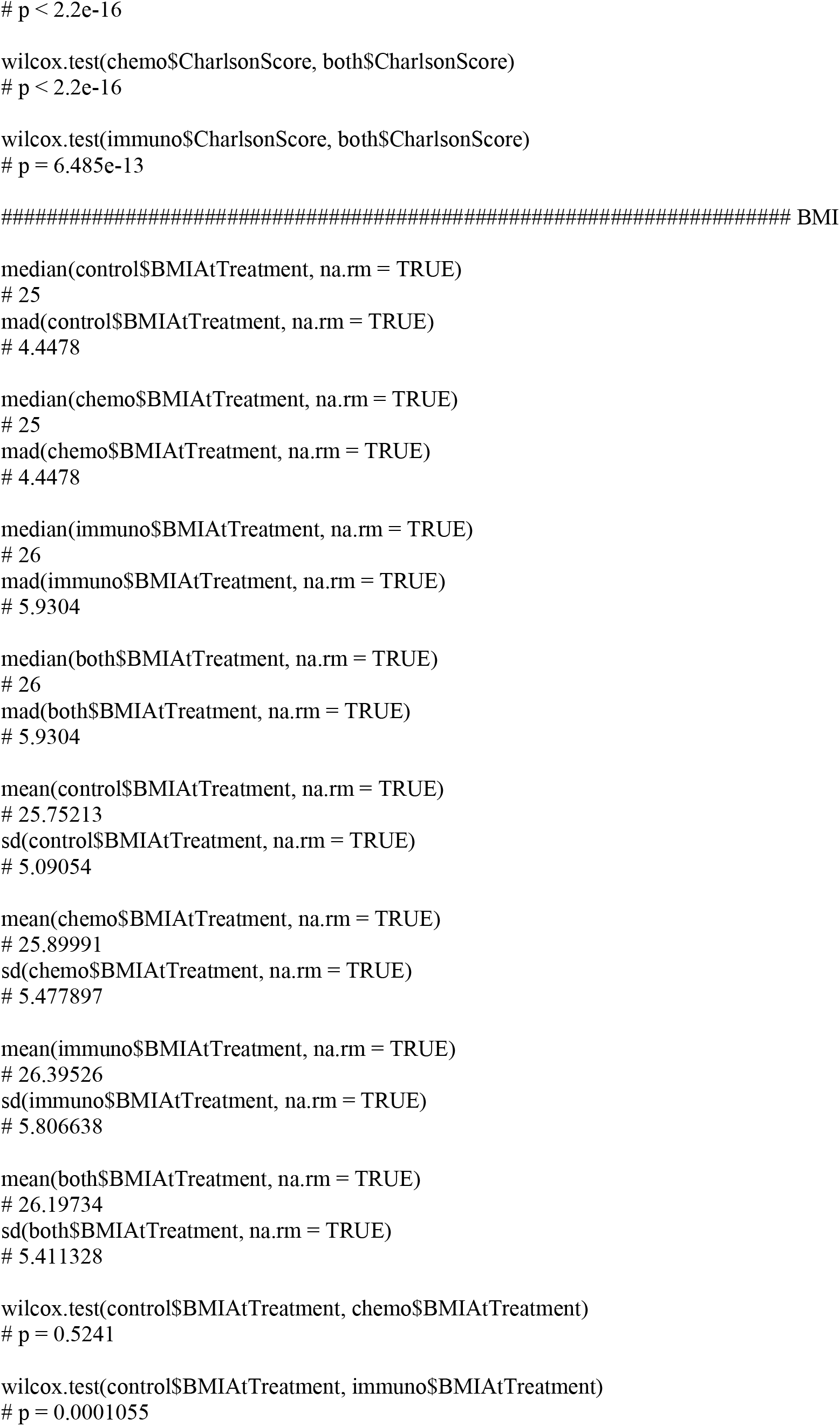

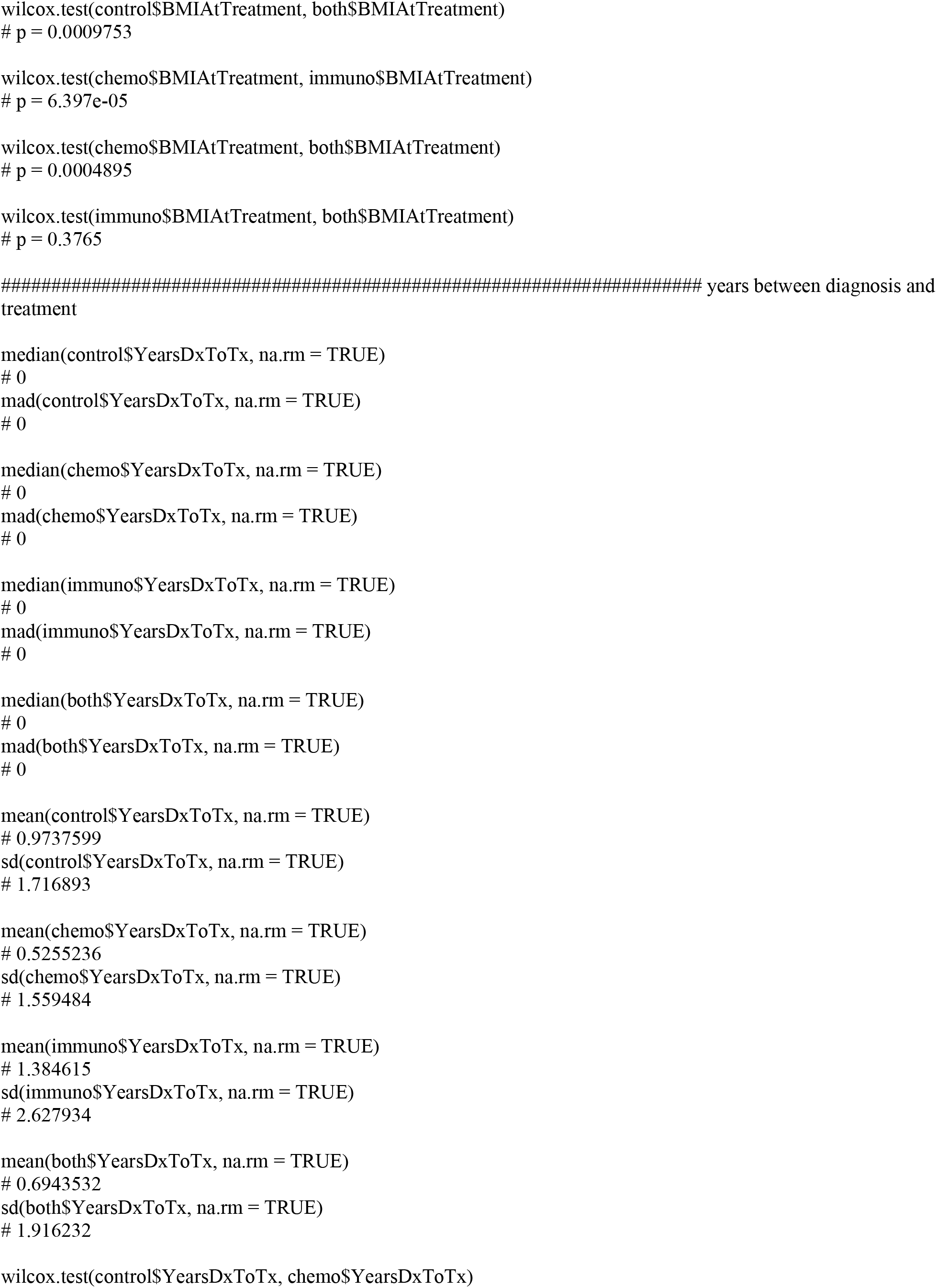

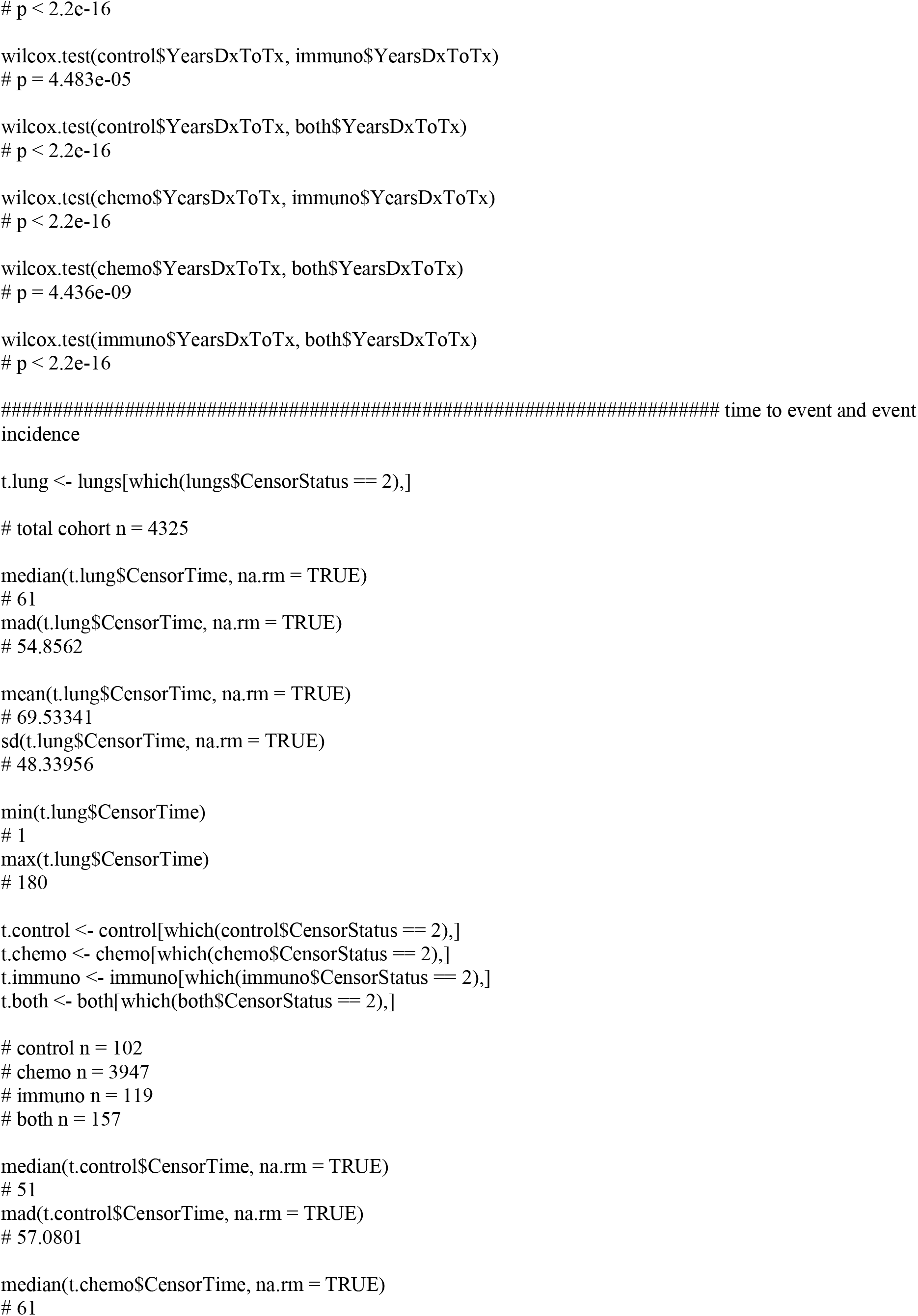

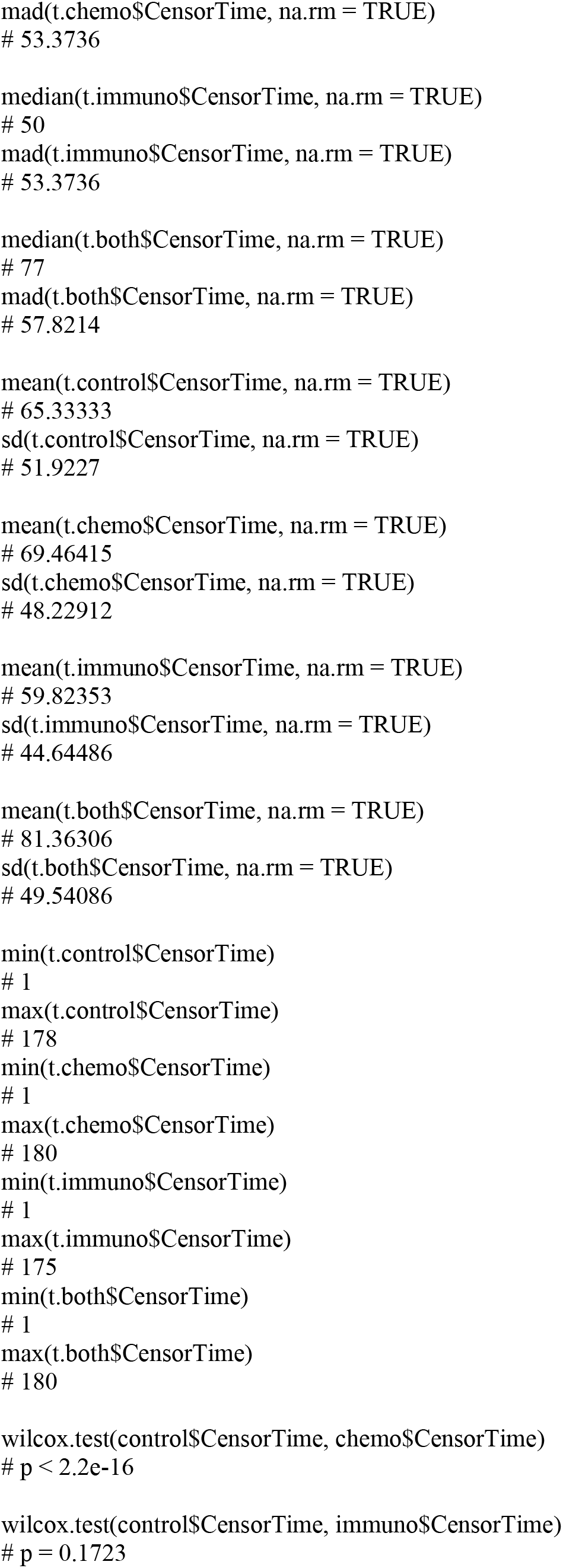

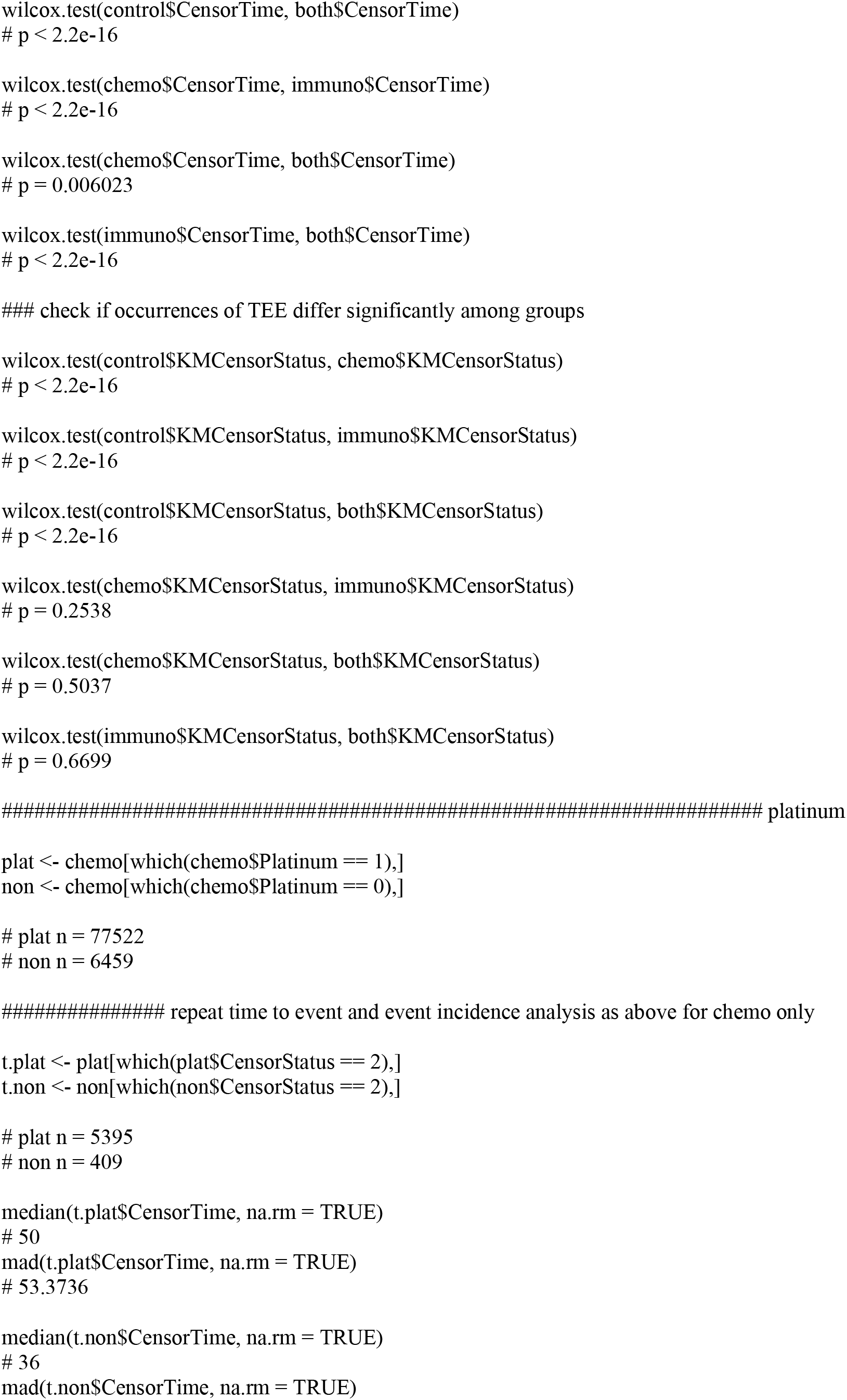

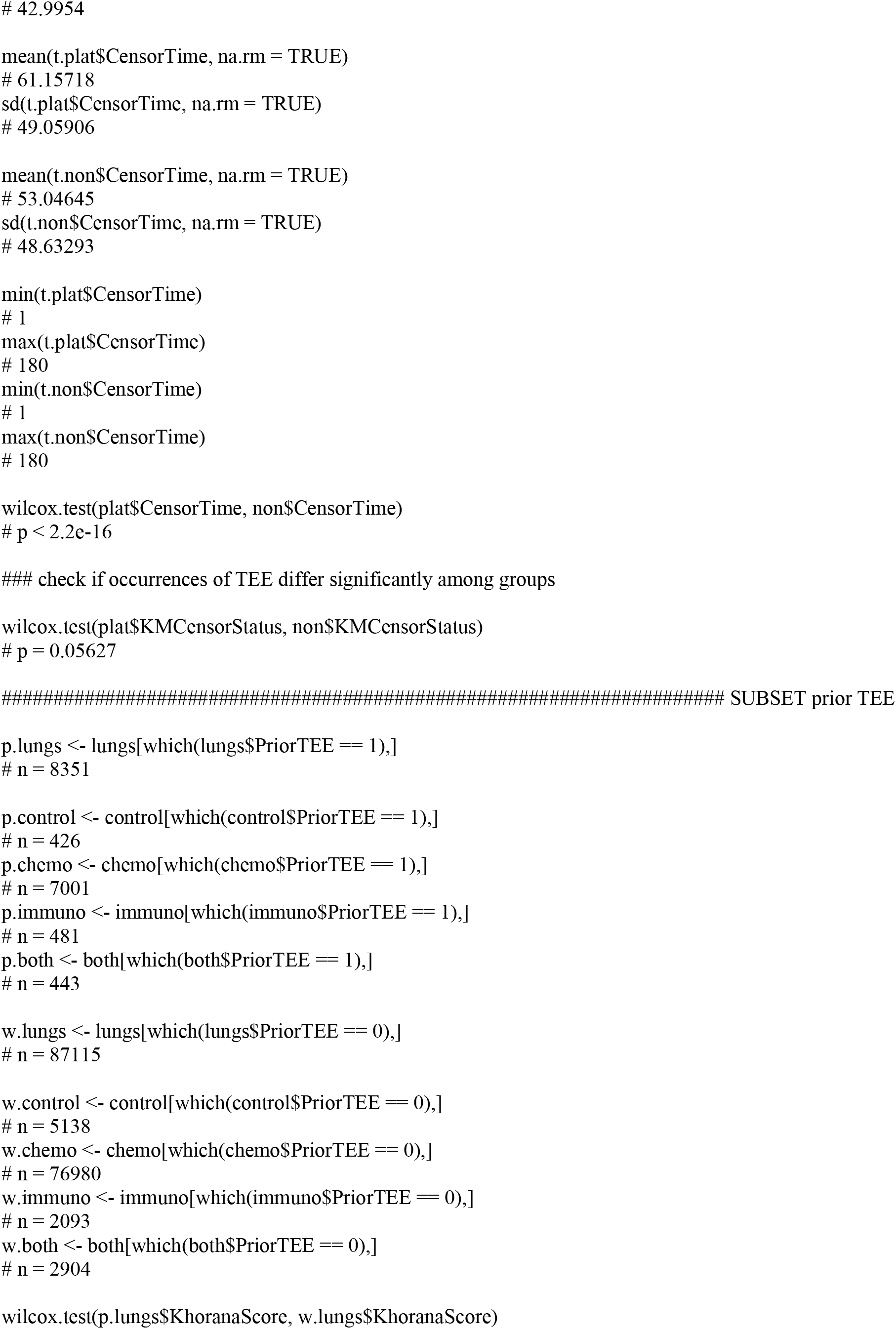

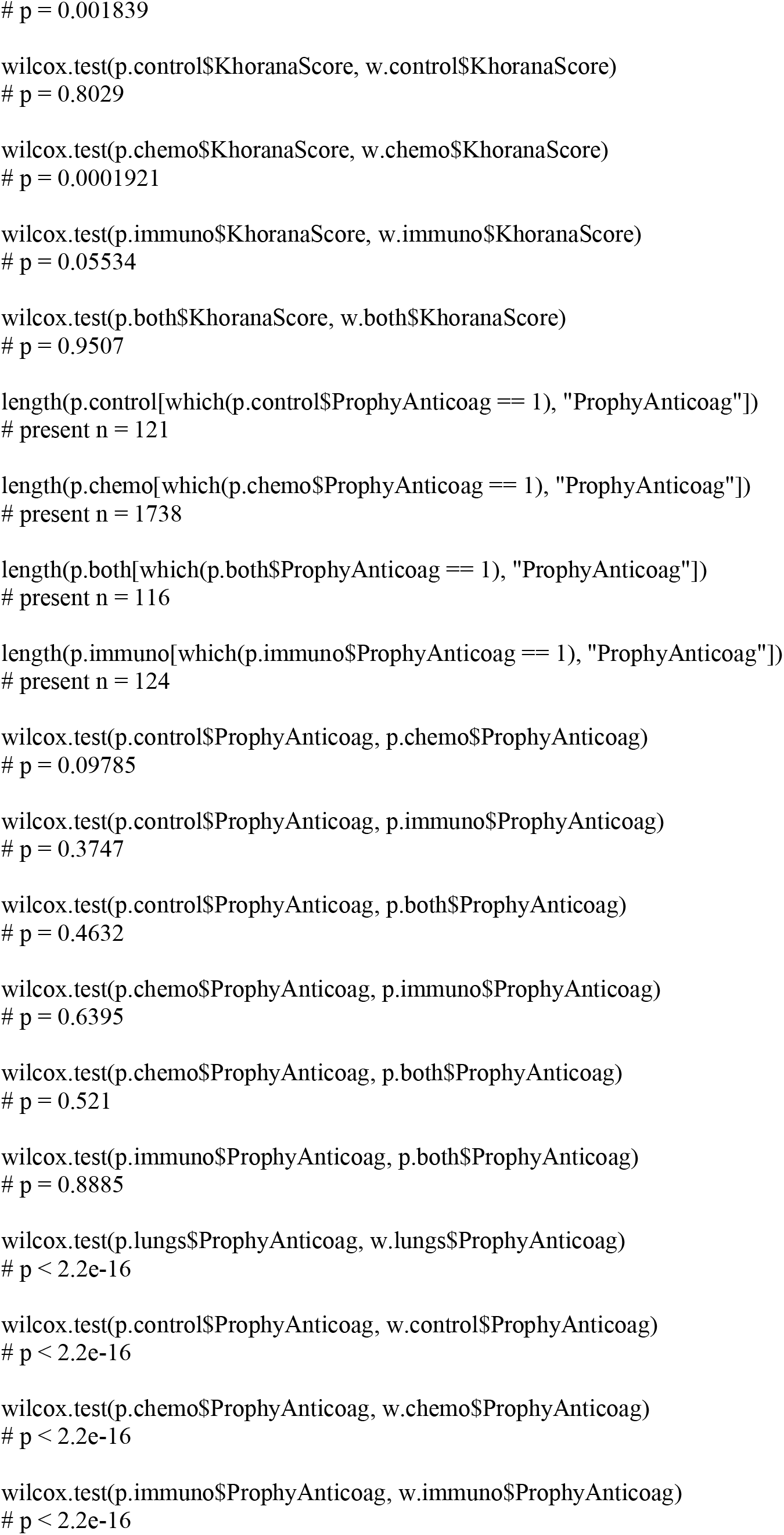

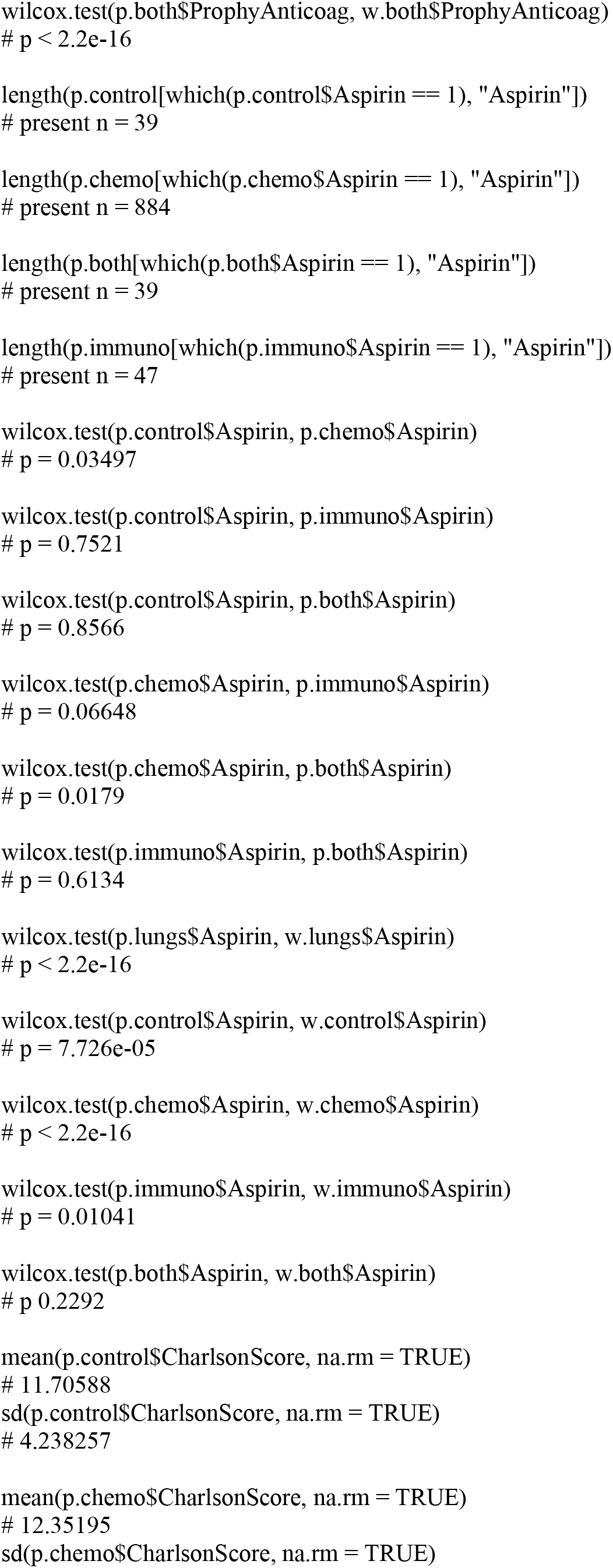

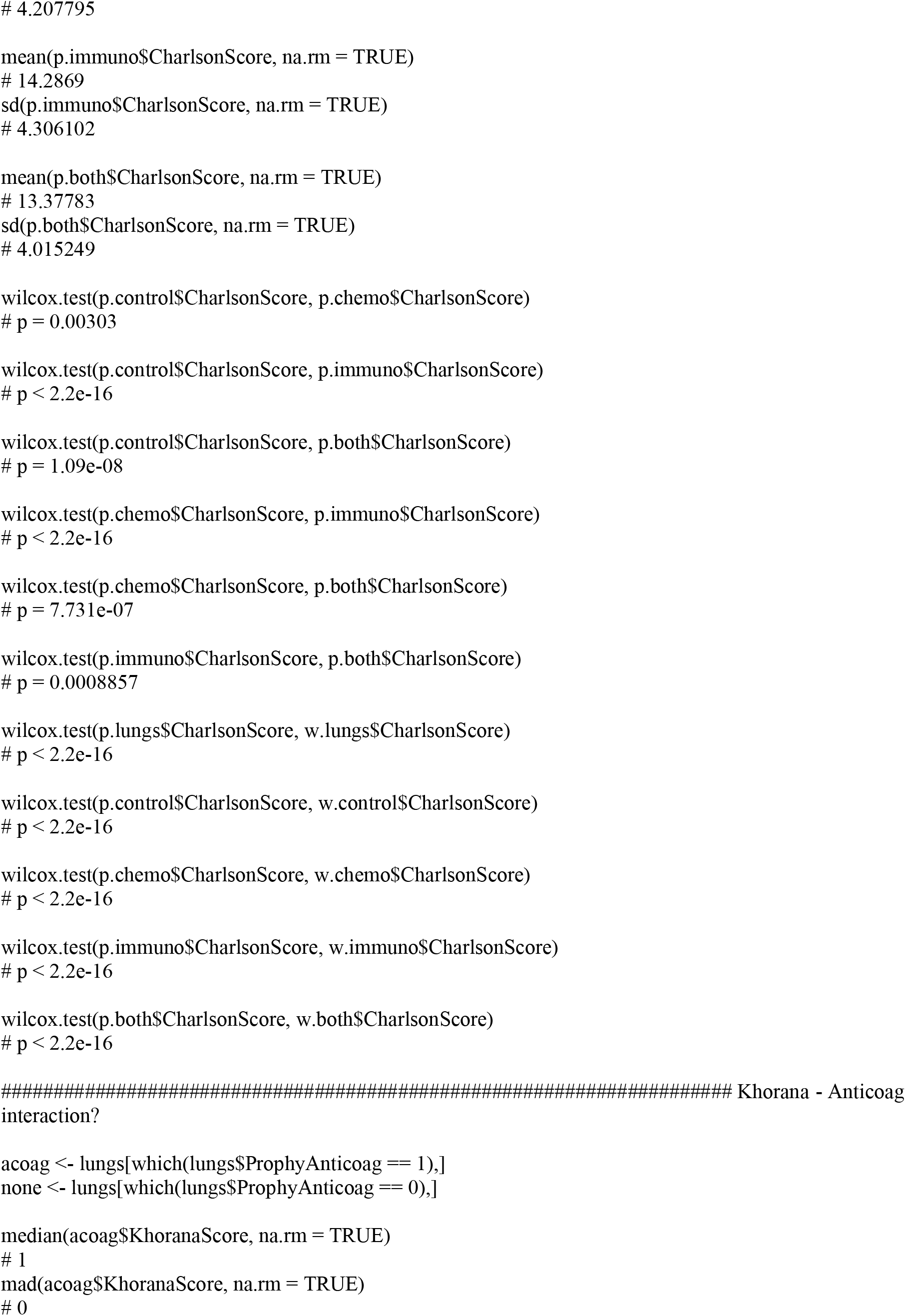

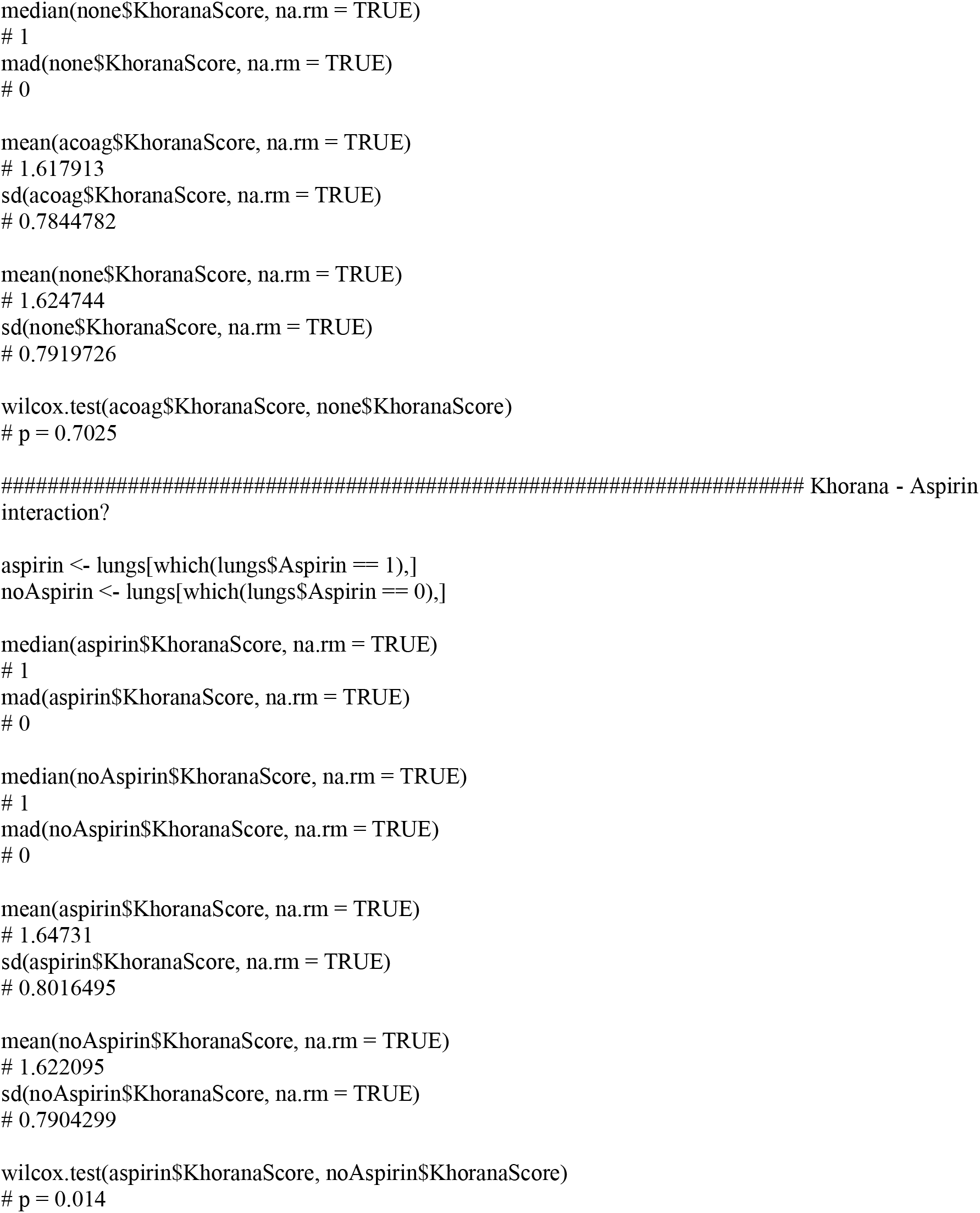

##### Perform Kaplan-Meier analysis

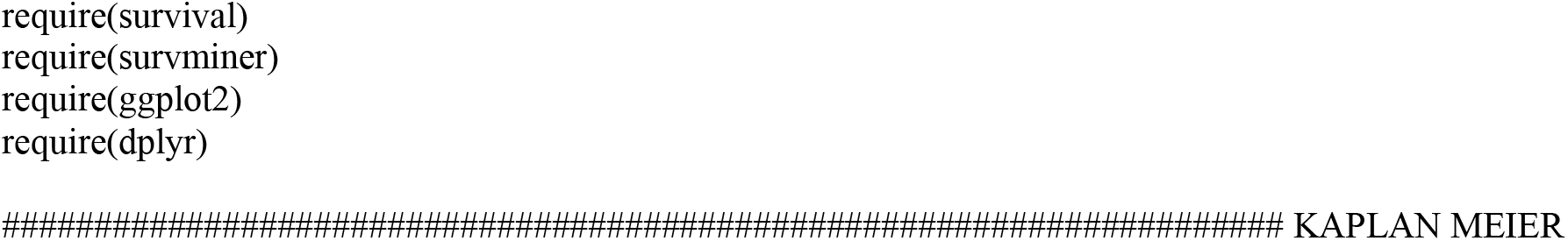

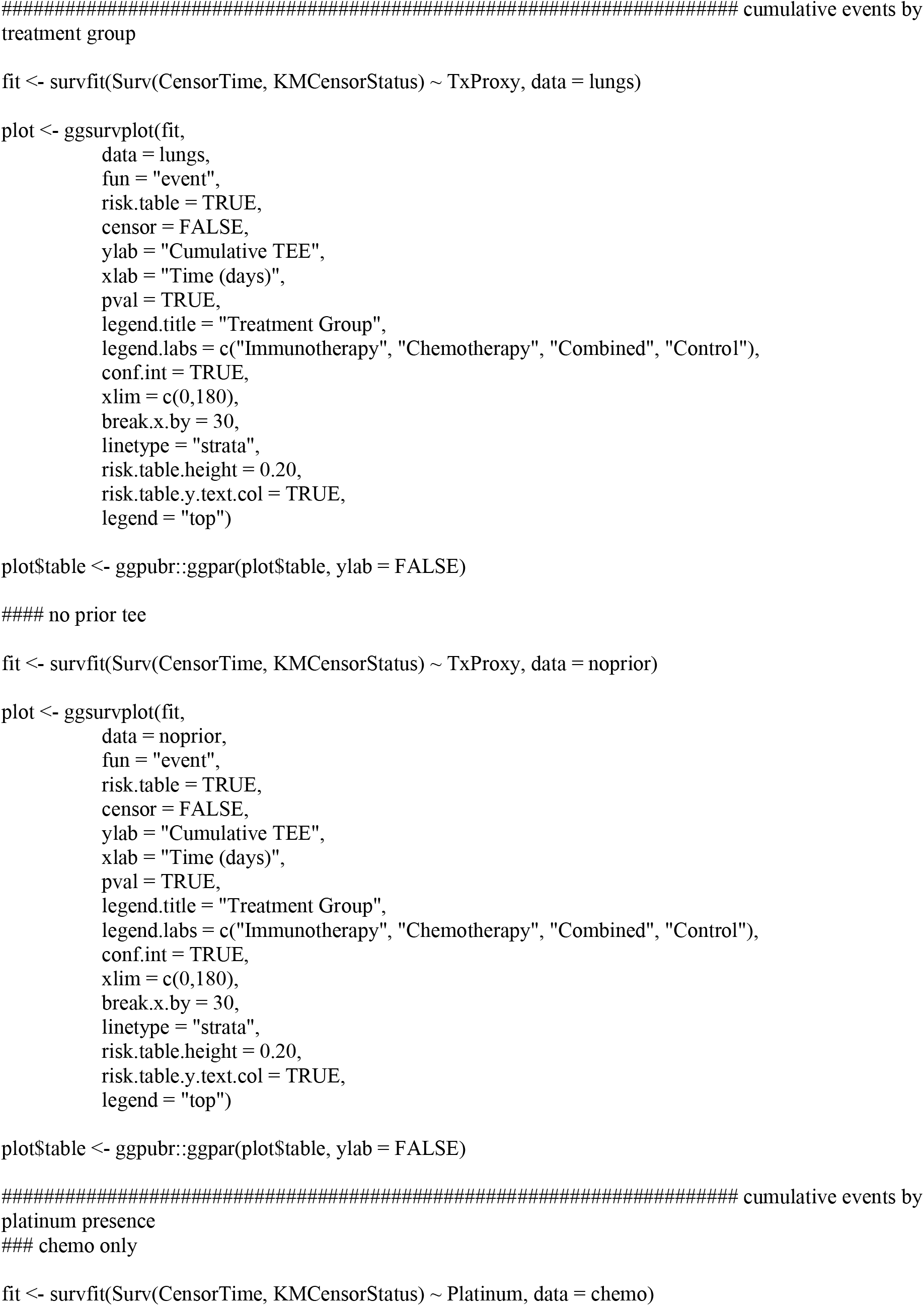

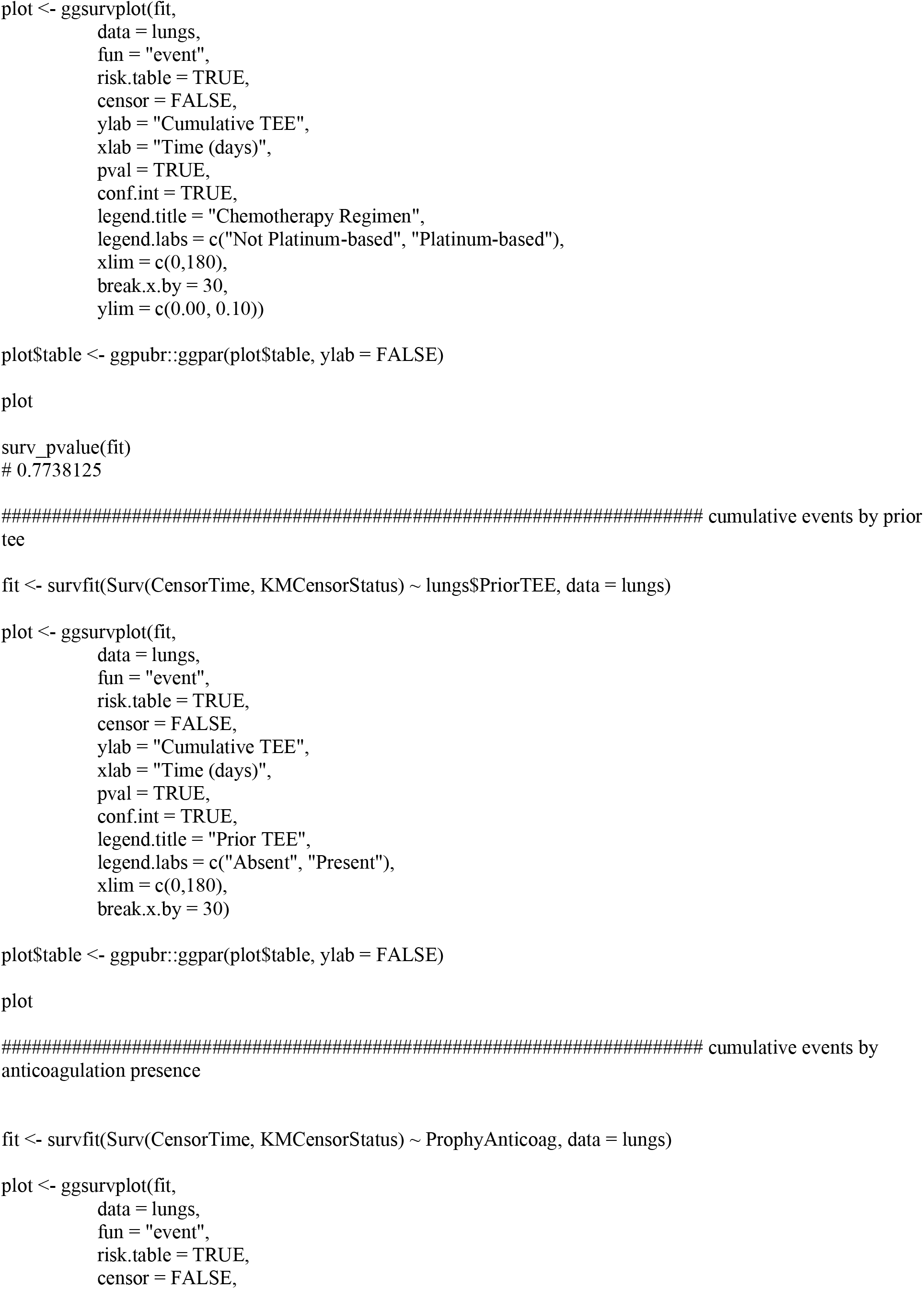

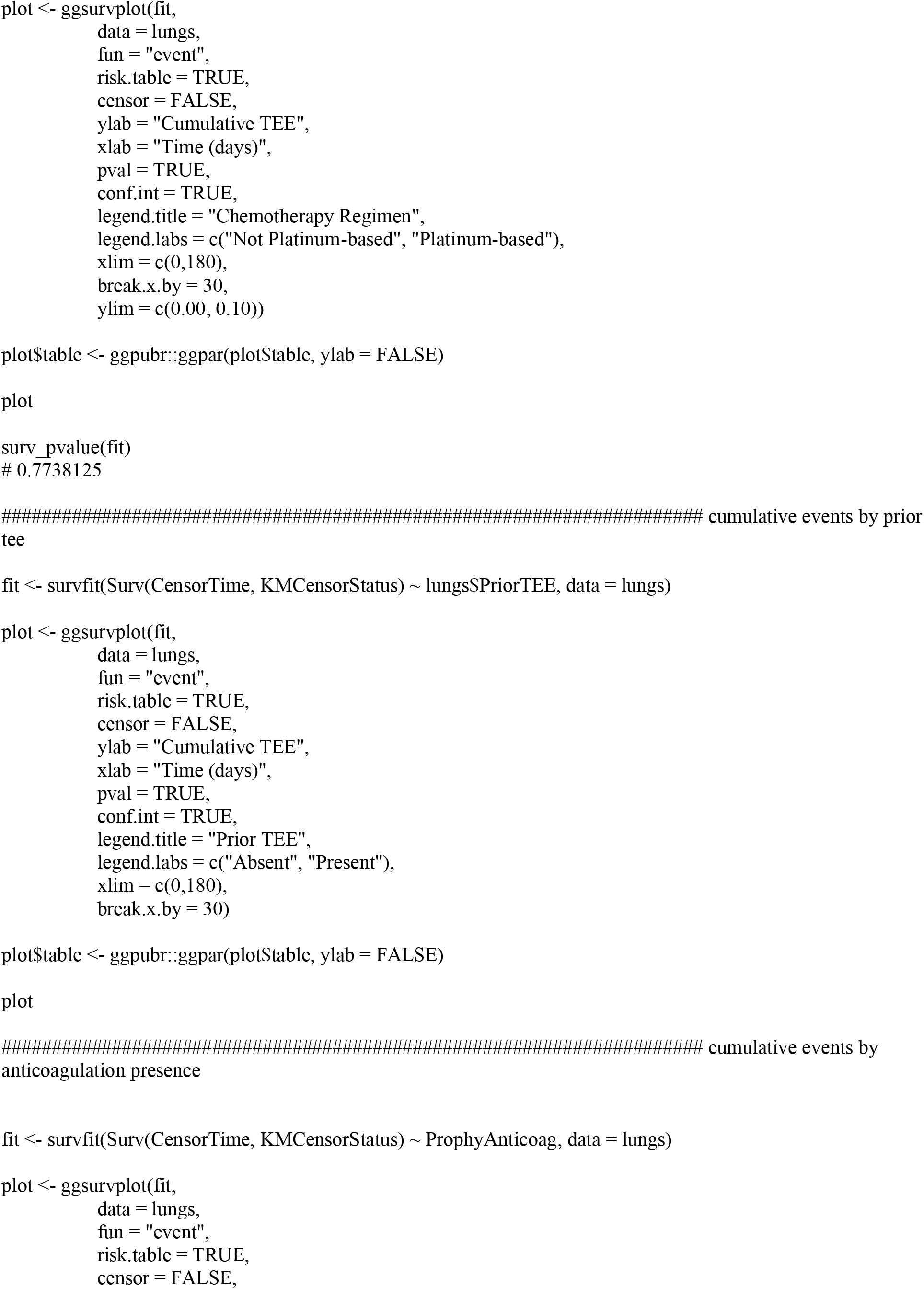

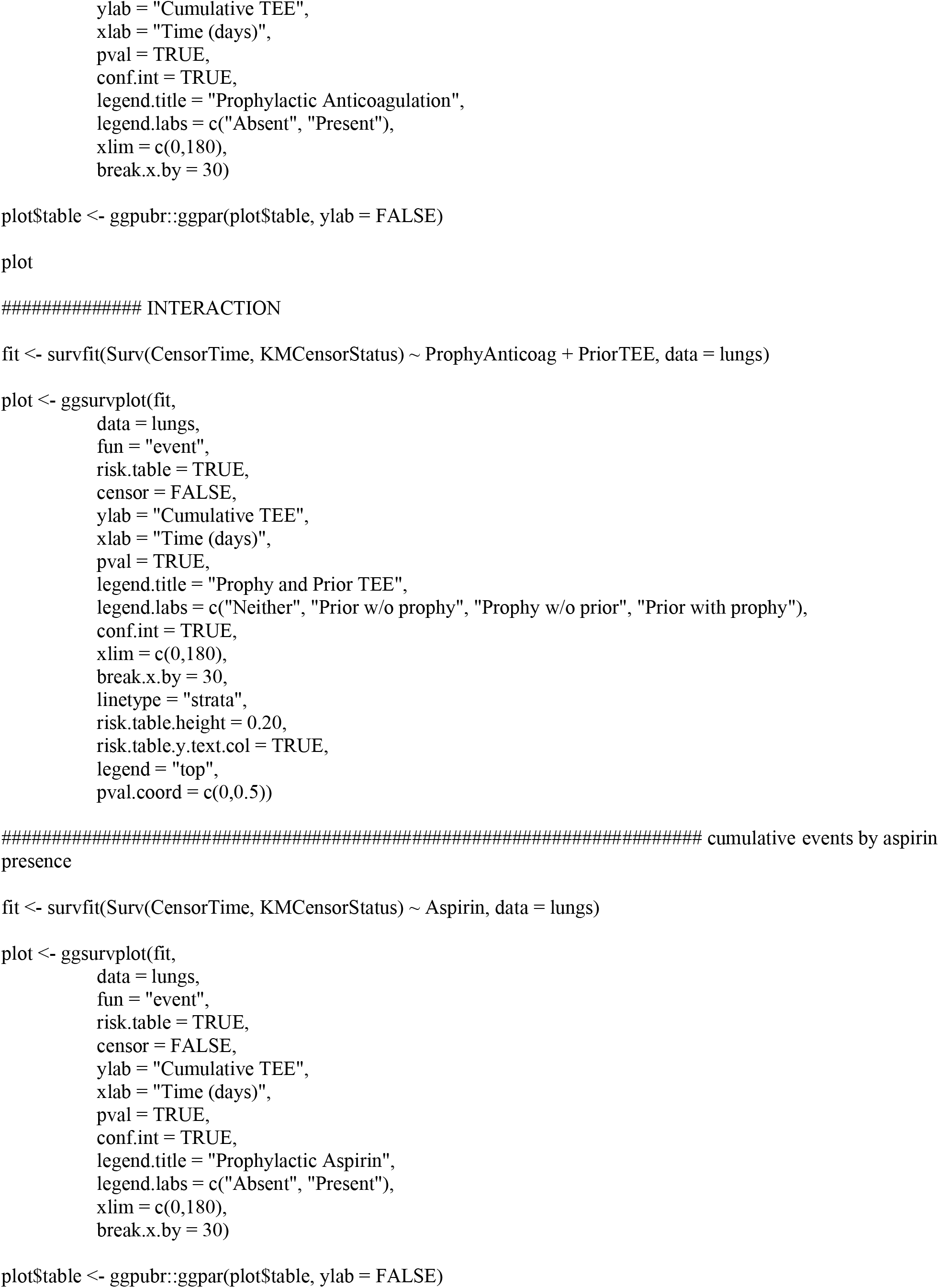

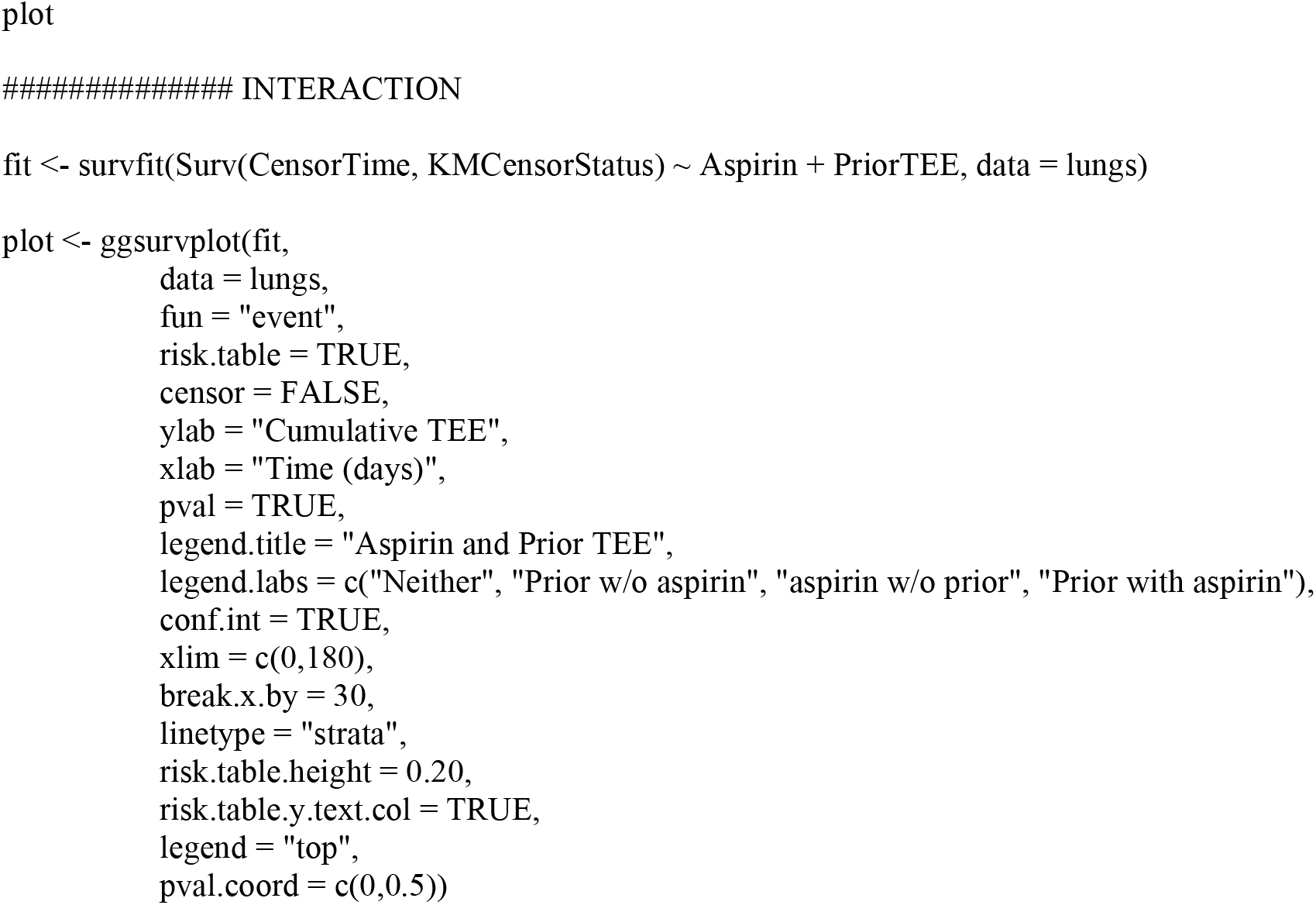

##### Perform Cox Proportional Hazard analysis

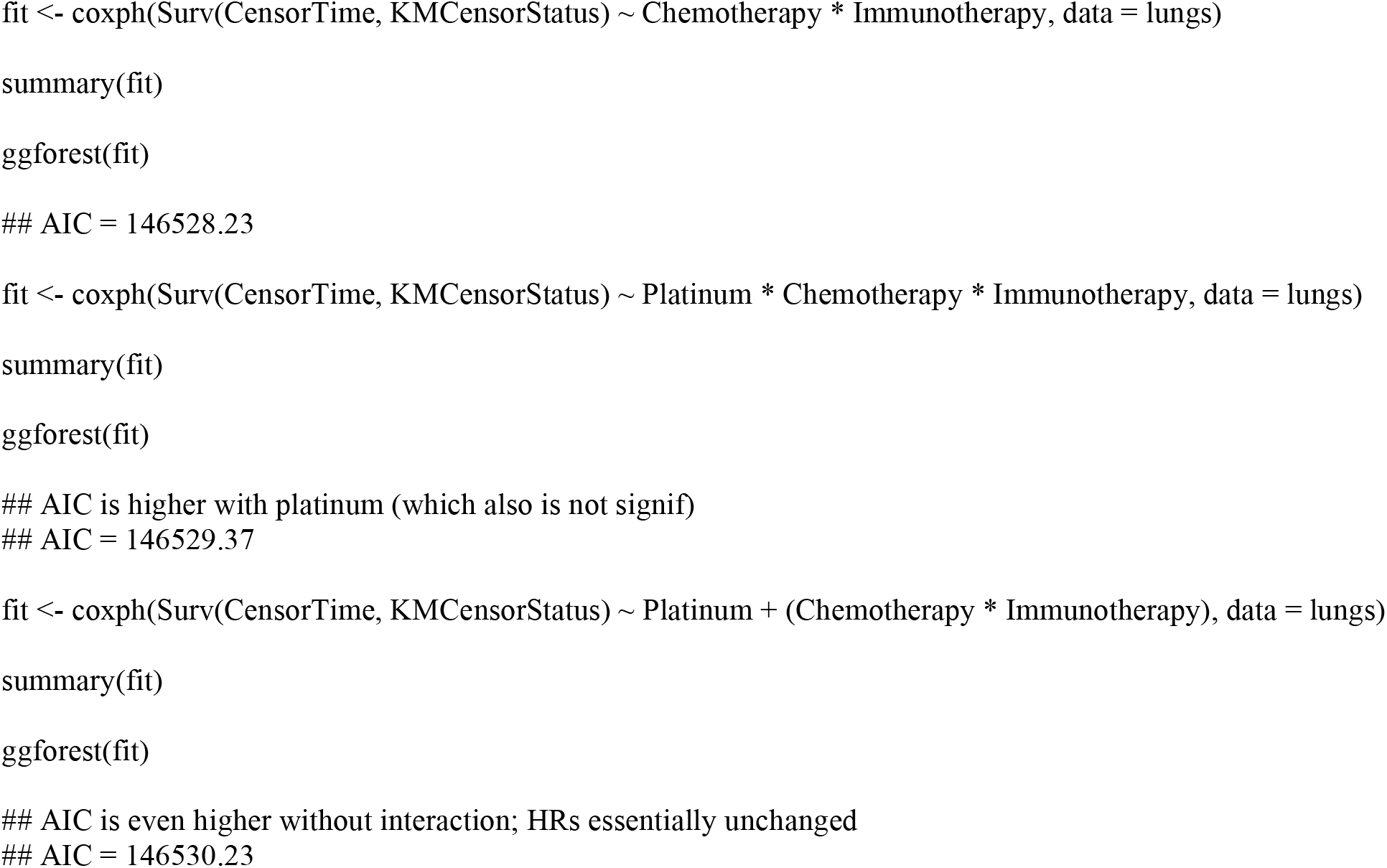

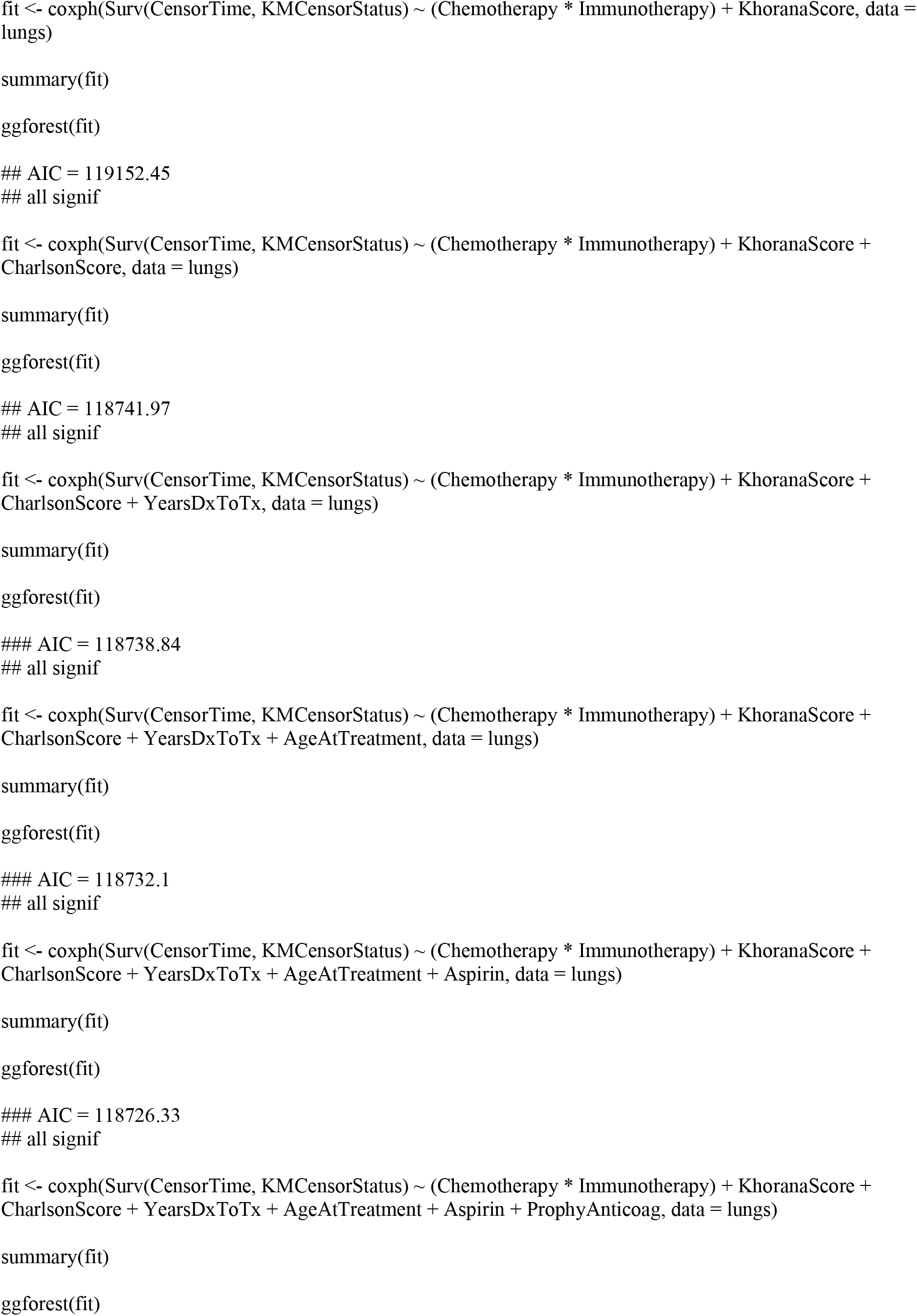

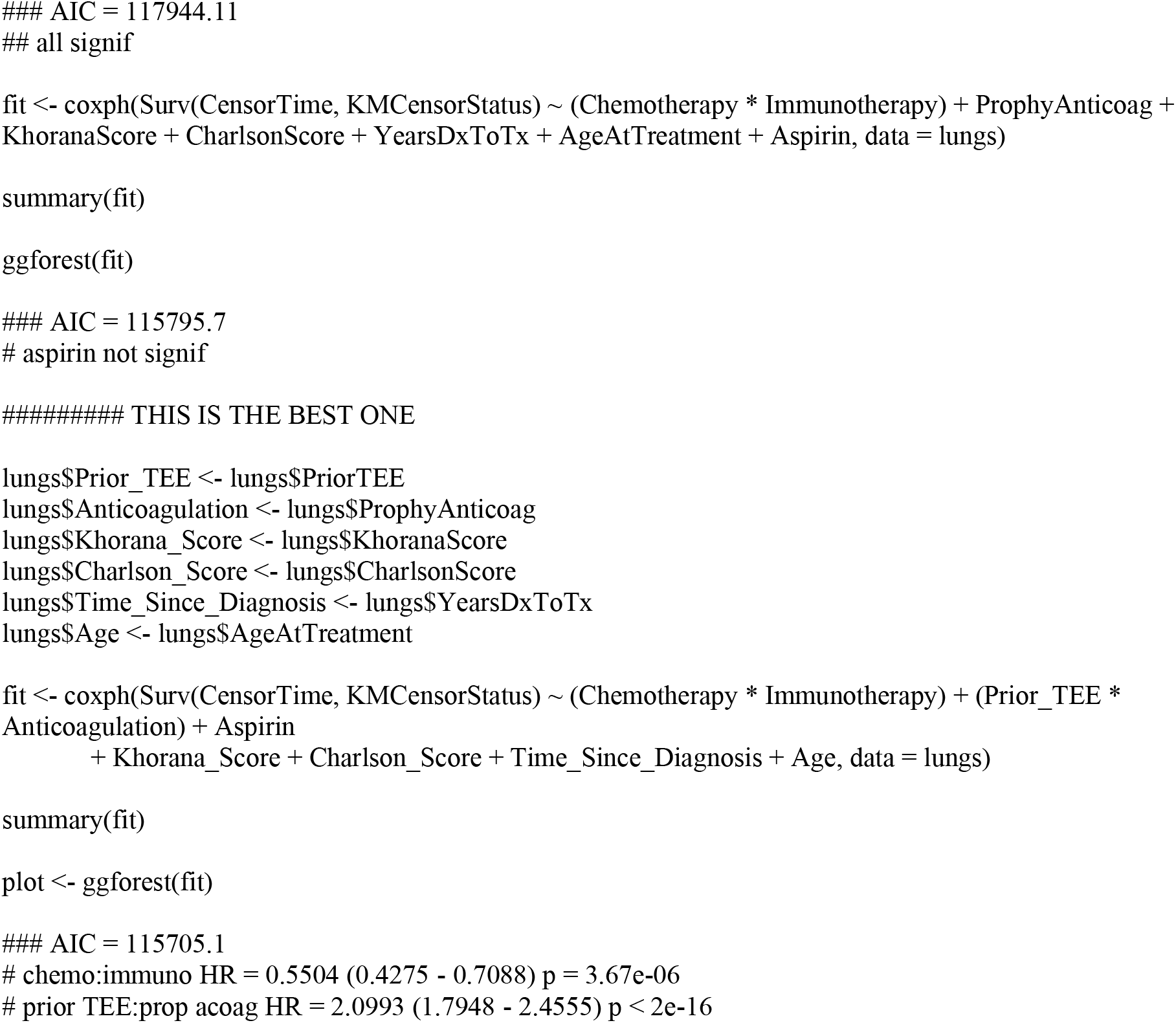

##### Perform competing risk and subdistribution hazard analyses

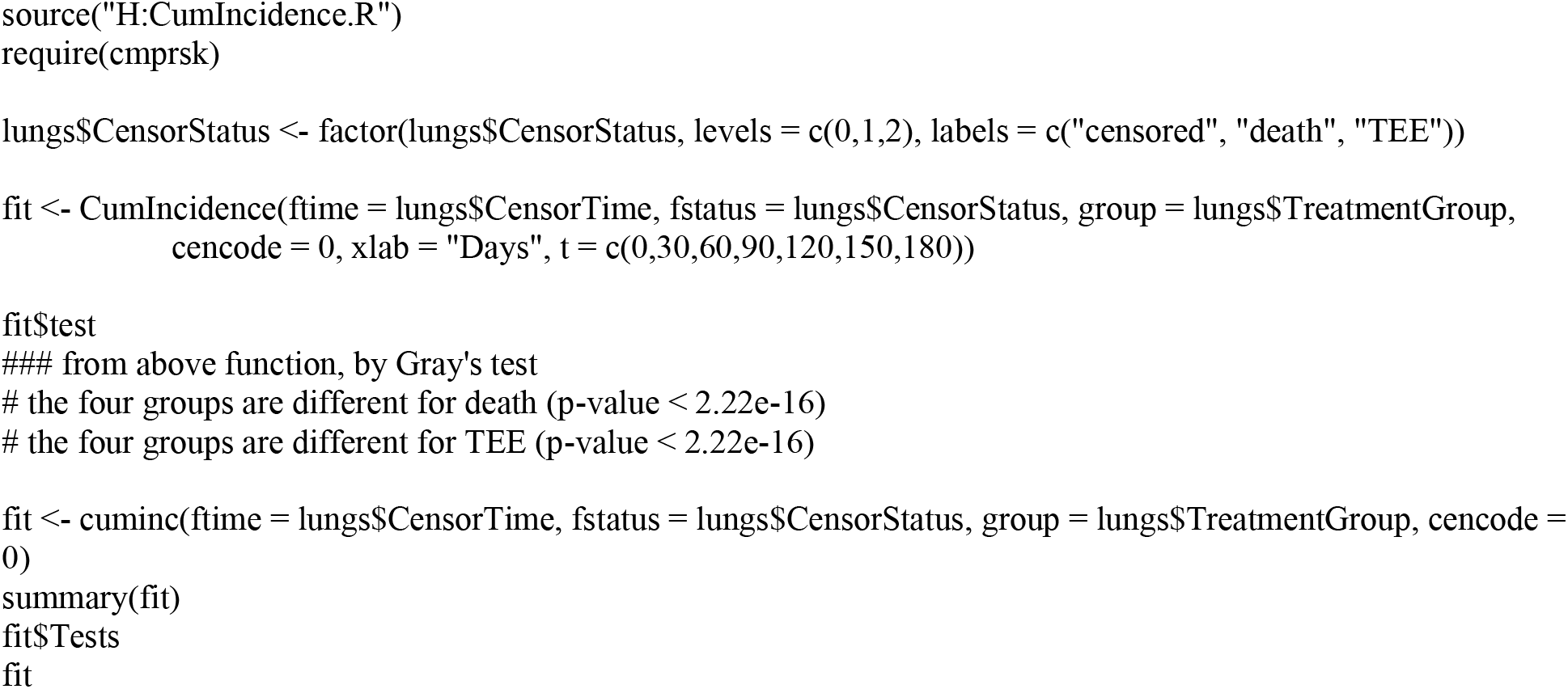

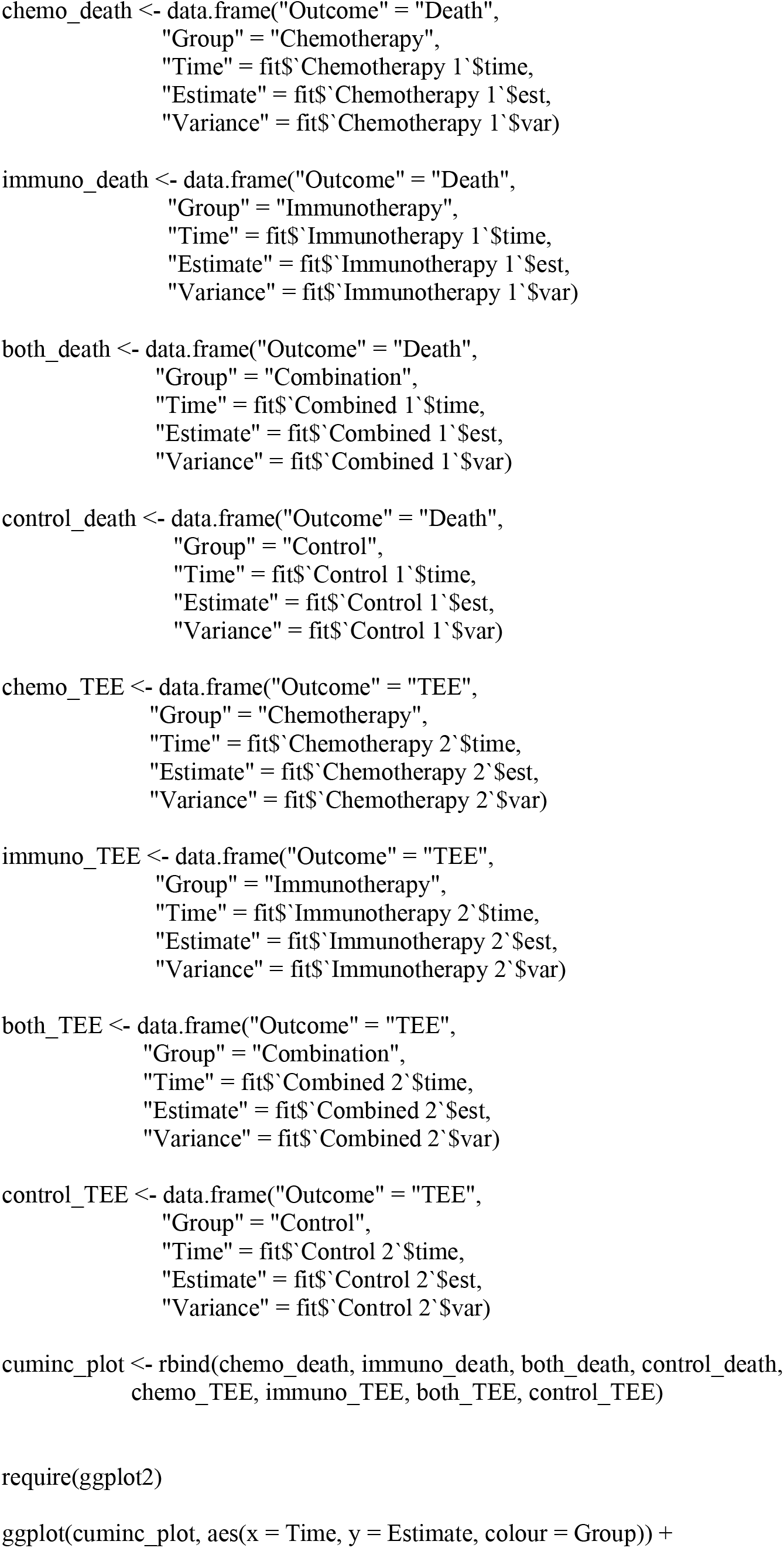

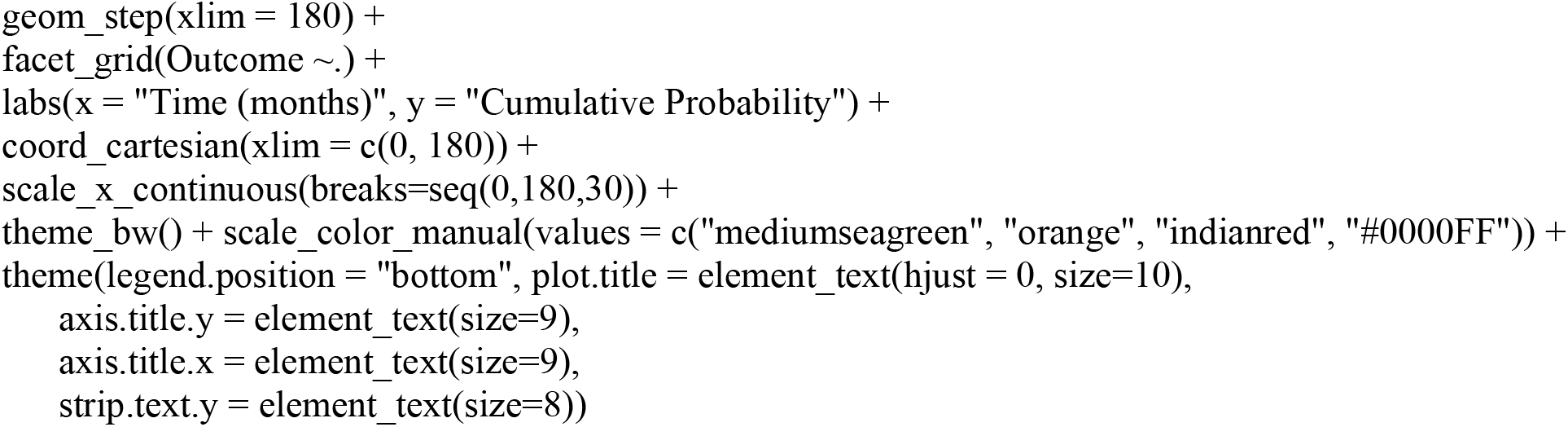

## Appendix 3

STROBE Statement—Checklist of items that should be included in reports of ***cohort studies***

|  | Item No | Recommendation |
| --- | --- | --- |
| **Title and abstract** | 1 | ☒ (a) Indicate the study's design with a commonly used term in the title or the abstract |
|  |  | ☒ (b) Provide in the abstract an informative and balanced summary of what was done and what was found |
| **Introduction** |  |  |
| Background/rationale | 2 | ☒ Explain the scientific background and rationale for the investigation being reported |
| Objectives | 3 | ☒ State specific objectives, including any prespecified hypotheses |
| **Methods** |  |  |
| Study design | 4 | ☒ Present key elements of study design early in the paper |
| Setting | 5 | ☐ Describe the setting, locations, and relevant dates, including periods of recruitment, exposure, follow-up, and data collection |
| Participants | 6 | ☒ (a) Give the eligibility criteria, and the sources and methods of selection of participants. Describe methods of follow-up |
|  |  | ☐ (b) For matched studies, give matching criteria and number of exposed and unexposed |
| Variables | 7 | ☒ Clearly define all outcomes, exposures, predictors, potential confounders, and effect modifiers. Give diagnostic criteria, if applicable |
| Data sources/measurement | 8* | ☒ For each variable of interest, give sources of data and details of methods of assessment (measurement). Describe comparability of assessment methods if there is more than one group |
| Bias | 9 | ☒ Describe any efforts to address potential sources of bias |
| Study size | 10 | ☒ Explain how the study size was arrived at |
| Quantitative variables | 11 | ☒ Explain how quantitative variables were handled in the analyses. If applicable, describe which groupings were chosen and why |
| Statistical methods | 12 | ☒ (a) Describe all statistical methods, including those used to control for confounding |
|  |  | ☒ (b) Describe any methods used to examine subgroups and interactions |
|  |  | ☐ (c) Explain how missing data were addressed |
|  |  | ☒ (d) If applicable, explain how loss to follow-up was addressed |
|  |  | ☐ (e) Describe any sensitivity analyses |
| **Results** |  |  |
| Participants | 13* | ☒ (a) Report numbers of individuals at each stage of study—eg numbers potentially eligible, examined for eligibility, confirmed eligible, included in the study, completing follow-up, and analysed<br>☐ (b) Give reasons for non-participation at each stage<br>☐ (c) Consider use of a flow diagram |
| Descriptive data | 14* | ☒ (a) Give characteristics of study participants (eg demographic, clinical, social) and information on exposures and potential confounders<br>☒ (b) Indicate number of participants with missing data for each variable of interest<br>☒ (c) Summarise follow-up time (eg, average and total amount) |
| Outcome data | 15* | ☒ Report numbers of outcome events or summary measures over time |
| Main results | 16 | ☒ (a) Give unadjusted estimates and, if applicable, confounder-adjusted estimates and their precision (eg, 95% confidence interval). Make clear which confounders were adjusted for and why they were included<br>☐ (b) Report category boundaries when continuous variables were categorized<br>☐ (c) If relevant, consider translating estimates of relative risk into absolute risk for a meaningful time period |
| Other analyses | 17 | ☒ Report other analyses done—eg analyses of subgroups and interactions, and sensitivity analyses |
| **Discussion** |  |  |
| Key results | 18 | ☒ Summarise key results with reference to study objectives |
| Limitations | 19 | ☒ Discuss limitations of the study, taking into account sources of potential bias or imprecision. Discuss both direction and magnitude of any potential bias |
| Interpretation | 20 | ☒ Give a cautious overall interpretation of results considering objectives, limitations, multiplicity of analyses, results from similar studies, and other relevant evidence |
| Generalisability | 21 | ☐ Discuss the generalisability (external validity) of the study results |
| **Other information** |  |  |
| Funding | 22 | ☒ Give the source of funding and the role of the funders for the present study and, if applicable, for the original study on which the present article is based |

\*Give information separately for exposed and unexposed groups.

---

**Supplementary Figure 1.**
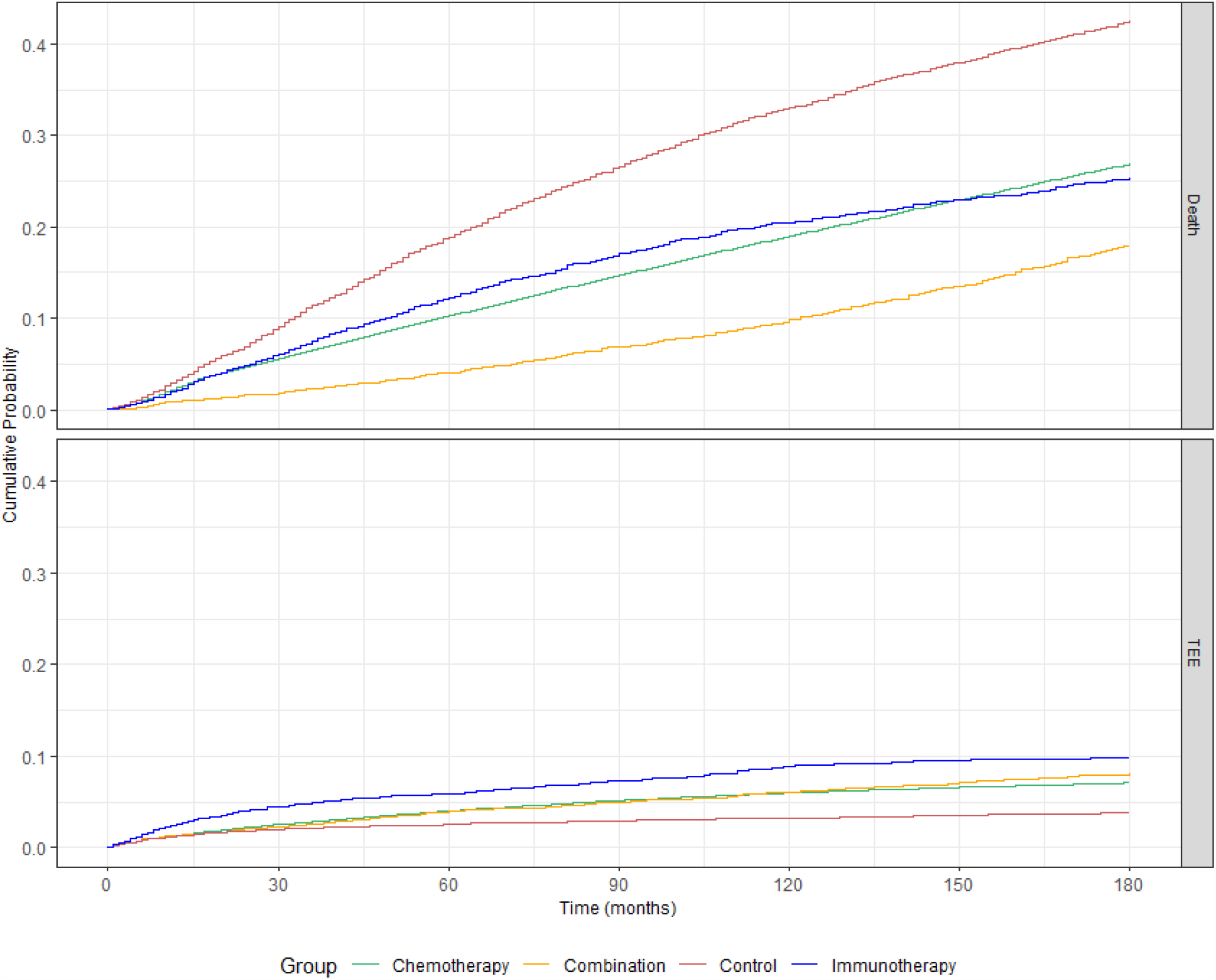
Cumulative Probability of TEE and Death Stratified by Treatment Group. Risk of experiencing death (upper panel) or TEE (lower panel) during the first 180 days after the start of treatment for lung cancer patients. The cohort is stratified by treatment group: chemotherapy alone (green), immunotherapy alone (blue), or combination of both chemo- and immunotherapies (orange), as well as a non-chemo non-immunotherapy control group (red). Time (in days from start of treatment) is shown along the x-axis, with cumulative risk along the y-axes.

**Supplementary Table 1.** Incidence of and Time to TEE Stratified by Chemotherapy Regimen. The incidences of TEE in chemotherapy patients receiving platinum-based or not platinum-based regimens did not differ significantly, but the median time to event was significantly shorter in patients receiving regimens without platinum-based agents.

|  | Platinum-based<br>n = 77,522 | Not platinum-based<br>n = 6,459 | p-value |
| --- | --- | --- | --- |
| **TEE** | 5,395 (6.96%) | 409 (6.33%) | p = 0.056 |
| **Time to event**<br>**median ± MAD** | 50 ± 53.4 | 36 ± 43.0 | p < 2.2e-16 |

P values by Wilcox test

**Supplementary Figure 2.**
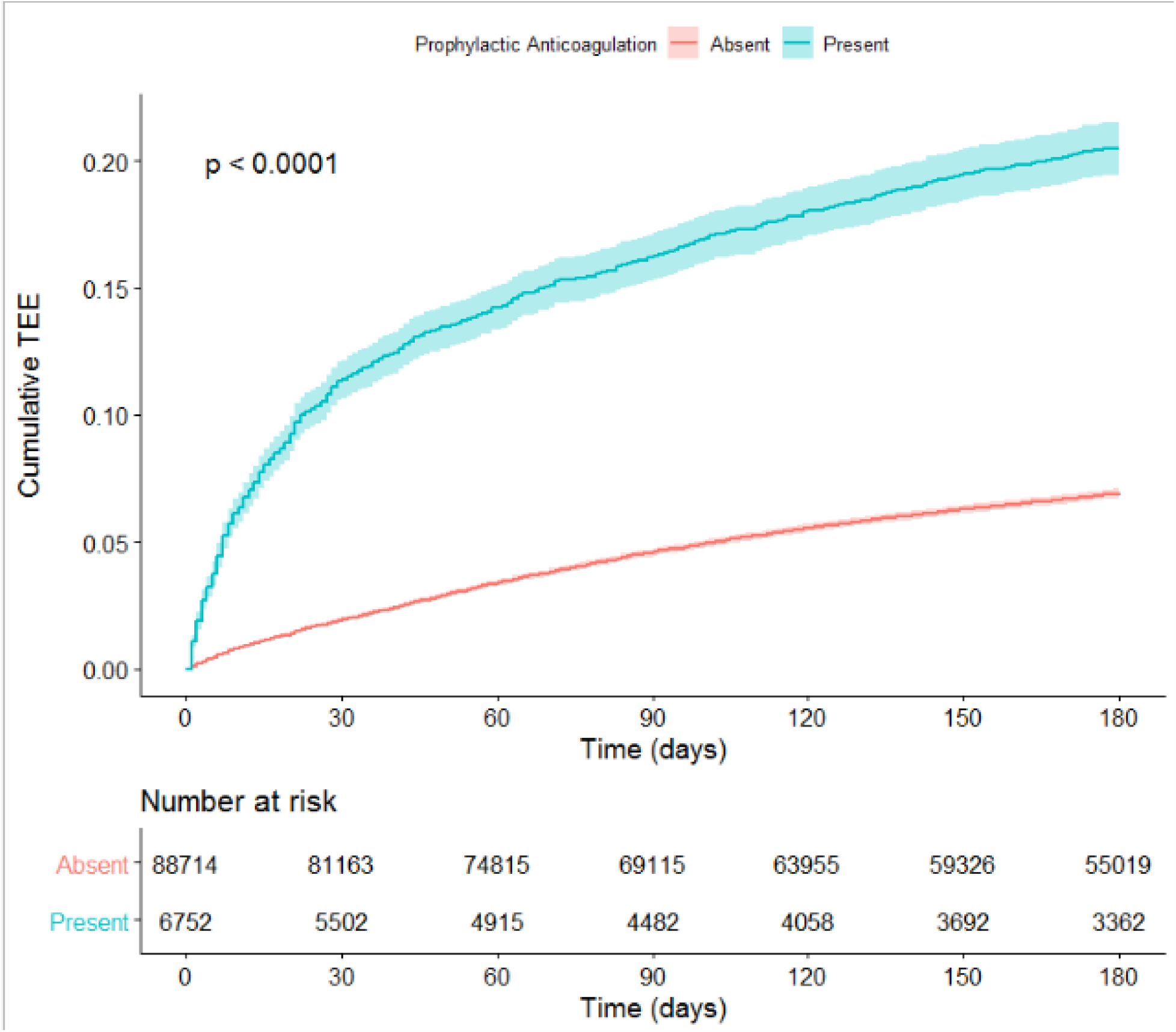
Cumulative Risk of Thrombosis in Lung Cancer Patients Stratified by Prophylactic Anticoagulation Use. Risk of experiencing a TEE during the first 180 days after the start of treatment for lung cancer patients. The cohort is stratified by prophylactic anticoagulant: presence of prophylactic anticoagulation (blue) or absence of prophylactic anticoagulation (red). Time (in days from start of treatment) is shown along the x-axis, with cumulative risk of TEE along the y-axis. The lower table depicts number of individuals at risk for each 30-day interval. P-value reflects a significant difference in cumulative risk of TEE for the two groups.

**Supplementary Figure 3.**
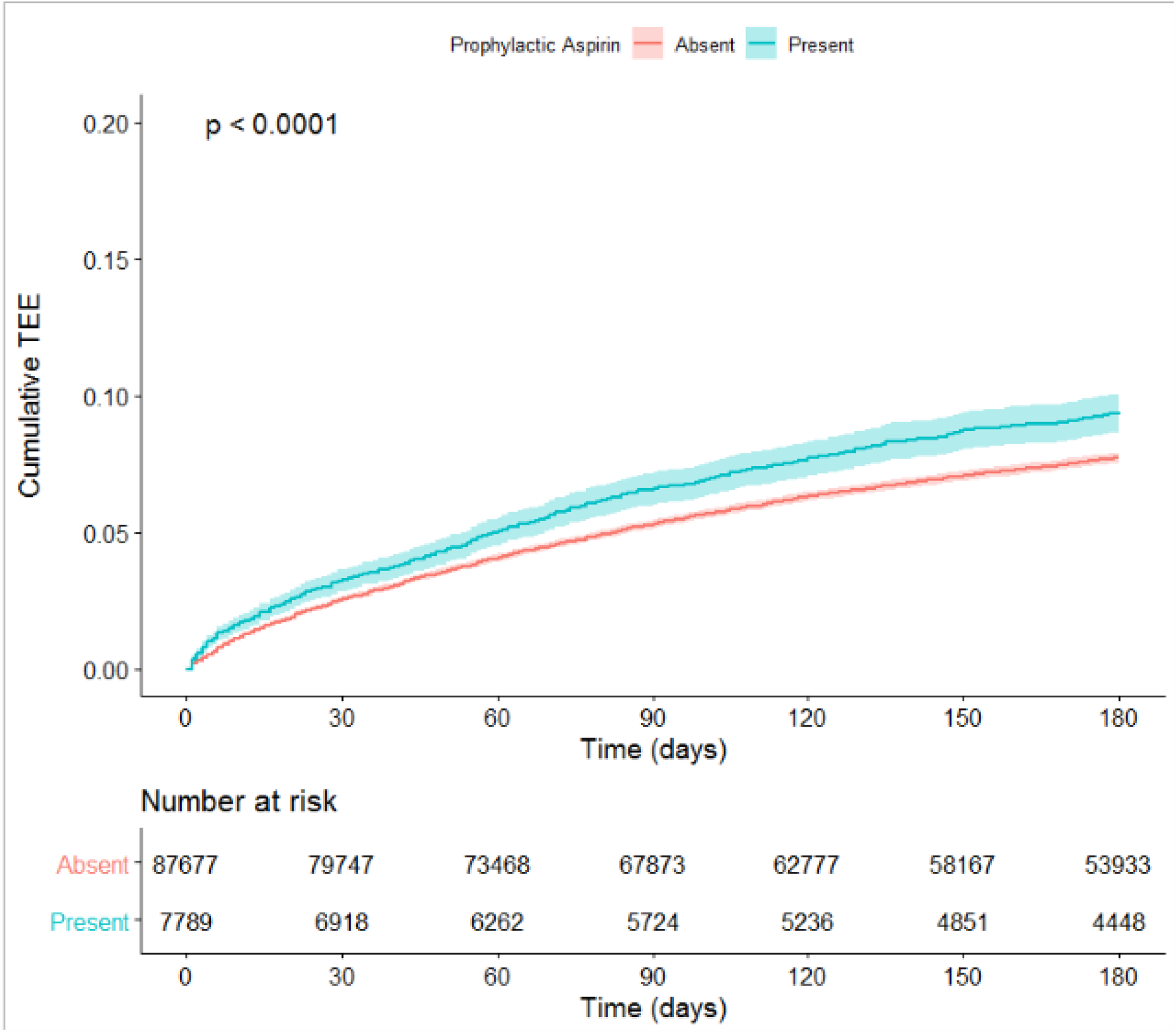
Cumulative Risk of Thrombosis in Lung Cancer Patients Stratified by Prophylactic Aspirin Use. Risk of experiencing a TEE during the first 180 days after the start of treatment for lung cancer patients. The cohort is stratified by prophylactic aspirin: presence of prophylactic aspirin (blue) or absence of prophylactic aspirin (red). Time (in days from start of treatment) is shown along the x-axis, with cumulative risk of TEE along the y-axis. The lower table depicts number of individuals at risk for each 30-day interval. P-value reflects a significant difference in cumulative risk of TEE for the two groups.

**Supplementary Figure 4.**
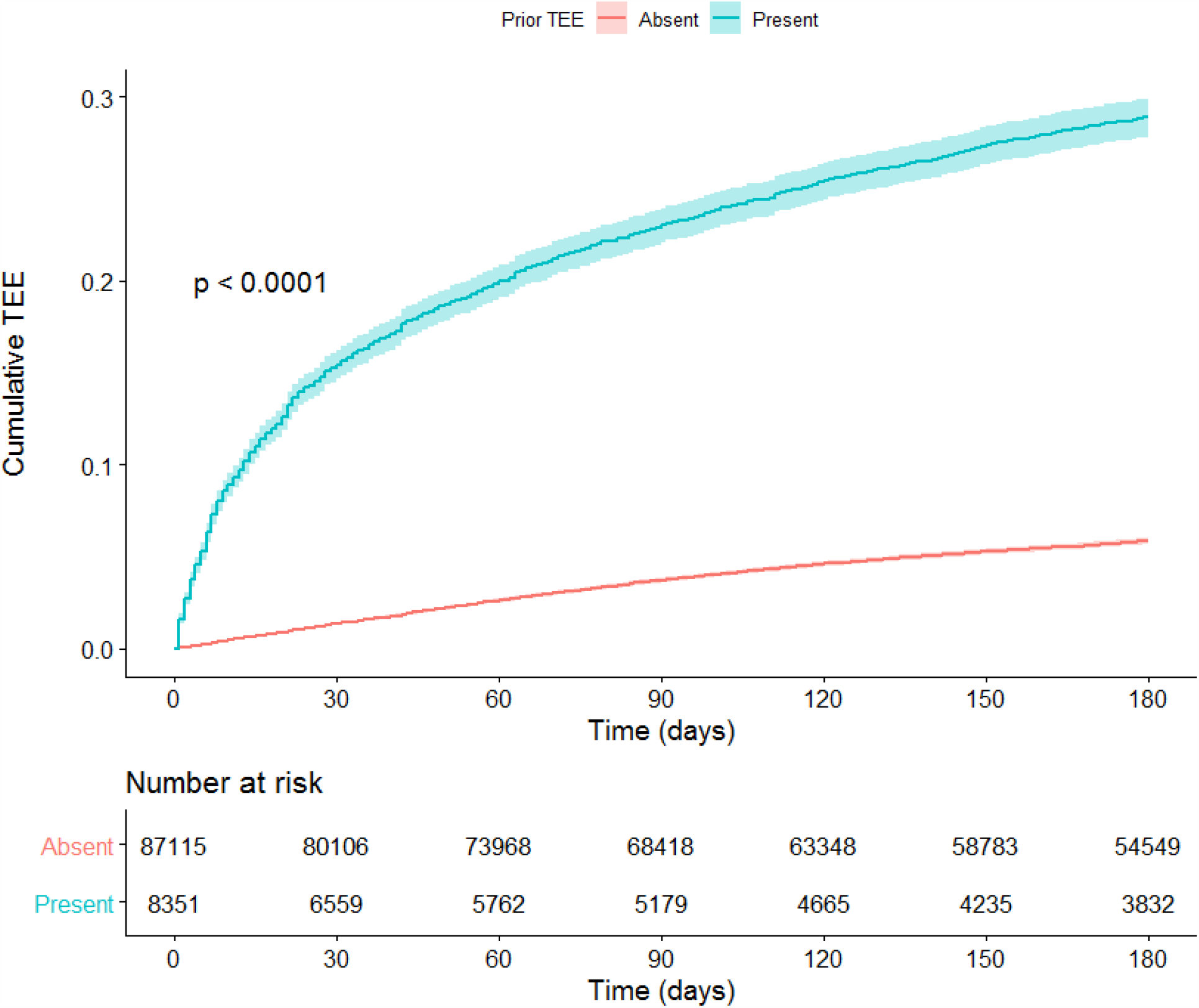
Cumulative Risk of Thrombosis in Lung Cancer Patients Stratified by History of Prior TEE. Risk of experiencing a TEE during the first 180 days after the start of treatment for lung cancer patients. The cohort is stratified by history of prior TEE: the presence thereof (blue) or the absence thereof (red). Time (in days from start of treatment) is shown along the x-axis, with cumulative risk of TEE along the y-axis. The lower table depicts number of individuals at risk for each 30-day interval. P-value reflects a significant difference in cumulative risk of TEE for the two groups.

## References

1. Ay C, Pabinger I, Cohen AT. Cancer-associated venous thromboembolism: burden, mechanisms, and management. Thromb Haemost 2017; 117: 219–230.

2. Di Nisio M, E Porreca E, Candeloro M, et al. Primary prophylaxis for venous thromboembolism in ambulatory cancer patients receiving chemotherapy. Cochrane Database Syst Rev, 12 (2016)

3. Levine MN, Gent M, Hirsh J, et al. The thrombogenic effect of anticancer drug therapy in women with stage II breast cancer. N Engl J Med 1988; 318: 404–407.

4. Saphner T, Tormey DC, Gray R. Venous and arterial thrombosis in patients who received adjuvant therapy for breast cancer. J Clin Oncol 1991; 9: 286–294.

5. Hicks LK, Cheung MC, Ding K, et al. Venous thromboembolism and non-small cell lung cancer: A pooled analysis of National Cancer Institute of Canada Clinical Trials Group trials. Cancer 2009; 115: 5516–5525.

6. Zer A, Moskovitz M, Hwang DM et al. ALK-rearranged non-small-cell lung cancer is associated with a high rate of venous thromboembolism. Clin Lung Cancer 2017; 18: 156–161.

7. Heit JA, Silverstein MD, Mohr DN, et al. Risk factors for deep vein thrombosis and pulmonary embolism: A population-based case-control study. Arch Intern Med 2000; 160: 809–815.

8. Bray F, Ferlay J, Soerjomataram I, et al. Global cancer statistics 2018: GLOBOCAN estimates of incidence and mortality worldwide for 36 cancers in 185 countries. CA Cancer J Clin 2018; 68: 394–424.

9. Cheng T-YD, Cramb SM, Baade PD, et al. The international epidemiology of lung cancer: Latest trends, disparities, and tumor characteristics. J Thorac Oncol 2016; 11: 1653–1671.

10. Ettinger DS, Wood DE, Aisner DL, et al. Non-small cell lung cancer, version 5.2017, NCCN clinical practice guidelines in oncology. J Natl Compr Canc Netw 2017; 15: 504–535.

11. Hanna N, Johnson D, Temin S, et al. Systemic therapy for stage IV non-small-cell lung cancer: American Society of Clinical Oncology clinical practice guideline update. J Clin Oncol 2017; 35: 3484–3515.

12. Novello S, Barlesi F, Califano R, et al. Metastatic non-small-cell lung cancer: ESMO clinical practice guidelines for diagnosis, treatment and follow-up. Ann Oncol 2016; 27(suppl 5): v1–v27.

13. Garon EB, Rizvi NA, Hui R, et al. Pembrolizumab for the treatment of non-small-cell lung cancer. N Engl J Med 2015; 372: 2018–2028.

14. Nelson SJ, Zeng K, Kilbourne J, Powell T, Moore R. Normalized names for clinical drugs: RxNorm at 6 years. J Am Med Inform Assoc. 2011 Jul-Aug;18(4)441–8. doi: 10.1136/amiajnl-2011-000116. Epub 2011 Apr 21. PubMed PMID: 21515544; PubMed Central PMCID: PMC3128404.

15. Khorana AA, Kuderer NM, Culakova E, et al. Development and validation of a predictive model for chemotherapy-associated thrombosis. Blood 2008; 111: 4902–4907.

16. Charlson ME, Pompei P, Ales KL, MacKenzie CR. A new method of classifying prognostic comorbidity in longitudinal studies: development and validation. J Chronic Dis 1987; 40: 373–383.

17. Sogaard M, Thomsen RW, Bossen KS et al. The impact of comorbidity on cancer survival: a review. Clin Epidemiol 2013; 5:3–29.

18. Quan H, Sundararajan V, Halfon P, et al. Coding Algorithms for Defining Comorbidities in ICD-9_CM and ICD-10 Administrative Data. Medical Care 2005; 43: 1130 – 1139.

19. Kassambara A, Kosinski M, Biecek P. suvminer: Drawing Survival Curves using ‘ggplot2’. 2020. https://CRAN.R-project.org/package=survminer

20. R Core Team. R: A language and environment for statistical computing. R Foundation for Statistical Computing. 2020. https://www.R-project.org/

21. Gray B. cmprsk: Subdistribution Analysis of Competing Risks. 2020. https://CRAN.R-project.org/package=cmprsk

22. Lesnoff M, Lancelot R. aod: Analysis of Overdispersed Data. 2012. https://CRAN.R-project.org/package=aod

23. Wickham H. ggplot2: Elegant Graphics for Data Analysis. Springer-Verlag New York, 2016.

24. Scrucca L, Santucci A, Aversa F. Regression modeling of competing risk using R: an in depth guide for clinicians. Bone Marrow Transplant 2010; 45, 1388–1395.

25. Fine JP, Gray RJ: A proportional hazards model for the subdistribution of a competing risk. J Am Stat Assoc 1999; 94:496–509.

26. von Elm E, Altman DG, Egger M, Pocock SJ, Gøtzsche PC, Vandenbroucke JP; STROBE Initiative. The Strengthening the Reporting of Observation Studies in Epidemiology (STROBE) statement: guidelines for reporting observation studies. J Clin Epidemiol 2008; 61(4):344–9.

27. Moore RA, Adel N, Riedel E, et al. High incidence of thromboembolic events in patients treated with cisplatin-based chemotherapy: A large retrospective analysis. J Clin Oncol 2011; 29: 3466–3473.

28. Jafri M, Protheroe A. Cisplatin-associated thrombosis. Anticancer Drugs 2008; 19:927–929.

29. Starling N, Rao S, Cunningham D, et al. Thromboembolism in patients with advanced gastroesophageal cancer treated with anthracycline, platinum, and fluoropyrimidine combination chemotherapy: A report from the UK National Cancer Research Institute Upper Gastrointestinal Clinical Studies Group. J Clin Oncol 2009; 27:3786–3793.

30. Lin J, Proctor MC, Varma M, Greenfield LJ, Upchurch GR Jr, Henke PK. Factors associated with recurrent venous thromboembolism in patients with malignant disease. J Vasc Surg. 2003 May;37(5):976–83.

31. Cella CA, Di Minno G, Carlomagno C, Arcopinto M, Cerbone AM, Matano E, Tufano A, Lordick F, De Simone B, Muehlberg K S, Bruzzese D, Attademo L, Arturo C, Sodano M, Moretto R, La Fata E, De Placido S. (2017). Preventing Venous Thromboembolism in Ambulatory Cancer Patients: The ONKOTEV Study. The oncologist, 22(5), 601–608.

32. Nichetti F, Ligorio F, Zattarin E, et al. Is there an interplay between immune checkpoint inhibitors, thromboprophylactic treatments, and thromboembolic events? Mechanisms and impact in non-small cell lung cancer patients. Cancers 2020; 12:67.

33. Moik F, Chan W-E, Weidemann S, et al. Incidence, risk factors, and clinical outcome of venous and arterial thromboembolic events in patients treated with immune-checkpoint inhibitors: A single-center cohort study. Res Pract Thromb Haemost 2020; 4 (Suppl 1).

34. Ibrahini S, Machiorlatti M, Vesely SK, et al. Incidence of vascular thromboembolic events in patients receiving immunotherapy: A single institution experience. Blood 2017; 130 (Suppl 1): 4864.

35. Kuderer NM, Poniewierski MS, Culakova E, et al. Predictors of venous thromboembolism and early mortality in lung cancer: Results from a global prospective study (CANTARISK). Oncologist 2018; 23: 247–255.

36. Mansfield AS, Tafur AJ, Wang CE et al. Predictors of active cancer thromboembolic outcomes: Validation of the Khorana score among patients with lung cancer. J Thromb Haemost 2016; 14: 1773–1778.

37. Noble S, Alikhan R, Robbins A, et al. Predictors of active cancer thromboembolic outcomes: Validation of the Khorana score among patients with lung cancer: Comment. J Thromb Haemost 2017; 15: 590–591.

